# Prevention of severe COVID-19 in the elderly by early high-titer plasma

**DOI:** 10.1101/2020.11.20.20234013

**Authors:** Romina Libster, Gonzalo Pérez Marc, Diego Wappner, Silvina Coviello, Alejandra Bianchi, Virginia Braem, Ignacio Esteban, Mauricio T. Caballero, Cristian Wood, Mabel Berrueta, Aníbal Rondan, Gabriela Lescano, Pablo Cruz, Yvonne Ritou, Valeria Fernández Viña, Damián Álvarez Paggi, Sebastián Esperante, Adrián Ferreti, Gastón Ofman, Álvaro Ciganda, Rocío Rodriguez, Jorge Lantos, Ricardo Valentini, Nicolás Itcovici, Alejandra Hintze, María Laura Oyarvide, Candela Etchegaray, Alejandra Neira, Ivonne Name, Julieta Alfonso, Rocío López Castelo, Gisela Caruso, Sofía Rapelius, Fernando Alvez, Federico Etchenique, Federico Dimase, Darío Alvarez, Sofía Aranda, Clara Sánchez Yanotti, Julián De Luca, Sofía Jares Baviglio, Sofía Laudanno, Florencia Nowogrodzki, Ramiro Larrea, María Silveyra, Gabriel Leberzstein, Alejandra Debonis, Juan Molinos, Miguel González, Eduardo Perez, Nicolás Kreplak, Susana Pastor Argüello, Luz Gibbons, Fernando Althabe, Eduardo Bergel, Fernando P. Polack, for the Fundación INFANT-COVID-19 Group

## Abstract

**Background:** Therapies to interrupt progression of early COVID-19 remain elusive. Among them, convalescent plasma in hospitalized patients was unsuccessful, perhaps because antibody should be administered earlier. We advanced plasma infusions to the first 72 hours of symptoms to arrest COVID-19 progression.

**Methods:** A randomized, double-blind, placebo-controlled trial of convalescent plasma with high IgG titers against SARS-CoV2 in elderly subjects within 72 hours of mild COVID-19 symptoms. The primary endpoint was severe respiratory disease defined as a respiratory rate ≥30 and/or an O_2_ sat<93% in room air. The study was interrupted at 76% of its projected sample size, because cases in the region decreased considerably and steady enrollment of study subjects became virtually impossible.

**Results:** 160 patients underwent randomization. In the intention-to-treat analysis (ITT), 13/80(16.2%) patients receiving plasma vs. 25/80(31.2%) receiving placebo experienced severe respiratory disease [RR(95%CI)= 0.52(0.29,0.94); p=0.026)] with an RRR=48%.

A modified ITT analysis, excluding six subjects who experienced the primary endpoint before infusion, showed a larger effect size [RR(95%CI) = 0.40(0.20, 0.81), p=0.007]. High- and low-titer donor analyses, based on a median IgG titer=1:3,200, evidenced a dose-dependent response with an RRR=73.3% for recipients of high-titer plasma (p=0.016) and a number needed to treat (NNT)=4.4. All secondary endpoints exhibited trends towards protection. No solicited adverse events were observed.

**Conclusions:** Early administration of high-titer convalescent plasma against SARS-CoV2 to mildly ill infected seniors reduced COVID-19 progression. This safe, inexpensive, outpatient intervention facilitates access to treatment from industrialized to LMIC, can decompress demands on hospitals, and may contribute to save lives.

**Funded by The Bill & Melinda Gates Foundation and The Fundación INFANT Pandemic Fund. Registered in the Dirección de Sangre y Medicina Transfusional del Ministerio de Salud (PAEPCC19), Plataforma PRIISA (1421), and clinicaltrials**.**gov (NCT04479163)**.

All authors have completed the ICMJE uniform disclosure form at www.icmje.org/coi_disclosure.pdf and declare: no support from any organization for the submitted work; RL, GPM, DW and FPP are investigators in a phase 3 SARS CoV2 trial from Pfizer; no other relationships or activities that could appear to have influenced the submitted work.

## Introduction

SARS-CoV2, the etiologic agent of coronavirus disease 2019 (COVID-19), elicits a particularly severe illness for the elderly, who experience high hospitalization rates and account for most fatalities worldwide [1,2]. Various comorbidities aggravate the prognosis of COVID-19 regardless of age, including hypertension, diabetes, cardiovascular disease, obesity, chronic renal failure, and chronic obstructive pulmonary disease (COPD) [1,2].

Treating COVID-19 early remains elusive. Few strategies provide some benefit, several failed, and others are under evaluation [3-6]. One of the latter strategies relies on infusion of specific antibodies present in plasma of convalescent subjects [7-9]. Plasma infusions are safe [10] and modulated other diseases in the past [11-13]. But antibodies in plasma must be administered early after infection to be effective [11-13].

To date, late infusion of convalescent plasma against SARS-CoV2 to hospitalized patients with COVID-19 has not shown clear benefits [7-9]. Therefore, we articulated a team of >250 health professionals and volunteers to address a critical question around timing of plasma as a therapeutic. The main objective of our study was to advance administration of high-titer plasma to the first days after initiation of symptoms. Here, we evaluated the efficacy of convalescent plasma with high titers of SARS-CoV2 antibody administered within 72 hours of mild symptoms to elderly subjects with COVID-19. The aim of our study was to prevent progression to severe disease.

## Methods

### Trial design and oversight

We conducted a randomized, double-blind, placebo-controlled trial between June 4 2020 and October 25 2020 (date of last patient completing follow up) at Hospitals Dr. Carlos Bocalandro, San Juan de Dios, Simplemente Evita, Central de San Isidro Dr. Melchor Ángel Posse, Clínica Olivos and a network of geriatric units in the Programa de Atención Médica Integral in the State of Buenos Aires, and Hospital Militar Central, Centro Gallego, Hospital Universitario CEMIC, and Sanatorios de los Arcos, Sagrado Corazón, Anchorena, and Finochietto in the City of Buenos Aires, in Argentina.

The trial was approved by the institutional review boards of participating institutions and the state of Buenos Aires, and registered in the Dirección de Sangre y Medicina Transfusional del MSAL (PAEPCC19; May 29 2020), Plataforma PRIISA (1421; May 17 2020), and clinicaltrials.gov (NCT04479163; July 23 2020). An independent Data and Safety Monitoring Board (DSMB) supervised the study. The authors designed the trial, compiled and analyzed the data, and vouch for their completeness and accuracy and for the adherence of the trial to the protocol.

### Trial patients

Subjects ≥75 years of age irrespective of presenting comorbidities or between 65-74 years of age with at least one comorbidity (hypertension or diabetes under pharmacologic treatment, obesity, chronic renal failure, cardiovascular disease, and COPD defined in Suppl. Material) were identified and assessed for eligibility. Eligible subjects had experienced at least one of each in the following two categories of signs and symptoms for <48 hours at the time of screening for SARS-CoV2 by reverse-transcriptase–polymerase-chain-reaction (RT-PCR): (1) a temperature ≥37.5°C and/or unexplained sweating and/or chills, and (2) dry cough, dyspnea, fatigue, myalgia, anorexia, sore throat, dysgeusia, anosmia, and/or rhinorrhea. Exclusion criteria included already presenting severe respiratory disease (our primary endpoint, see below) and/or any disease outlined in Suppl. Material.

Subjects consenting to screening were visited at home and tested in nasopharyngeal and oropharyngeal secretions by RT-PCR for SARS-CoV2 (Atila iAMP® COVID-19, Atila BioSystems). Those with detectable SARS-CoV2 RNA were transported to study hospitals and invited to sign the informed consent. Legal guardians signed consent for participating patients with cognitive impairment since July 22 2020. Starting July 27 2020, as several geriatric institutions facing SARS-CoV2 outbreaks were transformed into low-complexity units for mildly symptomatic residents, we also screened and invited residents meeting study criteria to participate in the study on site.

### Randomization and treatment regimens

After obtaining written informed consent, eligible patients were randomized to receive 250 ml of convalescent plasma with an IgG titer against SARS-CoV2 spike (S) protein >1:1,000 (COVIDAR IgG, Instituto Leloir, Argentina) or placebo (normal saline 0.9%) administered over 1.5 to 2 hours in a 1:1 ratio. Patients were randomly assigned using a computer-generated randomization sequence, with balanced permuted blocks of two. The sequence was prepared centrally at the Data Center. Both treatment and placebo were concealed using opaque bags and tape to cover the infusion line. Treatment was administered <72 hours from initiation of symptoms. Subjects were monitored for 12 hours after treatment for adverse events.

479 volunteers infected with SARS-CoV2 for a minimum of 10 days, asymptomatic for ≥3 days, and with two negative RT-PCR tests [14] were identified through hospital lists and an online campaign. Candidates were visited at home and screened for SARS-CoV2 S IgG titers >1:1,000 in serum. 135 (28.2%) were invited to donate 750 ml of plasma at four hemotherapy centers in Buenos Aires (Suppl. Material).

### Clinical and laboratory monitoring

Twenty-four hours after completing the infusion, a sample of venous blood (5 ml) was obtained from participants and preserved at −20°C until completion of the study to measure anti-S IgG SARS-CoV2 using COVIDAR IgG validated with SARS-CoV-2 Spike S1-RBD IgG ELISA Detection Kit (GenScript). In addition, we assayed samples with the SARS-CoV-2 Surrogate Virus Neutralization Test Kit (GenScript). Patient evolution was monitored daily by study physicians until day 15 to assess the primary endpoint at the hospitals, in participating geriatric institutions, or at home if patients were discharged. Those persistently symptomatic were followed until resolution of symptoms or a maximum of 25 days for secondary endpoints. Study physicians used predesigned questionnaires to collect clinical information. Data were recorded using paper forms scanned digitally due to a COVID-19 mitigation protocol and subsequently double-entered in an electronic database.

### Trial endpoints

The primary endpoint of the trial was development of severe respiratory disease defined as a respiratory rate (RR)≥30 and/or an O_2_ sat<93% when breathing room air determined between 12 hours after infusion of the investigational product (IP) and day 15 of study participation. Pre-specified secondary clinical endpoints assessed, as needed, until day 25 of study participation included (a) life threatening respiratory disease, defined as need for 100% oxygen supplementation and/or non-invasive or invasive ventilation and/or admission to intensive care; (b) critical systemic illness, defined as respiratory failure (PaO2/FiO2 ≤ 200 mm Hg) and/or shock and/or multiorganic distress syndrome (defined in Suppl. Material) and (c) death associated with COVID-19. On July 22 2020, we amended the protocol to include a fourth secondary endpoint including any of the three endpoints described above, alone or in combination.

### Early study termination

In late July, our region faced a dramatic increase in SARS-CoV2 infections and participating hospitals were overburdened with severe cases. This was compounded by an extraordinary demand for COVID-19 ambulances, which served as means of transportation for mildly symptomatic candidates from their homes to our hospitals. In addition, large groups of physician investigators infected or exposed to SARS-CoV2 were quarantined for weeks, compromising manpower. Hence, overburdening hospitals with elders presenting mild symptoms became challenging.

As several geriatric institutions facing outbreaks were transformed into low-complexity units for mildly symptomatic residents, we seek study candidates in them since July 27 2020. Eventually, COVID-19 cases in the region decreased considerably and, by late September and early October, we were enrolling one subject per week. This enrollment pace projected at best ∼5 months to complete the study in the unlikely case of the virus remaining in circulation at a constant rate. Consequently, after discussions with the DSMB and having enrolled 76% of the target population, the principal investigator (FPP) and lead sub-investigators (RL, GPM and DW) decided it would be logistically impossible, and ethically questionable given the urgency of the problem, to continue the study, and stopped to examine the results.

### Statistical analysis

Given the relative complexity of implementing this intervention, the minimally clinically important difference was set at a 40% relative reduction for an expected outcome rate of 50% in the control group reduced to 30% in the intervention group. A total sample size of 210 subjects (105 per trial arm) was estimated to have 80% power at a significance level (alpha) of 0.05 using a two-sided z-test with continuity correction.

We performed an intention-to-treat (ITT) analysis. A descriptive analysis was implemented by presentation of continuous variables as means and standard deviations or medians and interquartile ranges, as appropriate, and categorical variables as proportions.

Under the primary analysis strategy, we used the Kaplan-Meier product limit distribution to compare treatment groups for the time taken to reach the primary endpoint. An estimate of the relative risk and 95% confidence interval was also reported. A modified ITT (mITT) analysis was defined by the post-randomization exclusion of subjects that lost eligibility between randomization and treatment administration. The protocol pre-specified a subgroup analysis by two populations defined in the inclusion criteria, subjects ≥75 years of age irrespective of presenting comorbidities or between 65-74 years of age with at least one comorbidity and a second analysis of protection correlates in donors’ plasma and recipients’ sera.

## Results

### Study Population

165 subjects presented symptoms that met eligibility criteria and tested positive for SARS-CoV2 RNA (Suppl. Material). Four of them lost eligibility before enrollment and one did not consent to participate in the trial. Therefore, 160 subjects with SARS-CoV2 infection underwent randomization; 80 were assigned to receive convalescent plasma and 80 to receive placebo. Five (3.1%) patients (3 allocated to plasma and 2 to placebo) received IP after meeting the study’s primary endpoint. One additional patient randomized to receive plasma developed hypoxemia before the infusion and was not treated with IP.

160 subjects were included in the ITT analysis (Suppl. Material). One subject voluntarily abandoned the trial on day 11 of follow up. 38/160 (23.7%) subjects continued to experience COVID-19 symptoms and were followed until recovery or death for 16 to 25 days.

The mean age of subjects was 77.1 (S.D. 8.6) years; 100 (62.5%) were females (Table 1). 72 (45%) were 65-74 years and 88 (55%) ≥75 years old. Baseline characteristics showed no clinically significant imbalances between treatment and control groups. Most patients had prespecified comorbidities at enrollment (Table 1). Administration of plasma was safe, with no solicited adverse events (Suppl. Material).

**Table 1.** Characteristics of the population - ITT.

| **Variables** | **Plasma**<br>**(n=80)** |  | **Placebo**<br>**(n=80)** |  |
| --- | --- | --- | --- | --- |
| **Demographic** | **n/N** | **%** | **n/N** | **%** |
| Age in years* | 76.4 | (8.7) | 77.9 | (8.4) |
| Age by categories |  |  |  |  |
| 65-74 years | 40/80 | 50.0 | 32/80 | 40.0 |
| ≥75 years | 40/80 | 50.0 | 48/80 | 60.0 |
| Sex |  |  |  |  |
| Female | 54/80 | 67.5 | 46/80 | 57.5 |
| Male | 26/80 | 32.5 | 34/80 | 42.5 |
| **Vital signs** |  |  |  |  |
| Axillary temperature* | 36.5 | (0.7) | 36.8 | (0.8) |
| Heart Rate* | 79.8 | (13.4) | 78.6 | (14.1) |
| Systolic blood pressure* | 124.8 | (15.6) | 125.4 | (15.3) |
| Diastolic blood pressure* | 75.1 | (11.2) | 75 | (10.9) |
| Respiratory rate* | 17 | (2.8) | 17.3 | (3) |
| Oxygen saturation in room air* | 96.1 | (1.6) | 96.1 | (1.7) |
| Hours from initiation of symptoms* | 39.6 | (13.9) | 38.3 | (14.3) |
| **Comorbidities** |  |  |  |  |
| Hypertension under treatment | 62/80 | 77.5 | 52/80 | 65 |
| Diabetes under treatment | 23/80 | 28.7 | 13/79 | 16.5 |
| Obesity | 4/80 | 5.0 | 8/79 | 10.1 |
| COPD under treatment | 2/80 | 2.5 | 5/79 | 6.3 |
| Cardiovascular disease** | 14/80 | 17.5 | 7/79 | 8.9 |
| Chronic renal failure | 1/80 | 1.2 | 3/79 | 3.8 |
| At least one major comorbidity | 69/80 | 86.2 | 62/80 | 77.5 |
| **Secondary comorbidities** |  |  |  |  |
| Asthma or other respiratory disease | 3/80 | 3.8 | 3/80 | 3.8 |
| Non-cirrhotic liver disease | 0/80 | 0.0 | 0/80 | 0.0 |
| Cancer (not active) | 4/80 | 5.0 | 2/80 | 2.5 |
| Neurologic disease | 11/80 | 13.8 | 9/80 | 11.2 |
| History of smoking | 13/80 | 16.2 | 10/80 | 12.5 |
| Receives medications regularly | 76/80 | 95.0 | 73/80 | 91.2 |
| Received medications in last 15 days | 71/80 | 88.8 | 64/80 | 80.0 |

### Primary Endpoint

In the ITT analysis, 13/80 (16.2%) patients receiving plasma vs. 25/80 (31.2%) receiving placebo experienced severe respiratory disease [RR (95%CI) = 0.52 (0.29,0.94); p=0.026)] (Table 2). In the time to event analysis, patients in the plasma group also had a longer time to development of severe COVID-19 than patients receiving placebo [median (Q1-Q3) = 15 (15-15) vs. 15 (8.8-15); p=0.028 in Figure 1]. The relative risk reduction (RRR) of convalescent plasma was 48% and the NNT to avert an episode of severe COVID-19 in the population was 6.67. Pre-specified exploratory analyses by age group demonstrated a strong effect for plasma in subjects ≥75 years of age both in proportions [RR (95%CI) = 0.35 (0.14,0.87), p=0.014] and time to event (p=0.016; Suppl. Material).

**Table 2.**
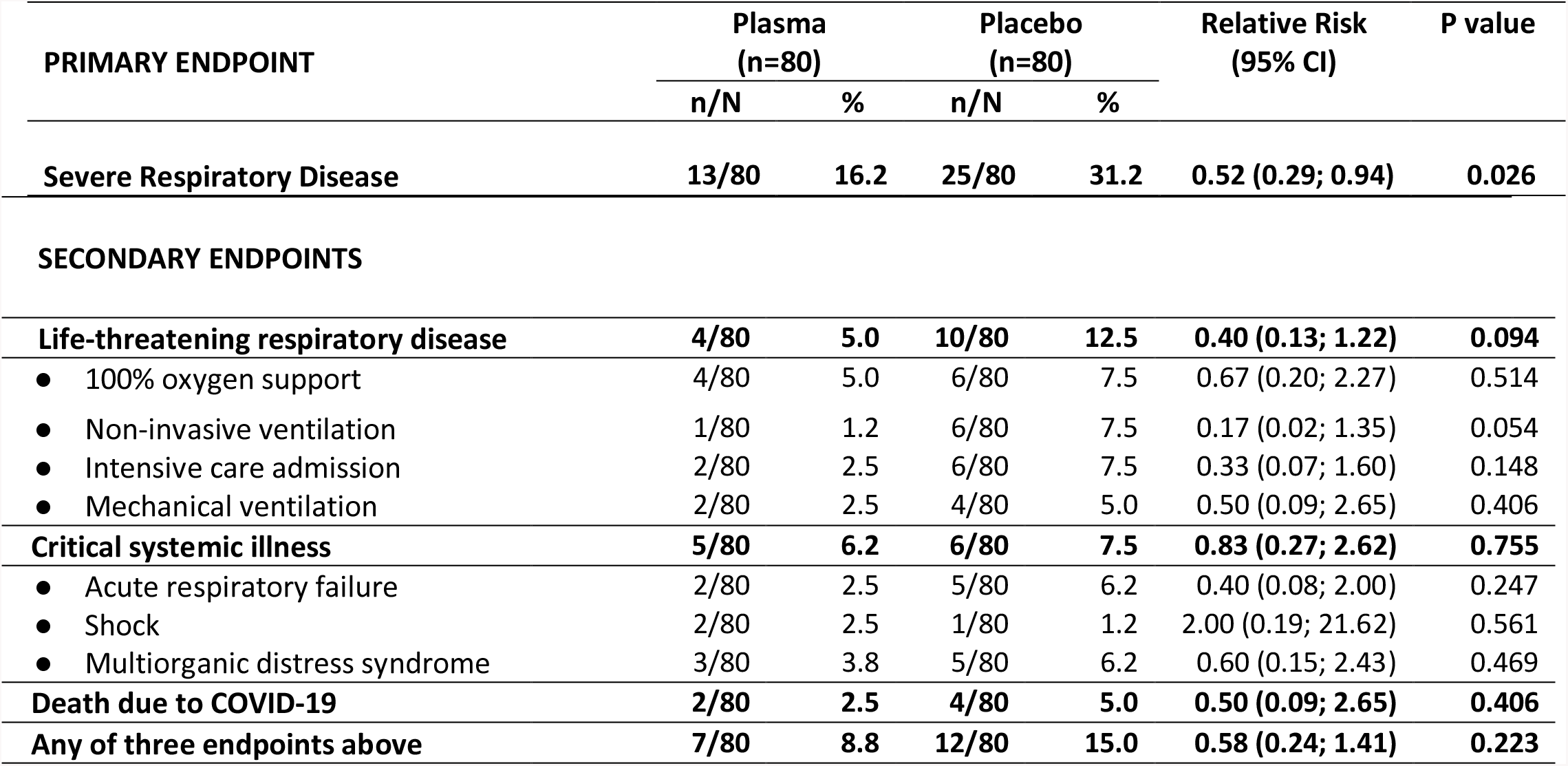
Study Endpoints. Proportion of cases – ITT.

| PRIMARY ENDPOINT | Plasma<br>(n=80) |  | Placebo<br>(n=80) |  | Relative Risk<br>(95% CI) | P value |
| --- | --- | --- | --- | --- | --- | --- |
|  | n/N | % | n/N | % |  |  |
| **Severe Respiratory Disease** | **13/80** | **16.2** | **25/80** | **31.2** | **0.52 (0.29; 0.94)** | **0.026** |
| **SECONDARY ENDPOINTS** |  |  |  |  |  |  |
| **Life-threatening respiratory disease** | **4/80** | **5.0** | **10/80** | **12.5** | **0.40 (0.13; 1.22)** | **0.094** |
| • 100% oxygen support | 4/80 | 5.0 | 6/80 | 7.5 | 0.67 (0.20; 2.27) | 0.514 |
| • Non-invasive ventilation | 1/80 | 1.2 | 6/80 | 7.5 | 0.17 (0.02; 1.35) | 0.054 |
| • Intensive care admission | 2/80 | 2.5 | 6/80 | 7.5 | 0.33 (0.07; 1.60) | 0.148 |
| • Mechanical ventilation | 2/80 | 2.5 | 4/80 | 5.0 | 0.50 (0.09; 2.65) | 0.406 |
| **Critical systemic illness** | **5/80** | **6.2** | **6/80** | **7.5** | **0.83 (0.27; 2.62)** | **0.755** |
| • Acute respiratory failure | 2/80 | 2.5 | 5/80 | 6.2 | 0.40 (0.08; 2.00) | 0.247 |
| • Shock | 2/80 | 2.5 | 1/80 | 1.2 | 2.00 (0.19; 21.62) | 0.561 |
| • Multiorganic distress syndrome | 3/80 | 3.8 | 5/80 | 6.2 | 0.60 (0.15; 2.43) | 0.469 |
| **Death due to COVID-19** | **2/80** | **2.5** | **4/80** | **5.0** | **0.50 (0.09; 2.65)** | **0.406** |
| **Any of three endpoints above** | **7/80** | **8.8** | **12/80** | **15.0** | **0.58 (0.24; 1.41)** | **0.223** |

**Figure 1.**
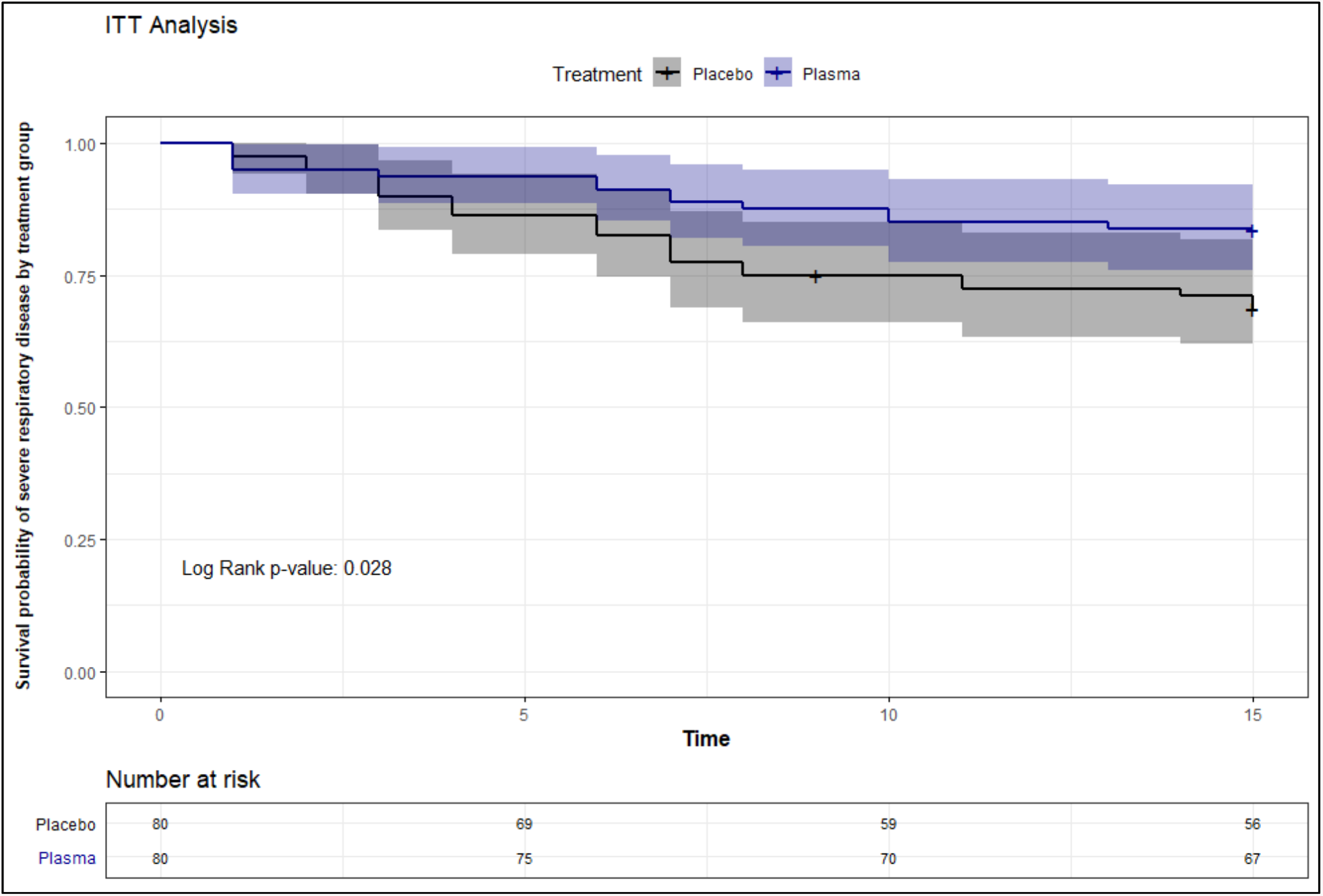
Kaplan-Meier survival curve of severe respiratory disease due to SARS-CoV2 by treatment group in days – ITT Analysis.

Four subjects in the intervention group and two subjects in the control group lost eligibility by reaching the primary endpoint before receiving the IP. A clinically relevant, modified ITT population was defined by excluding these subjects. The results of the mITT analysis showed a larger intervention effect size, with 9/76 (11.8%) patients receiving plasma vs. 23/78 (29.5%) receiving placebo experiencing severe respiratory disease [RR (95%CI) = 0.40 (0.20, 0.81), p=0.007]. In the mITT population, patients in the plasma group also had a longer time to development of severe COVID-19 than patients receiving placebo (p=0.006; Suppl. Material).

### Secondary Endpoints

Results for secondary endpoints are presented in Table 2. Four (5%) plasma recipients and 10 (12.5%) placebo recipients experienced life-threatening respiratory disease, while 5 (6.2%) and 6 (7.5%) had critical systemic illness, respectively. 2 plasma vs. 4 placebo recipients died. The combined secondary endpoint (life-threatening respiratory and/or critical systemic disease and/or death) was met by 7 (8.8%) subjects treated with plasma vs. 12 (15%) treated with placebo. Secondary endpoint results for the mITT analysis are presented in Suppl. Material.

### Antibody titers

The distribution of serum titers 24 hours after infusion in plasma vs. placebo recipients differed significantly, with higher concentrations in subjects allocated to the intervention (log median (Q1-Q3) = 5.7 (4.9-6.3) vs. 3.9 (3.9-4.7); p<0.0001; Fig.2). Comparison between severe and mild cases detected no correlate of protection in plasma recipients’ sera 24 hours after infusion (Suppl. Material).

**Figure 2.**
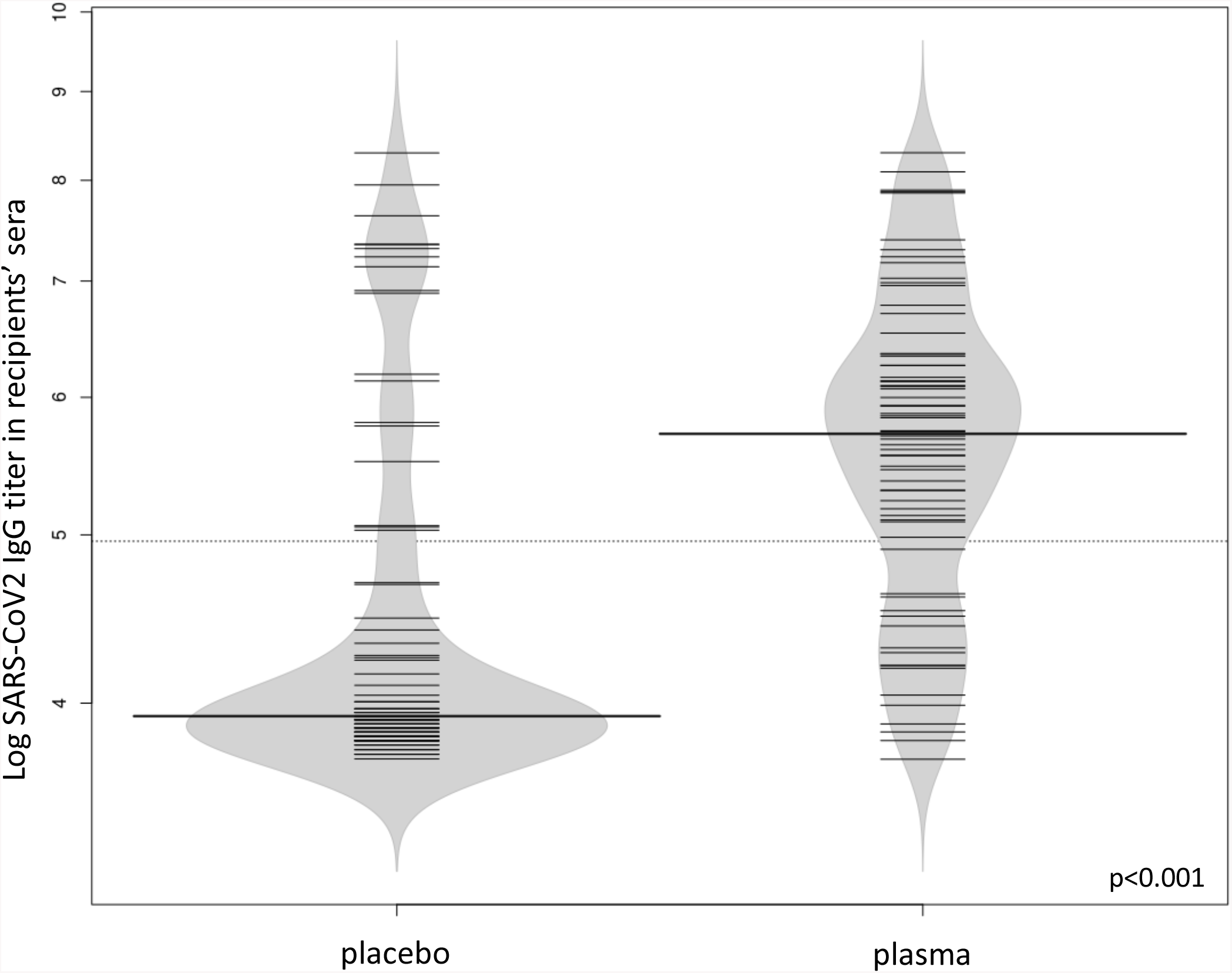
SARS-CoV2 serum titers in infected elderly subjects (violin plot).

Conversely, a dose-dependent effect was evident for SARS-CoV2 S IgG titers in plasma bags (Table 3). Analysis of high and low titer donors classified based on a median titer of 1:3,200, showed a RRR=73.3% with a NNT=4.4 for recipients of high titer products (p=0.016; Table 3). SARS-CoV2 S IgG results were replicated using a different SARS-CoV-2 Spike S1-RBD IgG commercial assay providing a potential alternative tool for donor selection (r=0.7; p<0.001 in Suppl. Material).

**Table 3.** Primary Endpoint by Donor SARS-CoV2 IgG Titer.

|  | **n** | **N** | **Rate** | **RR** | **95% CI** |  | **p value** | **RRR** |
| --- | --- | --- | --- | --- | --- | --- | --- | --- |
| **Placebo** | 25 | 80 | 31.3% | 1.00 |  |  |  |  |
| **Donor Plasma High\*** | 3 | 36 | 8.3% | 0.27 | 0.08 | 0.68 | 0.016 | 73.3% |
| **Donor Plasma Low** | 9 | 42 | 21.4% | 0.69 | 0.34 | 1.31 | 0.274 | 31.4% |

\* using a median SARS CoV2 IgG titer $\geq 1:3,200$ .

## Discussion

In this manuscript, we report the first specific treatment against early COVID-19 of potential global access and low relative cost. Early administration of plasma with high titers of antibody against SARS-CoV2 to infected seniors reduced progression to severe COVID-19 by 48%. Selecting plasma with IgG titers above a median of 1:3,200 increased protection to 73%. Our findings stress the need to return to the old paradigm of treating acute viral infections early and define IgG targets that facilitate donor selection.

In our study, timing and disease severity at the time of treatment differed from previous reports. A randomized trial of convalescent plasma vs. standard treatment in hospitalized patients with severe or life-threatening COVID-19 in China did not impact clinical improvement [7]; a second randomized trial in Indian hospitals found no differences in progression to life-threatening disease and mortality between plasma and placebo recipients [9]; and a third study did not show significant differences in probability of death for severely ill patients using propensity score-matched controls [8].

Early high-titer plasma administration is a therapeutic breakthrough, as alternatives against COVID-19 are scarce. The monoclonal antibody LY-CoV555 has been shown to decrease viral load in recently diagnosed mild or moderately-ill outpatients [15], remdesivir shortens the time to recovery in hospitalized patients with COVID-19 [16], and dexamethasone decreases mortality in severely ill subjects [17]. Convalescent plasma can bridge the transition to accessible vaccines as a potentially inexpensive (per patient cost in our region=u$186.25; Suppl. Material), outpatient, effective intervention with reasonably rapid access for many LMIC.

Six subjects reached the primary endpoint before receiving IP. To provide plasma of all blood types to 15 institutions, we instrumented two unblinded infusion teams that drove from a central hemotherapy station harboring the product to all hospitals after randomization. The study region was >100 miles^2^ and security challenges precluded access to several hospitals after 8 pm. Exclusion of these patients in the mITT analysis increased the efficacy of the intervention to 60%. Since plasma infusions are safe and relatively easy to administer, and acknowledging that situations may vary in different regions, implementation may work best at designated outpatient clinics with donations fostered at a local level reinforcing a sense of community kinship. Therefore, we speculate that efficacy may approach the mITT in many settings or ascend >70% if donors with IgG titers ≥1:3,200 are selected. The strong correlation between assays suggests that other S/RBD IgG immunoassays may provide reasonable alternative screening tools for implementation (Suppl. Material). Enhancing early symptom awareness in seniors will be vital, now that there is a time-limited effective intervention available. Plasma against COVID-19 is conceptually like health insurance. It should be in-hand when it intuitively seems unnecessary.

Our study demonstrates a dose-dependent IgG effect in plasma infusions. Plasma with IgG titers ≥1:3,200 reduced the risk of severe COVID-19 by 73%. Since IgG concentrations, and not volume, are critical for therapeutic success, numerous “super-donors” with IgG titers ≥1:12,800 could help >20 subjects by providing 750 ml. Maintenance of high IgG titers for months stress the value of fostering their loyalty for recurrent donations [18]. Interestingly, 71% of donors ≥1:3,200 among our volunteers had been hospitalized (not shown).

In summary, administration of select convalescent plasma against SARS-CoV2 to infected seniors within 72 hours of mild symptoms reduced COVID-19 progression to severe illness in a RCT. This simple, safe and inexpensive intervention can be rapidly available, decompress demands on the health care system, and may save lives. Early plasma infusions can provide a bridge to the future, until vaccines become widely available.

## Supporting information

Protocolo Original

Translated Protocol

Analysis Plan

## Data Availability

Individual participant data that underlie the results reported in this article, after de-identification, will be shared. Proposals should be directed to.

## FUNDACION INFANT- COVID-19 GROUP

**Fundación INFANT**. Florencia A. Izetta, María Teresa Panighetti, Paula Fernández Estrella, María E. Gutiérrez Meyer, Viviana Dominguez, Marcela Balduzzi, Romina Militerno, Jimena Ochoa, Sebastián Pérez Marc, Lucila Di Nunzio, Mariano Aizpurúa, Romina Zadoff, Carla Marchionatti, Natalia García Escude.

**Hospital Militar Central**. Romina Romero, Noelia Iraizos, Emmanuel Valls, Patricia Rearte Carvalho, Jimena Franco, Natali Estrada, Juan Rusconi, Guido A. Ochoa.

**Swiss Medical Group**. María Verónica Paz, Patricia Lesch.

**Hospital Dr. Carlos Bocalandro**. Fernanda Caracciolo, María Eugenia Macaneo.

**Centro Gallego**. Lia Fogos.

**Hospital San Juan de Dios**. Silvana Marquez, Gastón Pellegrino.

**Facultad de Medicina en la Universidad de Buenos Aires**. Jorge Geffner, Rocío B. Zarlenga, Camila H. Witteveen, Agustina Venditti, Indira Pichetto Olanda, Juan M. Vargas, Micaela A. Piani, Daniela C. Galnares, Florencia de la Fuente Balcarcel.

**Instituto Leloir**. Andrea Gamarnik.

**Hospital Simplemente Evita**. Maria del Carmen Nigro, Susana Villaroel.

**CEMIC**. Cristina Soler Riera.

**Hospital Municipal San Isidro**. Leonel Langelotti, Clarisa Taffarel.

**Sanatorio Anchorena**. Jose L. Scapellato.

**Sanatorio Sagrado Corazón OSECAC**. Mariano Girassolli, Maximiliano de Zan.

**Ministerio de Salud de la Provincia de Buenos Aires**. Juan Sebastian Riera, Enio Garcia, Mario Rovere, Juan Canela, Agostina Pagella, Cecilia Pampuro, Alfonso Raggio.

**Hospital J**.**P. Garrahan**. Silvina Kuperman

**Programa de Atención Médica Integral (PAMI)**. Yanina Mirayaga.

## ACKNOWLEDGEMENTS

We thank Ana L. Ayrolo, Paula Cipriani, Veronica Bianchi, Omar Lavieri, Cecilia Riera Sala, Carola Candurra, Roxana Olivera, Emiliano Sosa, Daniel Gollan, Fernan Quiros, Luana Volnovich, Alejandro Aimar, Mariana Bertolini, Daniel Stamboulian, Juan M. Rey Liste, Cristian Werb, Gerry Garbulsky, Alan Gegenschatz, Luciana Armengol, Adriana Romeo, Alejandra Castro, Alfredo de Monte, Ana La Rosa, Ana María Chiaro, Ana Stilman, Andrea Argüello, Andrea Churba, Andrea Di Fabio, Andrea Gonzalez, Carolina Castro, Carolina Chicote, Carolina Hardoy, Carolina Vairo, Cecilia Fernandez Parmo, Cecilia Masdeu, Daniel Picciola, Darío Ibañez, Enriqueta Chmielecki, Estela Kalinsky, Fabian Galperin, Felisa Rodríguez Palma, Florencia Martinez Pedemonte, Raúl A. Gómez, Gabriela Lombardi, Gisela Kremenchuzky, Gustavo Rizzo, Nelson Donato, Iván Urlich, Jonathan Cohen, Jorge Aguirre, Jorgelina Centeno, Juan Martín Redondo, Leandro Stitzman, Maria Cecilia Arias, Marcela Baldoni, Marcela Oksengendler, Marcela Testoni, Marcelo Suarez, María de las Mercedes Solis, Mariana Levy, María Eugenia Segretin, Maria Jose Sotti, María Laura Alzúa, María Soledad Oporto, Mariana Dunaiewsky, Mariana Loban, Marta Wydra, Miguel Angel Patane, Mirta Ludueña, Natalia Gitelman, Pablo Lopez, Pablo Rush, Paula Pini, Sandra Figoni Prado, Sandra Jiménez, Silvana Franco, Silvina Bosco, Silvina Moreno, Silvina Vallieri, Valeria Rios, Veronica Siciliano, Verónica Zlotogorski, Raul A. Gaivironsky, Juan Minatta, Grupo Pediátrico at HMC, and participating patients and families for their invaluable help.

The Fundacion INFANT Pandemic Fund received contributions from Laboratorio Roemmers, Bodega Vistalba, Swiss Medical Group, Laboratorio Bago, Laboratorio Raffo, Laboratorios Monserrat y Eclair, Tuteur Sacifia, TASA Logistica, Fundación IRSA, Puerto Asís Investments, Fundación Hematológica Sarmiento, Alec Oxenford, Carlos Kulish and family, Renato Montefiore and family, Irene Gorodisch, Alejandro Gorodisch, the Braun family, Agustín Otero Monsegur, and Luis Roque Otero.

We are particularly indebted to the members of the DSMB Dr. Gilda Piaggio, Dr. Jorge Hevia, and Dr. Roberto Freue.

