## Supplemental Material for "Prevention of severe COVID-19 in the elderly by early high-titer plasma"

### SUPPLEMENTARY MATERIAL

Libster R, Perez Marc G, Wappner D et al. Prevention of severe COVID-19 in infected elderly by early administration of plasma with antibody against SARS-CoV2

#### SUPPLEMENTARY MATERIALS INDEX

**Table 1.** DEFINITIONS FOR COMORBIDITIES **(Page 4)**

**Table 2.** HEART FAILURE **(Page 5)**

**Table 3.** CIRRHOSIS (PORTAL BAVENO IV HYPERTENSION CONSENSUS) **(Page 6)**

**Table 4.** KIDNEY DISEASE CLASSIFICATION KDIGO **(Page 7)**

**Table 5.** EXCLUSION CRITERIA **(Page 8)**

**Table 6.** SOLICITED ADVERSE EVENTS – ITT **(Page 9)**

**Table 7.** PROPORTION OF PRIMARY ENDPOINT CASES BY AGE GROUP – ITT **(Page 10)**

**Table 8.** CHARACTERISTICS OF THE POPULATION – mITT **(Page 11)**

**Table 9.** PRIMARY ENDPOINT. PROPORTION OF CASES – mITT **(Page 12)**

**Table 10.** PRIMARY ENDPOINT. DAYS TO EVENT – mITT **(Page 13)**

**Table 11.** SECONDARY ENDPOINTS: CASE PROPORTIONS –ITT **(Page 14)**

**Table 12.** PROPORTION OF PRIMARY ENDPOINT CASES BY AGE GROUP – mITT **(Page 15)**

**Table 13.** COST OF A PLASMA INFUSION IN BUENOS AIRES, ARGENTINA PER PATIENT **(Page 16)**

**Figure 1.** CONSORT **(Page 17)**

**Figure 2.** KAPLAN-MEIER SURVIVAL CURVE OF SEVERE RESPIRATORY DISEASE DUE SARS-COV2 IN 65-74 YEARS OLD BY TREATMENT GROUP – ITT ANALYSIS **(Page 18)**

**Figure 3.** KAPLAN-MEIER SURVIVAL CURVE OF SEVERE RESPIRATORY DISEASE DUE SARS-COV2 IN ≥75 YEARS OLD BY TREATMENT GROUP – ITT ANALYSIS **(Page 19)**

**Figure 4.** KAPLAN-MEIER SURVIVAL CURVE OF SEVERE RESPIRATORY DISEASE (PRIMARY ENDPOINT) DUE TO SARS-COV2 BY TREATMENT GROUP – mITT ANALYSIS **(Page 20)**

**Figure 5.** KAPLAN-MEIER SURVIVAL CURVE OF SEVERE RESPIRATORY DISEASE DUE SARS-COV2 IN 65-74 YEARS OLD BY TREATMENT GROUP – mITT ANALYSIS **(Page 21)**

**Figure 6.** KAPLAN-MEIER SURVIVAL CURVE OF SEVERE RESPIRATORY DISEASE DUE SARS-COV2 IN ≥75 YEARS OLD BY TREATMENT GROUP – mITT ANALYSIS **(Page 22)**

**Figure 7.** CORRELATION BETWEEN COVIDAR, GENTECH EIA AND GENTECH SURROGATE NEUTRALIZATION ANTIBODY ASSAYS **(Page 23)**

**Figure 8.** DISTRIBUTION OF SARS COV2 IGG TITERS >1:1,000 IN PLASMA DONORS **(Page 24)**

**Figure 9.** SERUM TITERS OF IGG AGAINST SARS-COV2 IN INFECTED PLASMA RECIPIENTS 24 HS AFTER INFUSION **(Page 25)**

**Figure 10.** PLASMA COLLECTION **(Page 26)**

**Figure 11.** PATIENT IDENTIFICATION, SCREENING AND TRANSPORTATION TO PARTICIPATING HOSPITALS **(Page 27)**

**Figure 12.** ENROLMENT, FOLLOW UP AND DATA ANALYSIS **(Page 28)**

**Table 1. DEFINITIONS FOR COMORBIDITIES.**

a. Diagnosis of arterial hypertension under pharmacological treatment.

b. Known diagnosis of diabetes in treatment with one or more of the available drugs.

c. Obesity (BMI -Body mass index- ≥ 30 kg / m2).

d. Chronic obstructive pulmonary disease (COPD) treated with any of the available drugs.

e. Cardiovascular disease defined as a) known diagnosis of coronary heart disease, b) history of ischemic stroke or hemorrhagic, or c) ICC (defined as FE < 40%).

f. Chronic kidney disease (defined as a reduction in GFR below 60 ml/min/1.73 $m^2$).

**Table 2. HEART FAILURE.**

**NYHA SCALE (NEW YORK HEART ASSOCIATION)**
**FUNCTIONAL ASSESSMENT OF HEART FAILURE.**

| NYHA FUNCTIONAL CLASSIFICATION | |
|---|---|
| Class I | Without limitation of physical activity. Normal physical exercise does not cause fatigue, palpitations, or dyspnea. |
| Class II | Slight limitation of physical activity. No symptoms at rest. Ordinary activity causes fatigue, palpitations, or dyspnea. |
| Class III | Marked limitation of physical activity. No symptoms at rest. Less than ordinary physical activity causes fatigue, palpitations or dyspnea. |
| Class IV | Inability to carry out any physical activity; symptoms of heart failure are present even at rest and increase with any physical activity. |

**Table 3. CIRRHOSIS (PORTAL BAVENO IV HYPERTENSION CONSENSUS).**

| CIRRHOSIS STAGES | |
|---|---|
| Stage 1 | Absence of esophageal varices and ascites |
| Stage 2 | Esophageal varices without a history of bleeding and without ascites. |
| Stage 3 | Presence of ascites with or without esophageal varices. |
| Stage 4 | Gastrointestinal bleeding due to portal hypertension, with or without ascites. |

**Table 4. KIDNEY DISEASE CLASSIFICATION KDIGO.**

| DEGREE | GLOMERULAR FILTRATION ML / MIN / 1.73 M2 | DESCRIPTION |
|---|---|---|
| GRADE 1 | > 90 G5 | Normal or elevated |
| GRADE 2 | 60-89 | Slightly diminished |
| GRADE 3 | 45-59 | Slight to moderately decreased |
| GRADE 3b | 30-44 | Moderate to severely diminished |
| GRADE 4 | 15-29 | Severely diminished |
| GRADE 5 | <15 | Kidney failure |

**Table 5. EXCLUSION CRITERIA.**

A subject who meets any of the following criteria will be excluded from the study:

1. Present at the time of inclusion SEVERE RESPIRATORY DISEASE defined as (a) respiratory rate ≥30 per minute and / or (b) oxygen saturation in ambient air <93%.
2. HEART FAILURE in functional class III and IV (according to the functional classification of the New York Heart Association).
3. Known diagnosis of CHRONIC KIDNEY INSUFFICIENCY in filtration stages G4 and G5 (according to KDIGO classification).
4. Previous diagnosis of PRIMARY HYPOGAMMAGLOBULINEMIA (congenital and hereditary).
5. Previous diagnosis of MONOCLONAL GAMMOPATHIES (multiple myeloma, Waldenström macroglobulinemia, primary amyloidosis, heavy chain disease, among others).
6. Previous diagnosis of SELECTIVE DEFICIENCY OF IgA.
7. Previous diagnosis of MYELODYSPLASIC SYNDROMES. [WHO classification: refractory thrombocytopenia (TR), refractory anemia (RA), etc.]
8. Previous diagnosis of CHRONIC LYMPHOPROLIFERATIVE SYNDROMES. (WHO classification: peripheral B lymphocyte lymphomas, T lymphocyte and NK cell lymphomas, Hodgkin lymphoma).
9. KNOWN HYPERSENSITIVITY to the administration of immunoglobulins, plasma, monoclonal antibodies and / or vaccines.
10. Active cancer, defined as receiving or having received or receiving chemotherapy, radiotherapy, or new designer or monoclonal drug treatments in the last six months.
11. Known HIV, HBV, or HCV infection.
12. Chronic administration (defined as more than 14 calendar days) of immunosuppressants or other drugs that modify the immune system at the time of enrollment or within the 6 months prior to administration of study medication. An immunosuppressive dose of glucocorticoids will be defined as a systemic dose of ≥10mg of prednisone per day or its equivalent.
13. Solid organ transplant.
14. Known chronic liver disease diagnosed with stage II-III-IV cirrhosis.
15. Chronic lung disease with oxygen requirement.
16. Any other physical, psychiatric or social condition that may, at the discretion of the investigator, increase the risks of participation in the study for the participant or that may lead to the collection of incomplete or inaccurate safety data.
17. Patients who are receiving any type of anticoagulant therapy orally: family of dicoumarins (Warfarin such as Acenocoumarol) or direct acting anticoagulants (DOAC) (Rivaroxaban, Dabigatran, Apixaban) as well as those who are receiving heparin in any of its types for anticoagulant therapy purposes.

**Table 6. Solicited Adverse Events – ITT.**

| | Serious | | | | Non serious | | | |
|---|---|---|---|---|---|---|---|---|
| | Plasma (n=80) | | Placebo (n=80) | | Plasma (n=80) | | Placebo (n=80) | |
| | n/N | % | n/N | % | n/N | % | n/N | % |
| Volume overload | 0/79 | 0.0 | 0/79 | 0.0 | 0/80 | 0.0 | 0/80 | 0.0 |
| Allergic reaction | 0/79 | 0.0 | 0/79 | 0.0 | 0/80 | 0.0 | 0/80 | 0.0 |
| Trombophlebitis | 0/79 | 0.0 | 0/79 | 0.0 | 0/80 | 0.0 | 1/80 | 1.2 |
| Vasovagal syndrome | 0/79 | 0.0 | 0/79 | 0.0 | 0/80 | 0.0 | 0/80 | 0.0 |
| Hematoma at site | 0/79 | 0.0 | 0/79 | 0.0 | 0/80 | 0.0 | 0/80 | 0.0 |
| Nerve injury | 0/79 | 0.0 | 0/79 | 0.0 | 0/80 | 0.0 | 0/80 | 0.0 |
| Tetany (hyperventilation) | 0/79 | 0.0 | 0/79 | 0.0 | 0/80 | 0.0 | 0/80 | 0.0 |
| Total events* | 0 | (0-0) | 0 | (0-0) | 0 | (0-0) | 0 | (0-0) |

**Table 7. Proportion of primary endpoint cases by age group – ITT.**

| | Plasma (n=80) | | Placebo (n=80) | | Relative Risk (IC 95%) | P value |
|---|---|---|---|---|---|---|
| | n/N | % | n/N | % | | |
| **Severe Respiratory Disease** | | | | | | |
| • 65-74 years | 8/40 | 20.0 | 8/32 | 25.0 | 0.80 (0.34; 1.90) | 0.6122 |
| • ≥75 years | 5/40 | 12.5 | 17/48 | 35.4 | 0.35 (0.14; 0.87) | 0.0240 |

**Table 8. Characteristics of the population – mITT.**

| Variables | Plasma (n=76) | | Placebo (n=78) | |
|---|---|---|---|---|
| **Demographic** | **n/N** | **%** | **n/N** | **%** |
| Age in years* | 76.5 | (8.5) | 77.7 | (8.4) |
| Age by categories | | | | |
| • 65-74 years | 37/76 | 48.7 | 32/78 | 41.0 |
| • ≥75 years | 39/76 | 51.3 | 46/78 | 59.0 |
| Sex | | | | |
| • Female | 52/76 | 68.4 | 46/78 | 59.0 |
| • Male | 24/76 | 31.6 | 32/78 | 41.0 |
| **Vital signs** | | | | |
| Axillary temperature* | 36.5 | (0.7) | 36.8 | (0.8) |
| Heart Rate* | 80.0 | (13.3) | 78.7 | (14.1) |
| Systolic blood pressure* | 125.3 | (15.7) | 125.5 | (15.5) |
| Diastolic blood pressure* | 75.4 | (11.2) | 75.0 | (11) |
| Respiratory rate* | 16.9 | (2.4) | 17.3 | (3) |
| Oxygen saturation in room air* | 96.2 | (1.6) | 96.1 | (1.7) |
| Hours from initiation of symptoms* | 39.3 | (14) | 38.6 | (14.3) |
| **Comorbidities** | | | | |
| Hypertension under treatment | 58/76 | 76.3 | 52/78 | 66.7 |
| Diabetes under treatment | 22/76 | 28.9 | 13/77 | 16.9 |
| Obesity | 3/76 | 3.9 | 8/77 | 10.4 |
| COPD under treatment | 1/76 | 1.3 | 5/77 | 6.5 |
| Cardiovascular disease** | 14/76 | 18.4 | 7/77 | 9.1 |
| Chronic renal failure | 1/76 | 1.3 | 3/77 | 3.9 |
| At least one comorbidity | 65/76 | 85.5 | 62/78 | 79.5 |
| **Secondary comorbidities** | | | | |
| Asthma or other respiratory disease | 3/76 | 3.9 | 3/78 | 3.8 |
| Non-cirrhotic liver disease | 0/76 | 0 | 0/78 | 0 |
| Cancer (not active) | 4/76 | 5.3 | 2/78 | 2.6 |
| Neurologic disease | 11/76 | 14.5 | 7/78 | 9 |
| History of smoking | 13/76 | 17.1 | 10/78 | 12.8 |
| Receives medications regularly | 72/76 | 94.7 | 72/78 | 92.3 |
| Received medications in last 15 days | 67/76 | 88.2 | 64/78 | 82.1 |

*mean and standard deviation

**Defined as a known diagnosis of coronary disease, history ischemic or hemorrhagic cardiovascular disease, ICC (defined as EF < 40%)

**Table 9. Primary Endpoint. Proportion of cases – mITT.**

| | Plasma (n=76) | | Placebo (n=78) | | Relative Risk (95%CI) | P value |
|---|---|---|---|---|---|---|
| | n/N | % | n/N | % | | |
| **Severe Respiratory Disease** | 9/76 | 11.8 | 23/78 | 29.5 | 0.40 (0.20; 0.81) | 0.011 |

**Table 10. Primary Endpoint. Days to event - mITT.**

| | Plasma (n=76) | | Placebo (n=78) | | P value* |
|---|---|---|---|---|---|
| | **Median** | **(Q1-Q3)** | **Median** | **(Q1-Q3)** | |
| **Severe Respiratory Disease** | 15 | (15-15) | 15 | (15-15) | 0.0064 |

*Log Rank Test

**Table 11. Secondary Endpoints: Case proportions –ITT.**

| | Plasma (n=76) | | Placebo (n=78) | | Riesgo Relativo (IC 95%) | Valor de P |
|---|---|---|---|---|---|---|
| | n/N | % | n/N | % | | |
| **Life-threatening respiratory disease** | **3/76** | **3.9** | **9/78** | **11.5** | **0.34 (0.10; 1.22)** | **0.0973** |
| • Maximum oxygen support | 3/76 | 3.9 | 5/78 | 6.4 | 0.62 (0.15; 2.49) | 0.4961 |
| • Non-invasive ventilation | 1/76 | 1.3 | 6/78 | 7.7 | 0.17 (0.02; 1.39) | 0.0983 |
| • Intensive care admission | 1/76 | 1.3 | 5/78 | 6.4 | 0.21 (0.02; 1.72) | 0.1439 |
| • Mechanical ventilation | 1/76 | 1.3 | 4/78 | 5.1 | 0.26 (0.03; 2.24) | 0.2189 |
| **Critical systemic illness** | **4/76** | **5.3** | **5/78** | **6.4** | **0.82 (0.23; 2.94)** | **0.7620** |
| • Acute respiratory failure | 1/76 | 1.3 | 4/78 | 5.1 | 0.26 (0.03; 2.24) | 0.2189 |
| • Shock | 1/76 | 1.3 | 1/78 | 1.3 | 1.03 (0.07; 16.11) | 0.9852 |
| • Multiorganic shock syndrome | 3/76 | 3.9 | 4/78 | 5.1 | 0.77 (0.18; 3.33) | 0.7259 |
| **Death due to COVID-19** | **1/76** | **1.3** | **3/78** | **3.8** | **0.34 (0.04; 3.22)** | **0.3482** |
| **Any of three endpoints above** | **6/76** | **7.9** | **11/78** | **14.1** | **0.56 (0.22; 1.44)** | **0.2280** |

**Table 12. Proportion of primary endpoint cases by age group – mITT.**

| | Plasma (n=76) | | Placebo (n=78) | | Relative Risk (95% CI) | P value |
|---|---|---|---|---|---|---|
| | n/N | % | n/N | % | | |
| **Severe Respiratory Disease** | | | | | | |
| • 65-74 years | 5/37 | 13.5 | 8/32 | 25.0 | 0.54 (0.20; 1.49) | 0.2336 |
| • ≥75 years | 4/39 | 10.3 | 15/46 | 32.6 | 0.31 (0.11; 0.87) | 0.0258 |

**Table 13. Cost of a plasma infusion in Buenos Aires, Argentina per patient.**

| PROCEDURE | In AR pesos | In US dollars (160:1) |
|---|---|---|
| Procedure – all cost | 17,000 | 106,25 |
| RT-PCR SARS CoV2 | 4,800 | 30 |
| Outpatient clinic 4 hour stay | 8,000 | 50 |
| TOTAL | 29,800 | 186,25 |

**Suppl. Figure 1. Consort.**

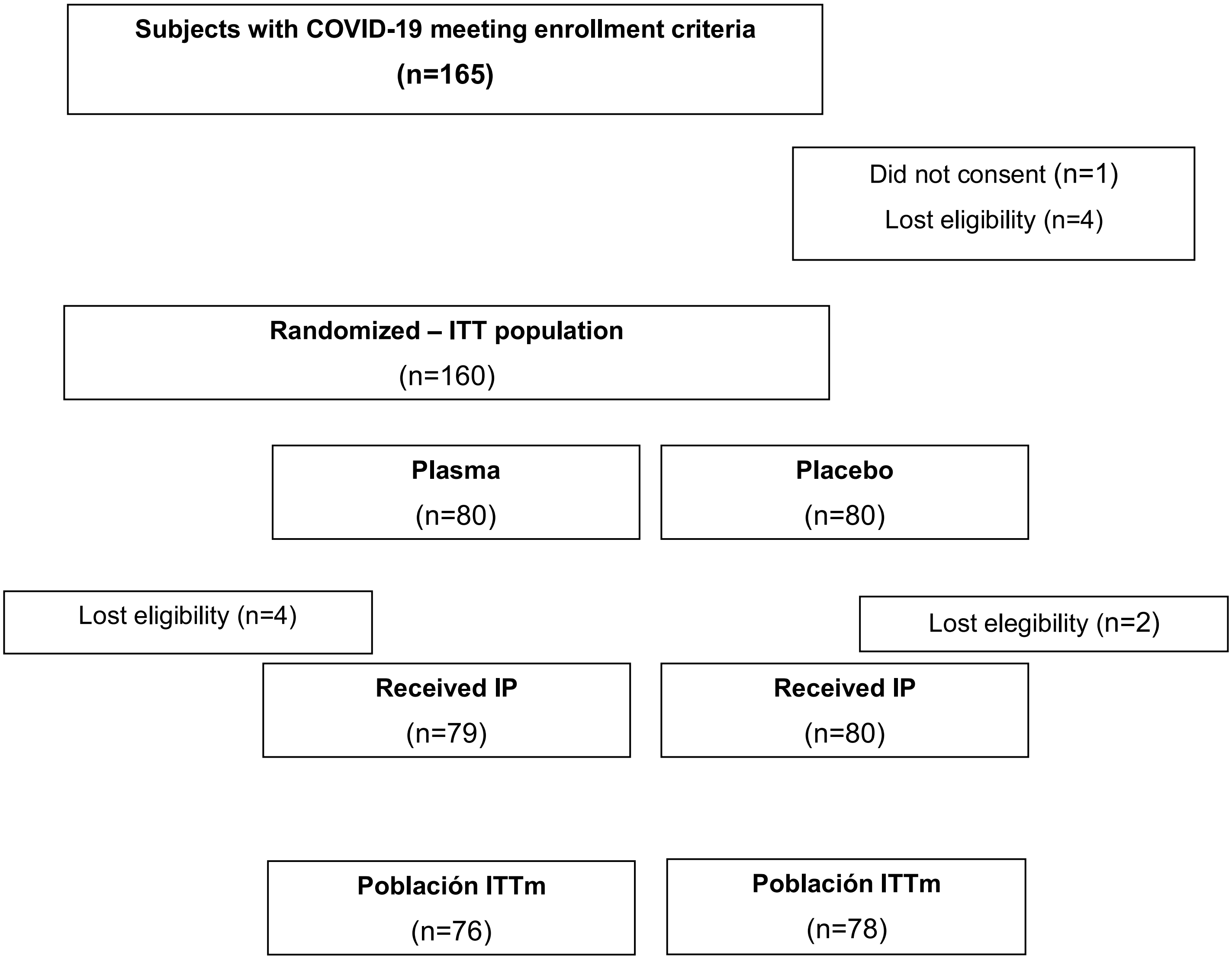

**Suppl. Figure 2. Kaplan-Meier survival curve of severe respiratory disease due SARS-CoV2 in 65-74 years old by treatment group in days – ITT Analysis.**

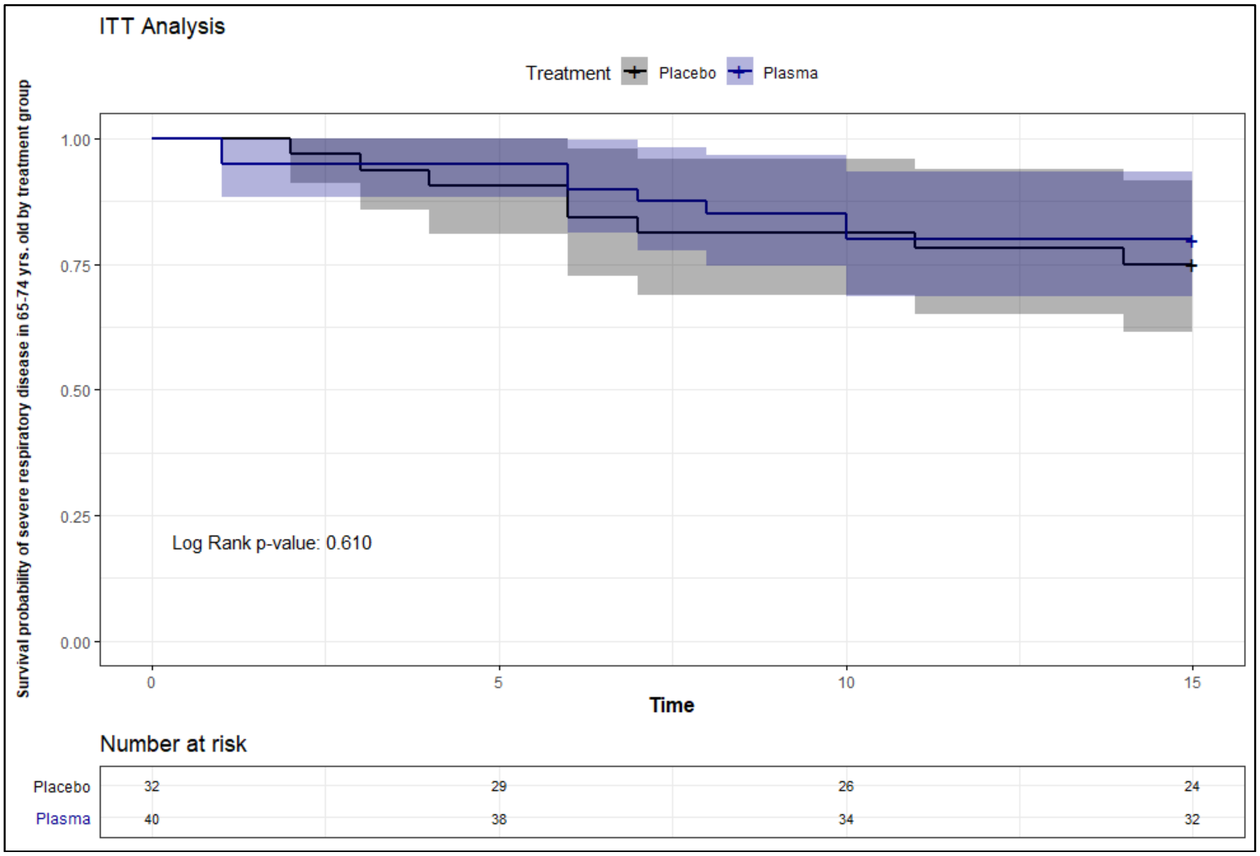


Log Rank test

**Suppl. Figure 3. Kaplan-Meier survival curve of severe respiratory disease due SARS-CoV2 in ≥75 years old by treatment group in days – ITT Analysis.**

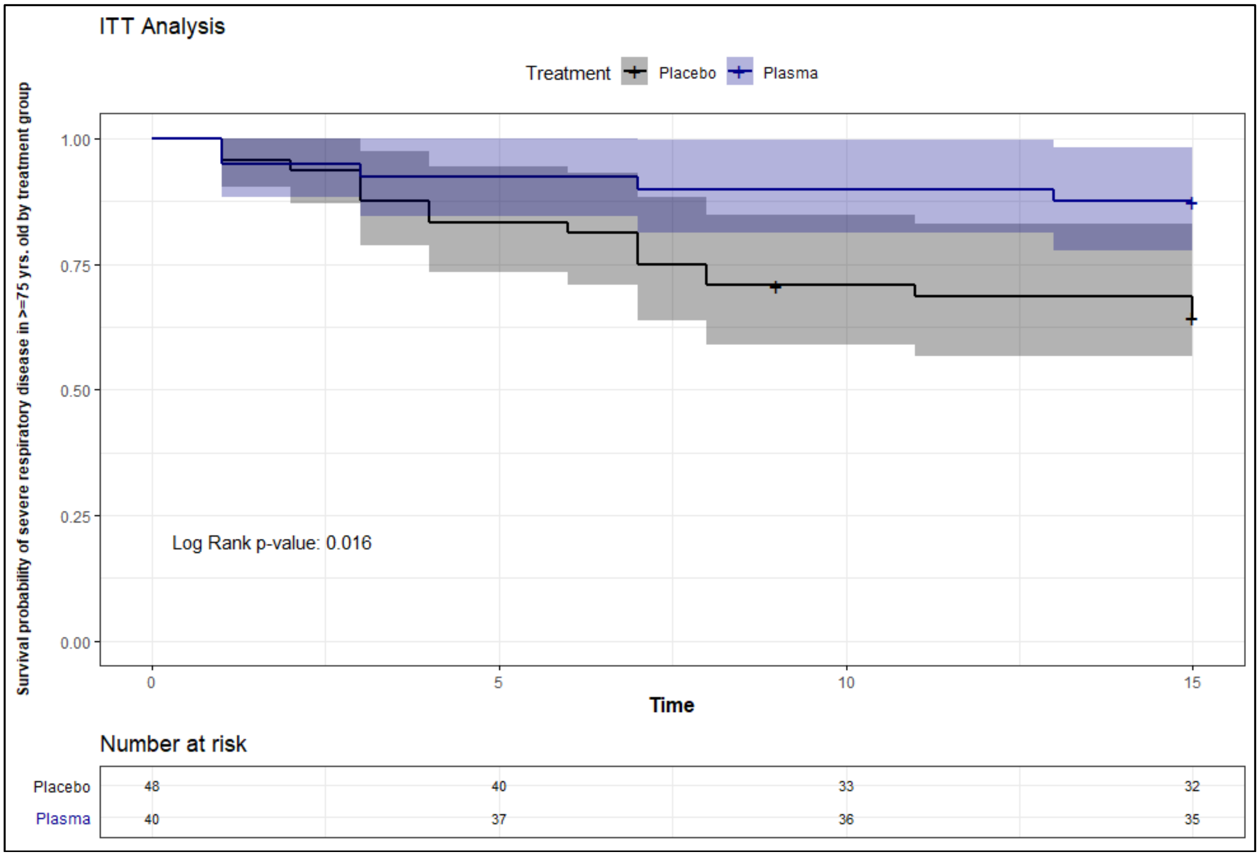


Log Rank test

**Suppl. Figure 4. Kaplan-Meier survival curve of severe respiratory disease (Primary Endpoint) due to SARS-CoV2 by treatment group in days – mITT Analysis.**

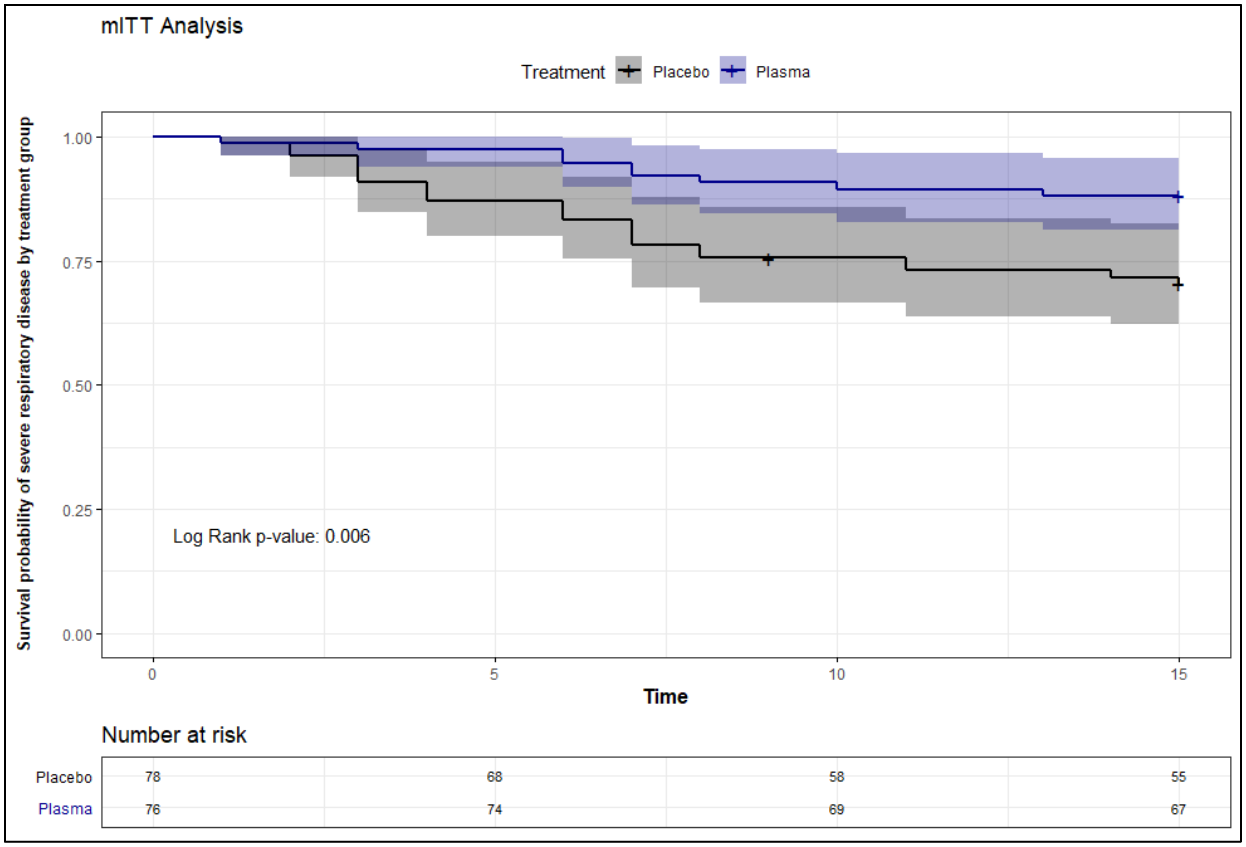


Log Rank test

**Suppl. Figure 5. Kaplan-Meier survival curve of severe respiratory disease due SARS-CoV2 in 65-74 years old by treatment group in days – mITT Analysis.**

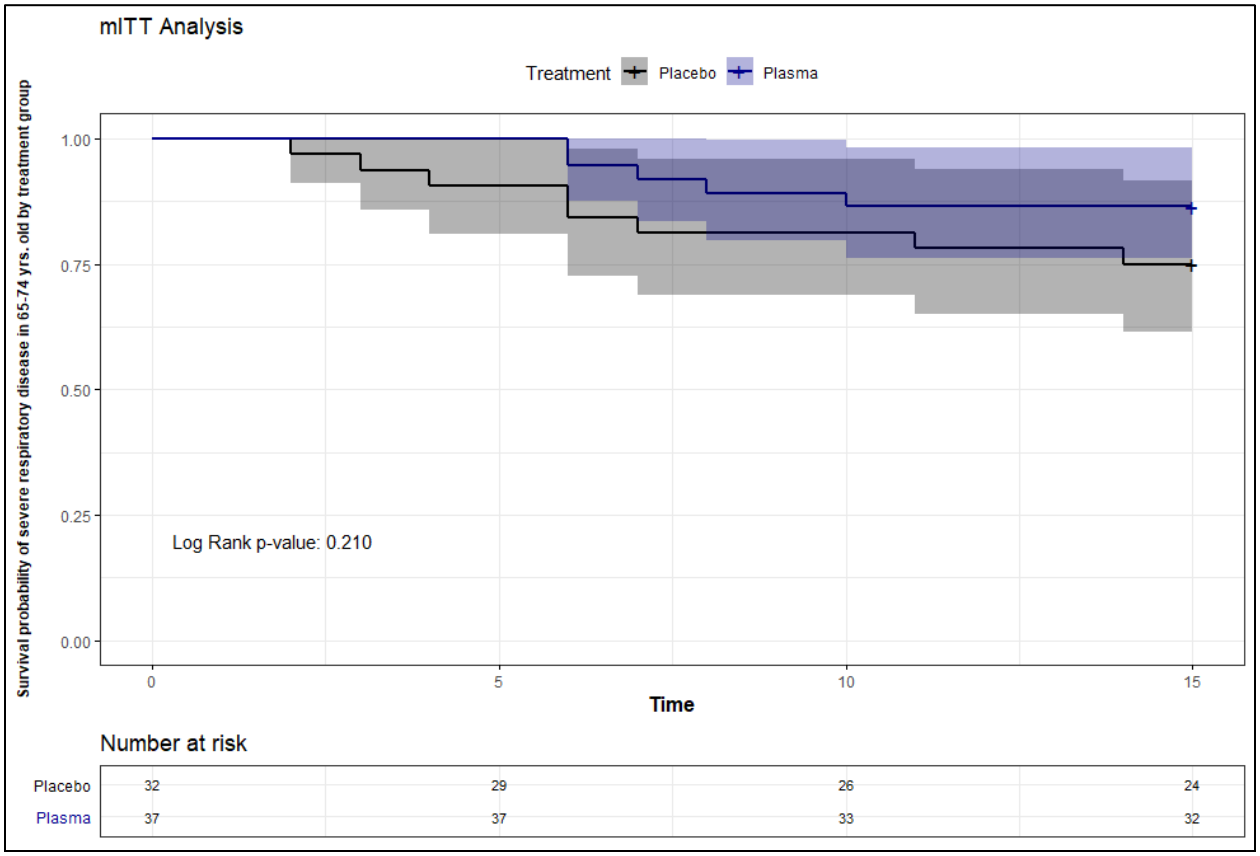


Log Rank test

**Suppl. Figure 6. Kaplan-Meier survival curve of severe respiratory disease due SARS-CoV2 in ≥75 years old by treatment group in days – mITT Analysis.**

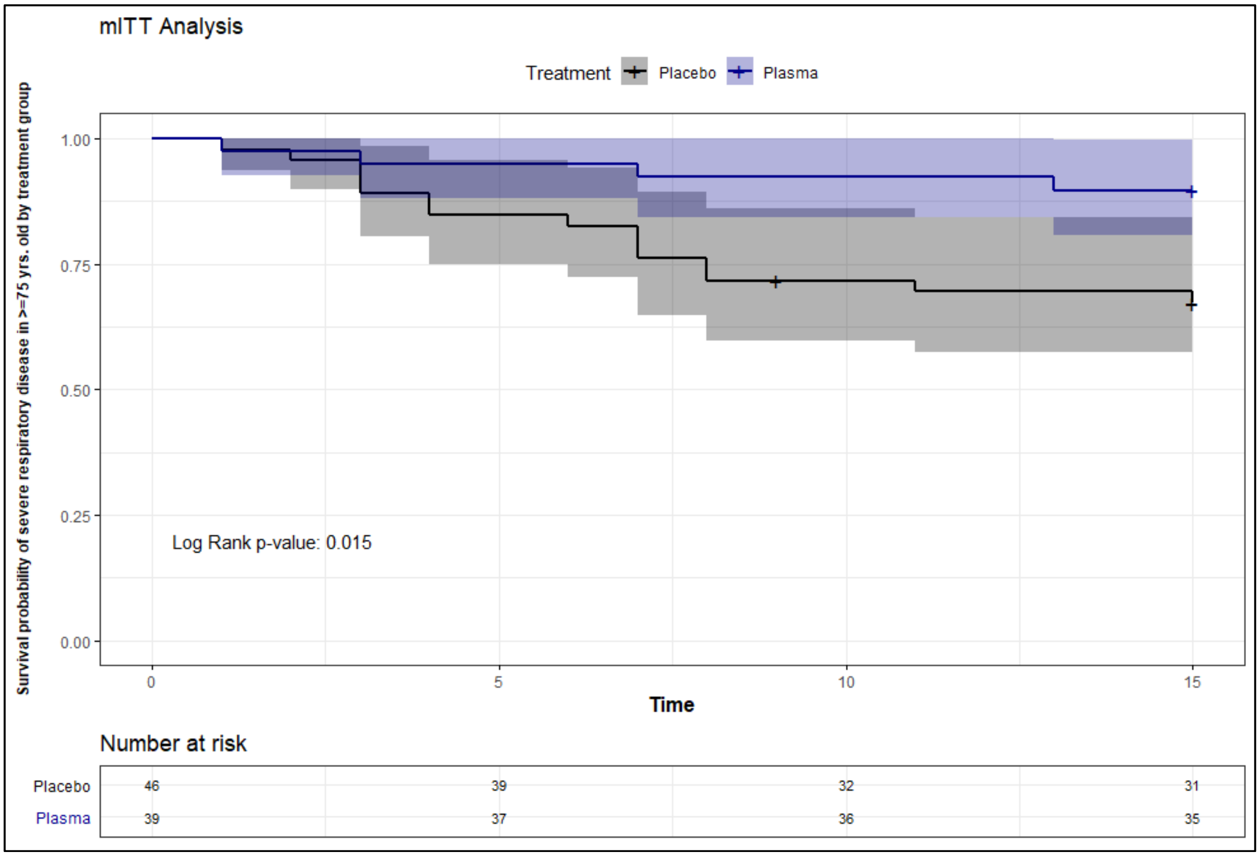


Log Rank test

**Suppl. Figure 7. Correlation between COVIDAR, Genscript EIA and Genscript surrogate neutralization antibody assays.**

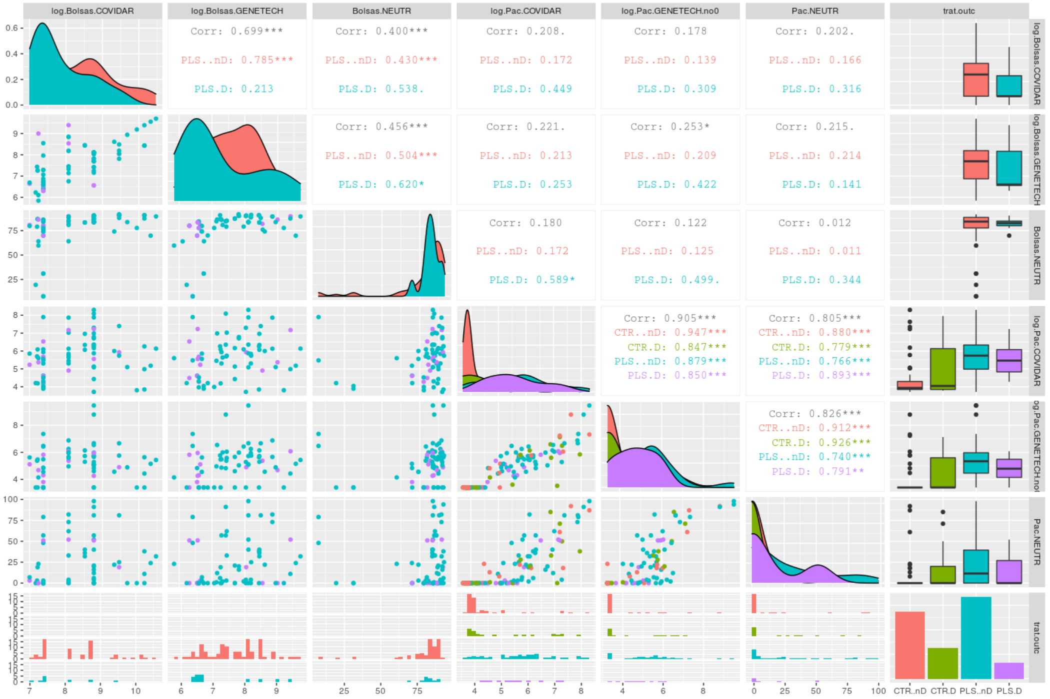

**Suppl. Figure 8. Distribution of SARS CoV2 IgG titers >1:1,000 in plasma donors.**

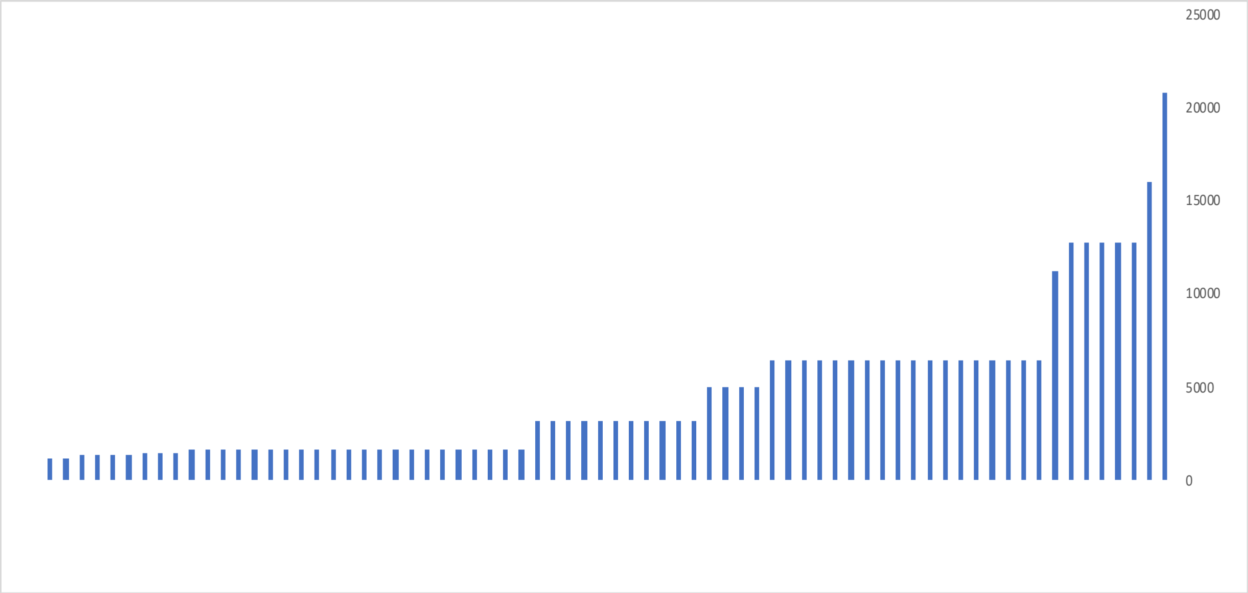

**Suppl. Figure 9. Serum titers of IgG against SARS-CoV2 in infected plasma recipients 24 hs after infusion (violin plot).**

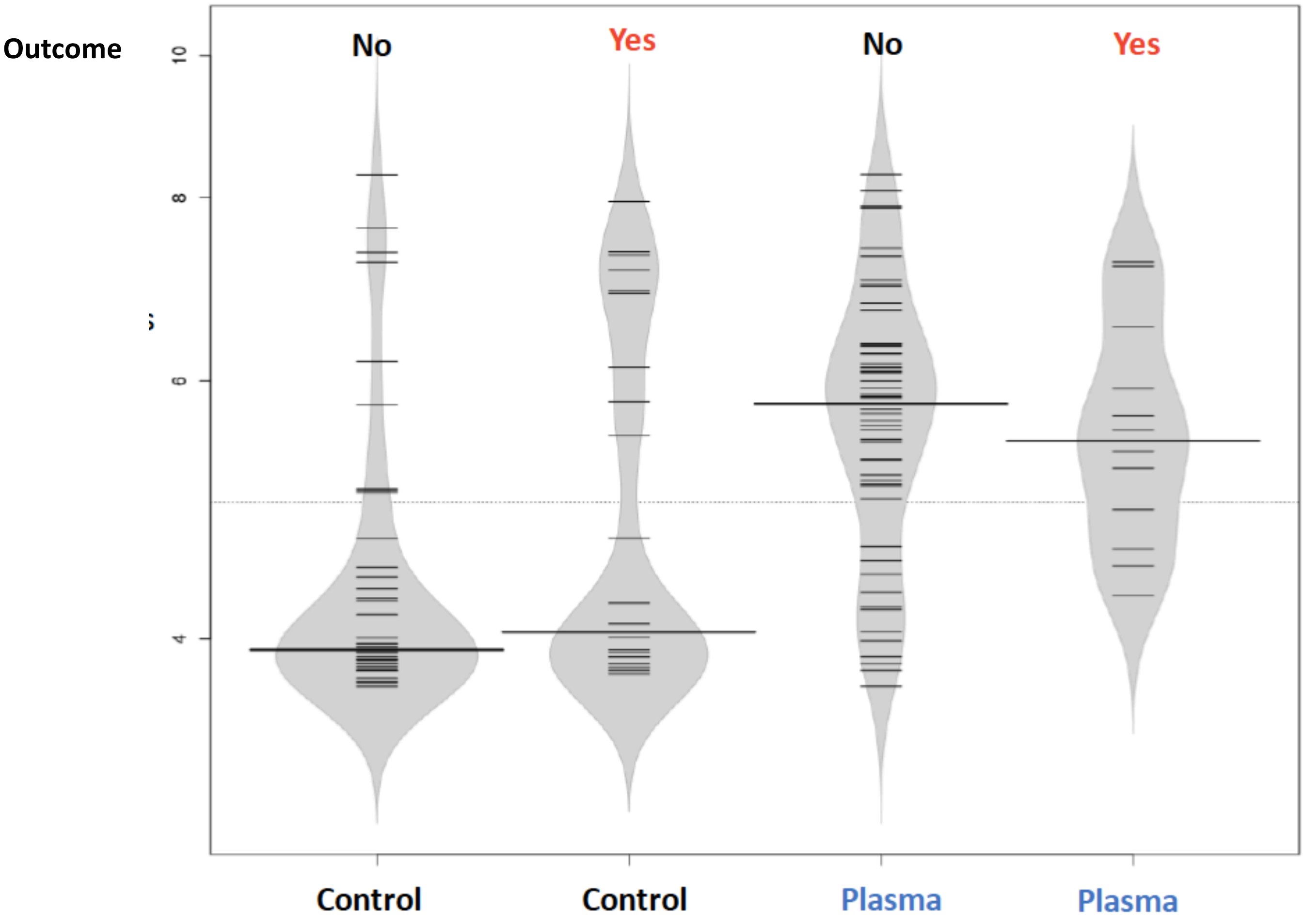

**Suppl. Figure 10. Plasma collection.**

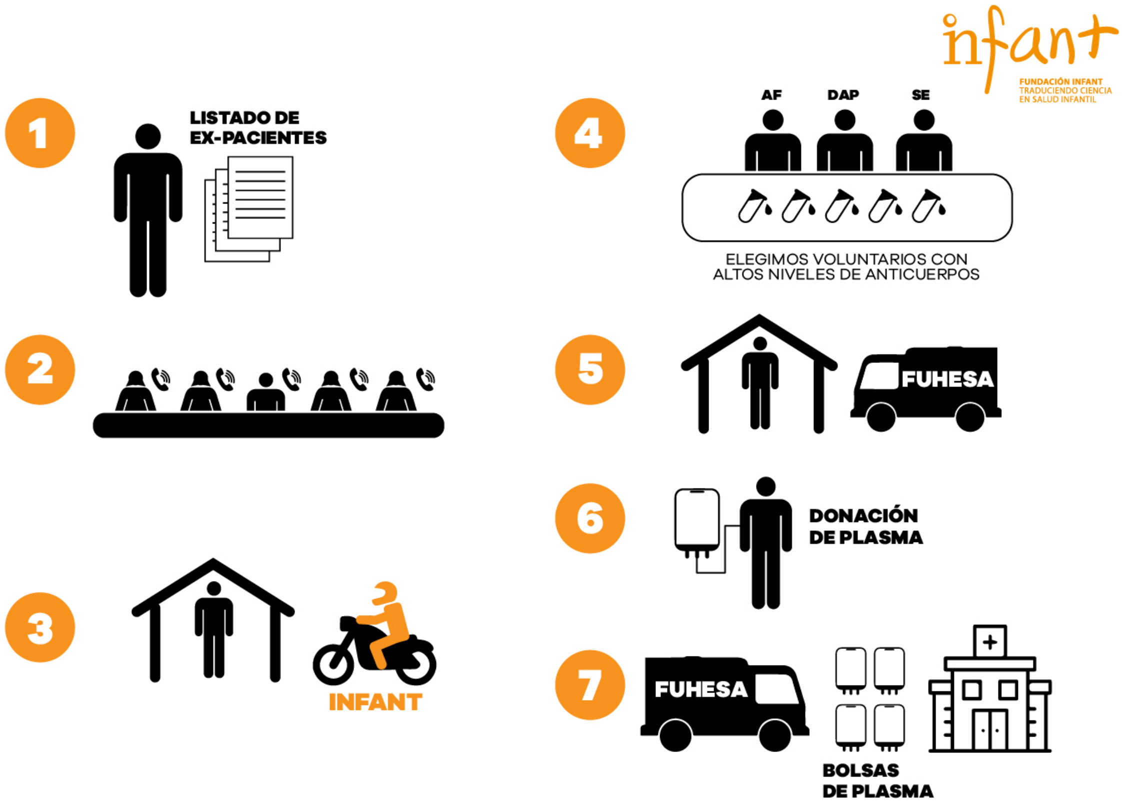

**Suppl. Figure 11. Patient identification, screening and transportation to participating hospitals.**

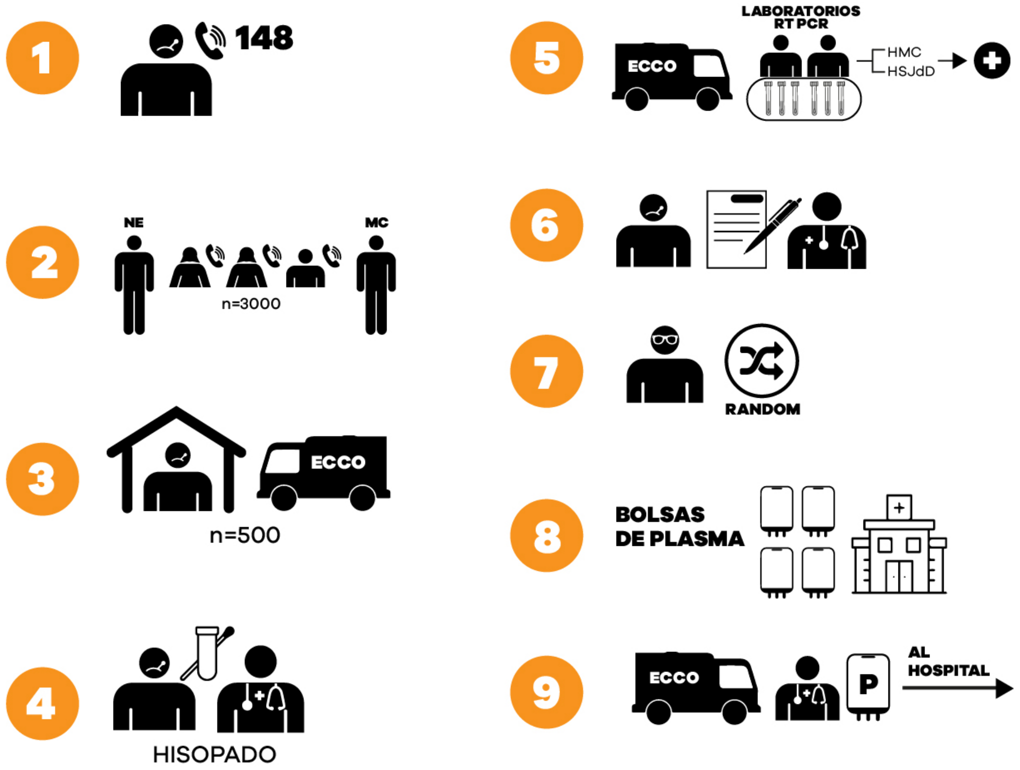

**Suppl. Figure 12. Enrolment, follow up and data analysis.**

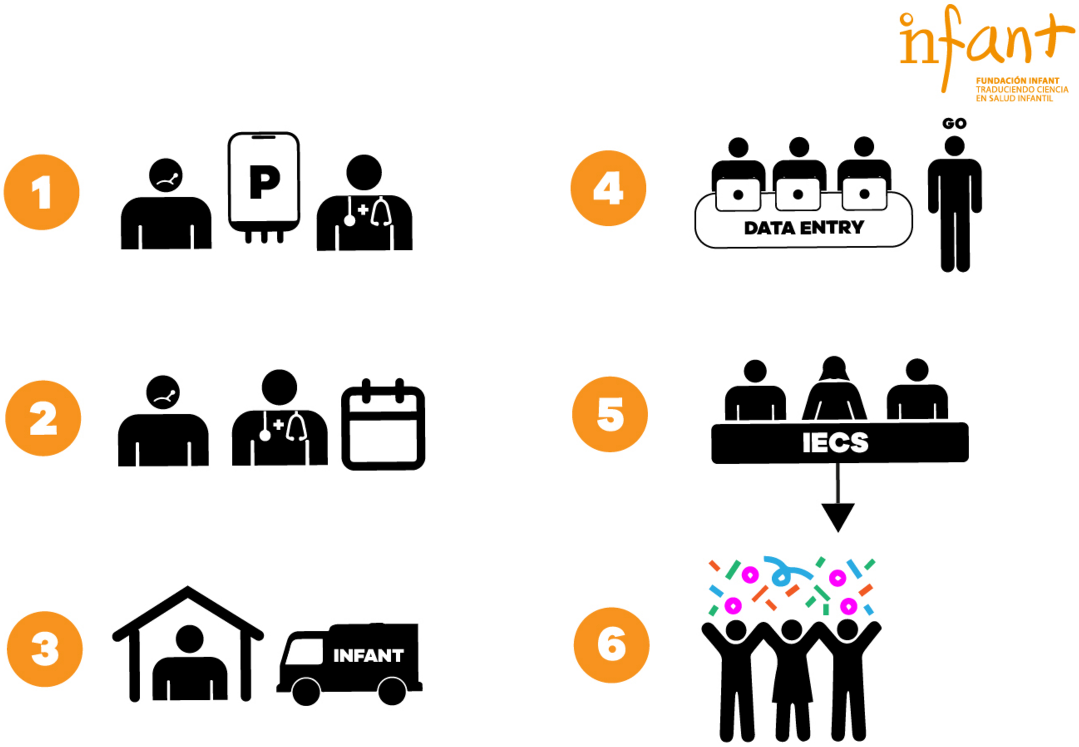
