## Supplementary material for "Prevention of severe COVID-19 in the elderly by early high-titer plasma": Protocolo Original

### EVALUACIÓN DE LA EFICACIA DE LA ADMINISTRACIÓN DE PLASMA DE CONVALECIENTE DE COVID-19 EN LA DISMINUCIÓN DE LA PROGRESIÓN A ENFERMEDAD SEVERA EN ADULTOS MAYORES CON SÍNTOMAS LEVES POR SARS-COV2

**Investigador Principal Hospital Central de San Isidro:** Dr. Ramiro Larrea
**Investigador Principal Hospital Dr. Carlos Bocalandro:** Dr. Aníbal Rondan
**Investigador Principal Hospital Simplemente Evita:** Dra. Valeria Fernández Viña
**Investigador Principal Hospital Evita Pueblo:** Dra. Sandra Azcárate
**Investigador Principal Hospital San Juan de Dios:** Dra. Ivonne Ritou
**Investigador Principal Centros COVID-PAMI**: Dr. Gonzalo Perez Marc
**Investigador Principal del Programa**: Dr. Fernando P. Polack (INFANT)

**Co-Investigadores:** Dra. Romina Libster (INFANT/CONICET), Dr. Gonzalo Pérez Marc (HMC), Dr. Diego Wappner (SMG), Dr. Jorge Lantos (Los Arcos), Dr. Ricardo Valentini (CEMIC), Dr. Federico Etchenique (Ecco), Dr. Maximiliano De Zan (OSECAC), Dr. Gabriel Leberzstein (OSECAC), Dr. Miguel González (Finochietto), Dra. Andrea Gamarnik (Leloir/CONICET), Dr. Jorge Geffner (UBA/CONICET), Bioq. Silvina Coviello (INFANT), Dr. Mauricio Caballero (INFANT/CONICET), Dr. Damián Alvarez Paggi (INFANT/CONICET), Dr. Sebastián Esperante (INFANT/CONICET), Dr. Federico Dimase (HMC), Dra. Susana Pastor Argüello (HMC), Dr. Juan Molinos (Clínica Olivos), Dr. Pablo Cruz (Centro Gallego), Dra. María Dolores Silveyra (Sanatorio Anchorena), Dr. Alfonso Raggio (Sanatorio Antártida), Dr. Juan Sebastián Riera (Ministerio de Salud de la Provincia de Buenos Aires), Dr. Enio García (Ministerio de Salud de la Provincia de Buenos Aires), Dr. Juan Canela (Ministerio de Salud de la Provincia de Buenos Aires), Dr. Mario Rovere (Ministerio de Salud de la Provincia de Buenos Aires), Dr. Fernando Althabe (OMS), Dr. Eduardo Bergel (IECS),

**Asesores:** Dr. Daniel Stamboulian (FUNCEI)

### 1. INTRODUCCIÓN

SARS-CoV2 es un virus nuevo y particularmente agresivo para individuos de edad avanzada, quienes representan entre el 73 y 90% de los casos fatales en distintas regiones del mundo [28, 30, 40]. La mortalidad, la necesidad de cuidados intensivos, la ventilación mecánica, el requerimiento de oxígeno, y las hospitalizaciones por enfermedad respiratoria aumentan marcadamente con el avance de la edad en los pacientes [2, 3, 29, 32, 46]. De hecho, la frecuencia de muerte por enfermedad por SARS-CoV2 (covid-19) es del 8% en los pacientes entre 70 y 80 años, y aumenta a 14,8% en los mayores de 80 [23]. Específicamente en pacientes hospitalizados mayores de 75 años, la mortalidad es de 29,4% [24], y en la terapia intensiva asciende a un alarmante 43,5% [25]. Varias comorbilidades asociadas con enfermedad vascular y/o pulmonar agravan el pronóstico de los infectados aun en pacientes menores de 70, incluyendo hipertensión arterial, diabetes, obesidad y enfermedad pulmonar obstructiva crónica (EPOC) [3,23,30,31]. A la fecha, el virus ha causado más de 290.000 muertes en el mundo, más de 300 de ellas en Argentina donde se encuentra contenido a través de una estrategia transitoria de cuarentena cuasi-universal [4]. No existe aún ningún tratamiento específico disponible. Los resultados de los ensayos de eficacia para vacunas contra SARS-CoV2 se conocerán dentro de mínimamente 10-14 meses.

El SARS-CoV2 es un virus envuelto que contiene ARN monocatenario en sentido positivo unido a una nucleoproteína (proteína N), dentro de una cápside compuesta por proteínas de la matriz (proteína M). La envoltura posee glicoproteínas en forma de espinas (proteína S) que se unen al receptor celular ACE2 en humanos y generan anticuerpos neutralizantes [13]. La primo infección por SARS-CoV2, como otras enfermedades respiratorias, genera una respuesta de anticuerpos con producción temprana de IgM seguida de IgG específica contra las proteínas virales [14]. La detección de anticuerpos neutralizantes del virus es frecuente, y alcanza niveles altos de hasta 1:21,500 $PRNT_{50}$, mayormente excediendo títulos de 1:500 después de 28 días del inicio de síntomas en pacientes leves [41]. Aproximadamente 5% de los pacientes presentan niveles de anticuerpos neutralizantes <1:40, especialmente los más jóvenes [41]. Un estudio de infección primaria vía respiratoria en macacos Rhesus, utilizando $10^6$ pfu de SARS-CoV2 seguido de un segundo desafío intratraqueal usando el mismo virus y dosis 28 días más tarde, demostró ausencia de reinfección tras una extensa evaluación por RT-PCR de tejidos post-necropsia [42].

Los títulos neutralizantes antes del segundo inóculo variaban entre 1:8-1:16 $PRNT_{50}$ [42]. **Estos hallazgos - limitados por cierto, dada la novedad del problema- sugieren que aún bajos títulos de anticuerpos neutralizantes podrían prevenir la enfermedad severa por el virus.**

Proteger a las poblaciones más vulnerables al SARS-CoV2 es imperioso, en el contexto de un germen altamente infeccioso ($R_0$ estimado entre 2 y 3,28) [27, 33, 34]. Su contagiosidad desborda rápidamente la capacidad sanitaria de países desarrollados en distintas regiones del mundo [5] y amenaza los recursos sanitarios de países en vías de desarrollo [6]. Regiones de Italia, España y Estados Unidos se han visto desbordadas por la pandemia, que representa una seria amenaza para Argentina, en coincidencia con el inicio de la circulación anual de otros patógenos pulmonares, como la gripe común y el virus sincicial respiratorio [26]. Evidentemente, resultaría imposible dar respuesta a una avalancha de pacientes graves [7], y toda estrategia tendiente a disminuir las infecciones y el desarrollo de enfermedad severa **se alinea directamente con el objetivo médico de salvar vidas y con las necesidades sanitarias del país para poder brindar la mejor atención posible a aquellos pacientes que lo necesiten**.

La profilaxis post-exposición a virus y el tratamiento en fases tempranas de enfermedad, a través de defensas obtenidas en plasma de convalecientes, es una práctica extendida con antecedentes exitosos en la historia de la medicina a nivel nacional y global. Ya en 1960, Brunell y col. lograron prevenir la varicela clínica en hermanos con padres enfermos, usando inmunoglobulinas anti-varicela zoster obtenidas del plasma de pacientes previamente infectados [8]. Hoy mismo, por ejemplo, la inmunoglobulina enriquecida para sarampión se utiliza en personas no vacunadas que estuvieron en contacto con un enfermo durante los 6 días previos con una eficacia cercana al 100% [9, 52]; un producto similar enriquecido para el virus de varicela zoster se administra en embarazadas, recién nacidos, e inmunosuprimidos no vacunados en contacto con infectados dentro de las 96 horas disminuyendo un 90% la incidencia de enfermedad grave [10, 53, 54]; la inmunoglobulina antitetánica suplementa la vacunación en víctimas de heridas sucias con inmunización primaria incompleta [43]; la gammaglobulina contra hepatitis B se utiliza como prevención para recién nacidos de madres infectadas con una eficacia del 75% [44, 55]; y la gammaglobulina antirrábica es ~100% efectiva en la profilaxis posterior a la agresión de un animal sospechoso [11,56]. **Las intervenciones utilizando inmunidad humana derivada del plasma de convalecientes han demostrado ser seguras, han prevenido innumerables casos de enfermedad severa, y han preservado vidas en todo el mundo.**

El objetivo principal de nuestro estudio es evaluar la eficacia del plasma de convaleciente en reducir la progresión a enfermedad severa en personas entre 65-74 años con al menos una comorbilidad y en todos los ≥ 75 años que se presenten con sintomatología leve de menos de 48 hs de evolución al momento del inicio de la pesquisa y reciban diagnóstico temprano de COVID-19. *Nuestra hipótesis central es que una dosis única de plasma de convaleciente comparada contra placebo (solución salina al 0,9%), y administrada hasta 72 horas después del inicio de los síntomas por SARS-Cov2 (o sea, hasta 24 horas después del límite de tiempo para presentarse a pesquisa y testeo), prevendrá la progresión a una enfermedad respiratoria severa en sujetos entre 65-74 años con al menos una comorbilidad y en los ≥ 75 años independientemente de presencia de comorbilidad basal.*

#### 2. OBJETIVOS DEL ESTUDIO

##### 2.1. Objetivo Primario

Evaluar la eficacia del plasma de convaleciente, a partir de 12 horas de la administración, en reducir la progresión a enfermedad respiratoria severa en personas entre 65-74 años con al menos una comorbilidad y en todos los ≥ de 75 años con sintomatología leve y diagnóstico temprano de COVID-19 hasta los 15 días de administración del tratamiento (16-18 días de enfermedad considerando el periodo sintomático pre-enrolamiento),

##### 2.2. Objetivos Secundarios

Los Objetivos Secundarios incluyen determinar si la administración de plasma de convaleciente hasta los 15 días o a partir de entonces hasta un máximo de *25 días de la administración de tratamiento si continuase internado* consigue:

- Disminuir la necesidad de soporte de oxígeno con oxigenoterapia máxima (mascara de reservorio de O2) y/o utilización de soporte respiratorio no invasivo (soporte de VNI incluyendo CPAP) y/o admisión a UTI y/o requerimiento de ventilación mecánica invasiva por SARS-CoV2 en pacientes entre 65-74 años con al menos una comorbilidad y en todos los ≥ 75 años
- Disminuir la enfermedad crítica definida como (a) presencia de falla respiratoria aguda (PaO2/FiO2 ≤ 200 mm Hg.) y/o (b) shock (definido por necesidades de soporte con drogas vasoactivas para mantener P.A.M. igual o mayor a 65 mm. Hg), y/o (c) síndrome de disfunción multiorgánica (SDMO): injuria renal aguda (definida por aumento de creatinina por dos o más veces con respecto al basal o aumento de creatinina 0.3 mg/dl), elevación de enzimas hepáticas

(transaminasas) mayor a tres veces el valor superior normal, miocardiopatía aguda (definida por elevación de troponina por encima del nivel normal y/o nuevas alteraciones electrocardiográficas o ecocardiográficas para daño miocárdico) por SARS-Cov2 en pacientes entre 65-74 años con al menos una comorbilidad y en todos los ≥ 75 años a causa de COVID-19.

- Disminuir la mortalidad por SARS-CoV2 en pacientes entre 65-74 años con al menos una comorbilidad y en todos los ≥ 75 años a causa de COVID-19.
- Disminuir la duración de soporte de oxigeno debido a covid-19 en pacientes hipoxémicos.
- Describir la seguridad de la administración de plasma de convaleciente en pacientes entre 65-74 años con al menos una comorbilidad y en todos los ≥ 75 años con COVID-19.
- Explorar la concentración de IgG anti-S de SARS CoV2 en plasma de los participantes asociada con ausencia de enfermedad respiratoria severa obtenidas a las 24 horas de la infusión.

##### 2.3. Objetivos Exploratorios

- Explorar si, en lugar de proteger contra la enfermedad respiratoria severa, la administración de plasma de convaleciente retarda la aparición de dicha enfermedad.
- Explorar los efectos del plasma de convaleciente sobre la respuesta inmune humoral primaria contra la infección por SARS CoV2. a los 90 días de enrolamiento
- Explorar si los efectos en enfermedad respiratoria, cardiovascular y mortalidad a largo plazo fueron afectados por la intervención a los 12 meses de enrolamiento.
- Explorar la correlación entre carga viral determinada por RT-PCR al diagnóstico y la respuesta a tratamiento con plasma.

#### 3. CRITERIOS DE VALORACIÓN DE EFICACIA

##### 3.1.1. Criterio Primario de Valoración de Eficacia

Enfermedad respiratoria severa por SARS-CoV2, confirmada por detección de ARN viral por RT-PCR, y definida por la presencia de cualquiera de las dos siguientes variables en forma no excluyente: (a) frecuencia respiratoria ≥30 por minuto, (b) saturación de oxígeno en aire ambiental <93% [20,21]. El endpoint primario se determinará a partir de 12 horas del inicio de la infusión hasta 15 días desde la administración del tratamiento.

###### 3.1.2. Criterios Secundarios de Valoración de Eficacia *(hasta los 15 días o, a partir de entonces, hasta un máximo de 25 días de la administración de tratamiento si continuase internado)*

- Necesidad de soporte de oxígeno con oxigenoterapia máxima (mascara de reservorio de O2) y/o soporte respiratorio no invasivo (soporte de VNI incluyendo CPAP) y/o admisión a UTI y/o requerimiento de ventilación mecánica invasiva, por SARS-Cov2.
- Enfermedad crítica definida como (a) presencia de falla respiratoria aguda ($PaO2/FiO_2$ ≤ 200 mmHg.) y/o (b) shock, y/o (c) síndrome de disfunción multiorgánica (SDMO) según criterios detallados en sección 2.2: injuria renal aguda, elevación de enzimas hepáticas, miocardiopatía aguda, por SARS-Cov2.
- Mortalidad por covid-19.
- Combinación de los criterios secundarios de valoración de eficacia #1 (necesidad de soporte de oxigeno con…) y/o #2 (enfermedad critica definida…) y/o #3 (mortalidad por covid-19).
- Duración del requerimiento de soporte de oxigeno en pacientes con covid-19 debido a saturación en aire ambiental <93%.

#### 4. HIPÓTESIS Y JUSTIFICACIÓN

##### 4.1. Hipótesis

Nuestra hipótesis central es que una dosis de plasma de convaleciente comparada contra placebo, y administrada hasta 72 horas desde el inicio de los síntomas leves, prevendrá la progresión a enfermedad respiratoria severa por covid-19 en pacientes entre 65-74 años con al menos una comorbilidad y en todos los ≥ 75 años que se presenten con menos de 48 horas de evolución al proceso de pesquisa y con sintomatología leve.

#### 4.2. Justificación

La eficacia de una preparación terapéutica contra el SARS-CoV2 económicamente accesible analizada mediante un estudio aleatorizado, doble ciego, en pacientes vulnerables permitiría escalar de inmediato su aplicabilidad a nivel nacional e internacional mediante campañas masivas de donación de plasma de convalecientes a la espera de soluciones definitivas tales como medicamentos exitosos, anticuerpos monoclonales, o vacunas [17]. Inclusive, de existir protección parcial concentrada en el tracto respiratorio inferior [18], *esta misma estrategia podría hipotéticamente permitir la inmunización de los sujetos mediante la replicación controlada del germen en el tracto respiratorio alto* (como en el caso de la administración pasiva de anticuerpos contra el virus sincicial respiratorio en lactantes [19]).

### 5. DISEÑO DEL ESTUDIO

Un estudio aleatorizado, doble-ciego, controlado con placebo para testear la eficacia del plasma de convaleciente, administrado hasta 72 horas después del inicio de síntomas, con el fin de evitar la progresión hacia una enfermedad respiratoria severa. El estudio se llevará a cabo en sujetos entre 65-74 años con al menos una comorbilidad y en todos los ≥ 75 años que se presenten con sintomatología leve de menos de 48 horas de evolución al momento de la pesquisa (definida en sección 5.1) y diagnóstico precoz de covid-19. El estudio se realizará durante un período de observación mínimo de 15 días en el que los participantes permanecerán bajo control médico diario por protocolo para determinar el criterio de valoración primario. El período de monitoreo se extenderá a partir de entonces, hasta un máximo de 25 días de la administración de tratamiento, si continuase internado, para analizar los criterios de valoración de eficacia secundarios.

Se proyecta que el estudio incorpore un máximo estimado de 210 pacientes (ver 9.0 Tamaño muestral). Los participantes serán aleatorizados en una proporción de 1:1.

**Tabla 1. Asignaciones de Tratamiento**

| Grupo de Tratamiento | Rango de sujetos asignados por rama | Artículo de Prueba | Volumen de Dosificación intravenoso | Día de Administración de la intervención |
|---|---|---|---|---|
| A | 105 | Placebo (solución salina al 0,9%) | 250 cc | Día 0 |
| B | 105 | Plasma de convaleciente | | |

Todos los participantes recibirán una dosis única endovenosa (EV) - acorde a las NORMAS ADMINISTRATIVAS Y TÉCNICAS RM 797/13 – 139/14 – 1507/15 Dirección de Sangre y Hemoderivados Ministerio de Salud de la Nación (por http://www.msal.gob.ar/disahe/images/stories/pdf/normas-hemoterapia.pdf) el Día 0 con el artículo de prueba asignado, plasma de convaleciente o placebo (ver Tabla 1). Para cada sujeto, la participación en el estudio se extenderá un mínimo de 15 días o hasta la resolución de síntomas desde la administración del artículo de prueba con un máximo de 25 días. Los sujetos participantes serán monitoreados durante y después de la administración del artículo de prueba para evaluar eventos de seguridad esperables y no esperables luego de la administración de plasma. Un Comité de Monitoreo de Datos y Seguridad supervisará el enrolamiento, la eficacia y la seguridad de los participantes a lo largo del estudio.

El estudio se llevará a cabo en el Hospital San Juan de Dios, el Hospital Simplemente Evita, el Hospital Dr. Carlos Bocalandro, el Hospital Evita Pueblo, el Hospital Central de San Isidro y el Sanatorio Antártida de la Provincia de Buenos Aires en estrecha coordinación con los efectores sanitarios del Ministerio de Salud provincial y en el Hospital Militar Central, el Sanatorio de Los Arcos, CEMIC, el Centro de Investigación OSECAC, el Sanatorio Finochietto, el Centro Gallego de Buenos Aires, el Sanatorio Anchorena y la Clínica Olivos garantizando que no se interferirá con los cuidados y las normas de atención de pacientes dispuestas para la notificación y manejo de esta enfermedad.

**EQUIPO MOVIL-COVID.** Dado el contexto de la situación covid-19 en el AMBA con la reducción de disponibilidad de camas y la sobrecarga del personal hospitalario sumado al aumento exponencial de los casos graves, los pacientes levemente sintomáticos pero sin requerimientos de oxígeno y provenientes de geriátricos encuentran extraordinariamente difícil (casi imposible) su internación en los hospitales. Por tanto, para facilitar la incorporación de los principales beneficiarios potenciales de esta intervención al estudio (es decir, los ancianos mas vulnerables) y dada la enorme necesidad de salud publica existente para proveer alternativas terapéuticas contra este flagelo, se agregara una unidad COVID-Móvil que monitoreará participantes voluntarios en Centros COVID-PAMI donde la administración del producto de investigación (hoy demostrado claramente como seguro en publicaciones de más de 20.000 pacientes) se realizará bajo las mismas normas y por el mismo equipo que en todos los otros sitios de investigación y el equipo COVID-Móvil registrara la evolución de dichos participantes que se encuentran bajo el cuidado de médicos y personal de salud del PAMI *in situ*. Todo traslado de estos pacientes a centros de mayor complejidad por la evolución natural de su enfermedad será decisión de los médicos tratantes del PAMI, y el equipo de investigación móvil se limitará a continuar recabando datos de su progreso clínico de la misma forma que lo hace hoy cuando un paciente es trasladado o recibe un alta temprana en otra institución.

#### 5.1. Criterios de elegibilidad

##### 5.1.1 Criterios de Inclusión:

Un sujeto debe cumplir los siguientes criterios para ser incluido en el estudio:

1. Edad ≥ 75 años o edad entre 65-74 años con al menos una de las comorbilidades siguientes:
    a. Diagnóstico de hipertensión arterial bajo tratamiento farmacológico.
    b. Diagnóstico conocido de diabetes en tratamiento con algunas de las drogas disponibles (Apéndice I).
    c. Obesidad (IMC -Índice de masa corporal- ≥ a 30 kg/m2),
    d. Diagnóstico de enfermedad pulmonar obstructiva crónica (EPOC) en tratamiento con algunas de las drogas disponibles (Apéndice I).
    e. Enfermedad cardiovascular establecida definida como a) diagnóstico conocido de enfermedad coronaria, b) historia de enfermedad cerebrovascular isquémica o hemorrágica, o c) ICC (definida como FE < 40%).
    f. Enfermedad renal crónica (definida como reducción del FG por debajo de 60 ml/min/1,73 $m^2$).
2. Presentar hace menos de 48 horas: (A)Temperatura axilar ≥37,5°C o equivalente febril (definido por sensación de frío y/o escalofríos y/o sudoración inexplicada), combinado con (a) tos seca y/o (b) dificultad respiratoria y/o (c) odinofagia y/o (d) anosmia/disgeusia y/o (e) alguno de los siguientes síntomas: fatiga, anorexia, mialgias o rinorrea.
3. Diagnóstico de SARS-Cov2 confirmado por RT-PCR
4. Ser capaz, según juicio médico, de comprender y cumplir con los procedimientos del estudio y de consentir o no su participación.
5. Proveer consentimiento informado por sí mismo.
6. Pacientes con vulnerabilidad cognitiva con apoyo en la toma de decisión por su representante según corresponda. (ver sección 12)

##### 5.1.2 Criterios de Exclusión

Un sujeto que cumple cualquiera de los siguientes criterios será excluido del estudio:

1. Que presente al momento del ingreso ENFERMEDAD RESPIRATORIA SEVERA definida como (a) frecuencia respiratoria ≥30 por minuto y/o (b) saturación de oxígeno en aire ambiental <93%.
2. INSUFICIENCIA CARDIACA en clase funcional III y IV (según clasificación funcional de la New York Heart Association).
3. Diagnóstico conocido de INSUFICIENCIA RENAL CRÓNICA en estadios de filtrado G4 y G5 (según clasificación KDIGO).
4. Diagnóstico previo de HIPOGAMMAGLOBULINEMIAS PRIMARIAS (congénitas y hereditarias).
5. Diagnóstico previo de GAMMAPATÍAS MONOCLONALES (mieloma múltiple, macroglobulinemia de Waldenström, amiloidosis primaria, enfermedad de las cadenas pesadas, entre otras).
6. Diagnóstico previo de DEFICIENCIA SELECTIVA DE IgA.
7. Diagnóstico previo de SÍNDROMES MIELODISPLÁSICOS. [clasificación OMS: trombocitopenia refractaria (TR), anemia refractaria (AR), etc.]
8. Diagnóstico previo de SÍNDROMES LINFOPROLIFERATIVOS CRÓNICOS. (clasificación OMS: linfomas periféricos de linfocitos B, linfomas de linfocitos T y células NK, Linfoma de Hodgkin).
9. HIPERSENSIBILIDAD CONOCIDA a la administración de inmunoglobulinas, plasma, anticuerpos monoclonales y/o vacunas.
10. Cáncer activo, definido por estar recibiendo o haber recibido o estar recibiendo quimioterapia, radioterapia o tratamientos con drogas nuevas de diseño o monoclonales en los últimos seis meses.
11. Infección conocida por HIV, HBV o HCV.
12. Administración crónica (definida como más de 14 días corridos) de inmunosupresores u otras drogas que modifican el sistema inmune **al momento del enrolamiento** o dentro de los 6 meses previos a la administración de la medicación del estudio. Una dosis inmunosupresora de glucocorticoides se definirá como una dosis sistémica ≥10mg de prednisona por día o su equivalente.
13. Antecedente de transplante de órgano sólido.
14. Enfermedad hepática crónica conocida con diagnóstico de cirrosis estadio II-III-IV.
15. Enfermedad pulmonar crónica con requerimiento de oxígeno.
16. Cualquier otra condición física, psiquiátrica o social que pueda, a criterio del investigador, aumentar los riesgos por la participación en el estudio a los participantes o que puedan conducir a la recolección de datos de seguridad incompletos o inexactos.
17. Pacientes que estén recibiendo cualquier tipo de terapia anticoagulante sea por vía oral; familia de los dicumarínicos (Warfarina como Acenocumarol) o anticoagulantes de acción directa (ACOD) (Rivaroxaban, Dabigatran, Apixaban)

así como tampoco aquellos que estén recibiendo heparina en cualquiera de sus tipos con fines de terapia anticoagulante.

##### 5.2. Procedimientos del estudio (Tabla 2)

##### 5.2.a. Población participante y proceso de identificación de potenciales sujetos.

Sujetos entre 65-74 años con al menos una comorbilidad y todos los ≥ 75 años que presenten hace menos de 48 hs alguno de los siguientes síntomas: (**a**) temperatura ≥37,5°C o equivalente febril (definido por sensación de frío o escalofríos o sudoración inexplicada), combinado con (**b**) tos seca y/o disnea y/o anosmia/disgeusia y/o odinofagia y/o fatiga y/o anorexia y/o mialgias y/o rinorrea.

Dichos candidatos serán invitados a contactarse de inmediato con el fin de evaluar su participación a través de los números designados por las organizaciones intervinientes.

Aquellos pacientes que cumplan con los criterios de inclusión y no cumplan ninguno de los criterios de exclusión arriba mencionados serán invitados a participar en la fase de pesquisa del estudio en el cual, en caso de aceptar (plasmando su conformidad a través de la firma del consentimiento informado de pesquisa - ANEXO), se obtendrá una muestra de secreción respiratoria por medio de hisopados nasofaríngeo y orofaríngeo (de acuerdo al procedimiento y normativa implementada por el Ministerio de Salud) para detectar ARN de SARS-CoV2 por la prueba de cadena de la polimerasa (RT-PCR).

Dicha muestra será procesada y analizada por los laboratorios intervinientes designados para las instituciones participantes, y que actualmente proveen este mismo servicio diagnostico diariamente, utilizando el mismo test de RT-PCR para evaluar la presencia de ARN de SARS-CoV2. Esta determinación se realizará lo antes posible (en un máximo de 24 horas) y, tal como hasta hoy, bajo todas las normas de bioseguridad indicadas por las autoridades sanitarias. Las muestras se conservarán para posterior testeo por RT-PCR de co-infecciones con influenza A y B, virus sincicial respiratorio y otros patógenos virales de invierno en la Fundación INFANT.

Aquellos pacientes que ya cuenten con un resultado positivo (+) y considerado válido podrán ser invitados a participar de la fase de pesquisa del estudio y no serán hisopados nuevamente.

##### 5.2.b. Enrolamiento para la participación en el estudio

En caso de detectarse en la muestra respiratoria ARN de SARS-CoV2, se informará al candidato sobre los detalles del estudio y se verificará su elegibilidad nuevamente. El candidato podrá decidir no participar de la investigación según su libre albedrío y sin consecuencia alguna de ningún tipo. En caso de que el sujeto esté interesado en participar en el estudio, y tras su consentimiento atestado con la firma del consentimiento informado de participación (ANEXO), se procederá a asignarle un número de identificación único. Una vez asignado su código alfanumérico, se procederá a obtener los signos vitales y realizar un examen físico completo para obtener la siguiente información demográfica y clínica [1-3, 25, 47-51]:

- Sexo
- Edad
- Medicaciones habituales y de los últimos 15 días.
- Antecedentes de tabaquismo
- Comorbilidades de interés primario: hipertensión arterial, diabetes, enfermedad coronaria, insuficiencia cardíaca, antecedentes de ACV e IAM, EPOC, nefropatía crónica, y obesidad.
- Comorbilidades secundarias: asma, antecedentes de cáncer, enfermedades hepáticas, y enfermedades neurológicas.

##### 5.2.c. Asignación aleatoria

Todos los participantes con elegibilidad confirmada serán asignados aleatoriamente a un grupo de tratamiento. La asignación aleatoria de sujetos participantes se realizará utilizando un sistema electrónico por el encargado de aleatorización del estudio. Una vez aleatorizado el paciente, el encargado de la asignación se comunicará con un miembro del equipo de Medicina Transfusional Central del estudio quien procederá a extraer la bolsa opaca de tratamiento indicada del freezer del Laboratorio Central. El equipo de Medicina Transfusional procederá a llevar dicha bolsa en frío al hospital asignado. Allí completará la preparación del producto enmascarado y administrará el tratamiento indicado al paciente participante utilizando para su infusión un acceso venoso previamente obtenido por un miembro del equipo del hospital. Solo este equipo (el encargado de aleatorización y el encargado de Medicina Transfusional Central de administrar el tratamiento) conocerán el producto a administrar a cada paciente (equipo no ciego). Ni el paciente ni los investigadores tendrán conocimiento del tratamiento administrado, realizando un estudio de doble ciego. El equipo no ciego llevará un registro de las asignaciones de producto y sus administraciones que será guardado de manera confidencial. En la eventual necesidad de una ruptura de ciego se podrá recurrir a dichos registros.

##### 5.2.d. Preparación y administración del artículo de prueba

La preparación y administración del plasma de convaleciente o placebo (solución fisiológica) se realizará acorde a las NORMAS ADMINISTRATIVAS Y TÉCNICAS RM 797/13 – 139/14 – 1507/15 Dirección de Sangre y Hemoderivados Ministerio de Salud de la Nación. (http://www.msal.gob.ar/disahe/images/stories/pdf/normas-hemoterapia.pdf).

Una vez enmascarado el producto, el equipo de Medicina Transfusional lo administrará en forma endovenosa lenta al paciente durante un mínimo de 1,5 horas y hasta 2 horas de acuerdo a las normas. Luego de la administración se monitoreará la ocurrencia de reacciones locales en el sitio de inyección y sistémicas por 12 horas. *Este es un estudio de mínimo riesgo, ya que la administración de plasma y de solución salina (placebo en este estudio) son de hábito diario en la práctica médica.* En el caso de una emergencia médica, cuando se tenga conocimiento de que la asignación de tratamiento puede influir en la atención médica del paciente, el investigador o la persona designada pueden solicitar que se rompa el ciego para el sujeto que está experimentando la emergencia. Sin embargo, antes del quiebre de ciego el investigador debe realizar todos los esfuerzos razonables de ponerse en contacto con la persona designada para discutir la decisión de quebrar el ciego. Se espera que el investigador proporcione una razón para la necesidad de romper el ciego, en base a un cambio significativo en la atención médica inmediata o de corto plazo del participante que resultará del conocimiento de la asignación del tratamiento.

##### 5.2.e. Toma de muestra de sangre para titulación de IgG anti-S SARS CoV2 en suero.

A las 24 horas de completada la infusión del tratamiento, se obtendrá una muestra de 5 ml de sangre venosa de todos los participantes para la titulación en suero de IgG anti-S SARS-CoV2, a realizarse en la Fundación INFANT por método ELISA (COVIDAR IgG, Leloir/CONICET). El suero se preservará a -20°C hasta su traslado en frío por un sistema de transporte autorizado para manejo de muestras biológicas a la Fundación.

Ventana de toma de muestra: +/- 4 horas. La muestra podrá tomarse en un plazo mínimo de 20 hs y uno máximo de 28 horas post-finalización de la administración del Producto de Investigación.

##### 5.2.f. Seguimiento clínico diario

Se realizará un control diario a partir del Dia 0, y por lo menos durante 15 días desde la infusión y hasta 25 días para aquellos sujetos que continúan hospitalizados a partir del día 15. Aquellos pacientes que sean dados de alta previo al día 15, serán monitoreados por un equipo de médicos domiciliarios entrenados para tal fin que trabajarán en colaboración con los centros de investigación. La siguiente información clínica se recolectará diariamente (utilizando un cuestionario diseñado para tal fin) por personal del estudio y **sin interferir con la atención médica del paciente ni alterar las normas recomendadas por las autoridades sanitarias para el manejo de los pacientes con COVID-19**: (a) frecuencia respiratoria (mirando como excursiona el tórax durante 30 segundos), (b) saturación de oxígeno, (c) temperatura, (d) frecuencia cardiaca central, (e) tos y duración, (f) dolor de garganta y duración, (g) sensación de fiebre/escalofríos, (h) necesidad de provisión de oxígeno suplementario y duración, (i) necesidad de cuidados intensivos y duración, (j) necesidad de asistencia respiratoria no invasiva o invasiva y duración, (k) evidencia de SDMO: según criterios de injuria renal aguda, criterios de elevación de enzimas hepáticas y criterios de miocardiopatía aguda (detallados en sección 2.2), (l) sobrevida.

#### Tabla 2. Cronograma de procedimientos del estudio.

| Día del Estudio | -1 Pesquisa | 0 Día 0 | Seguimiento Diario |
|---|---|---|---|
| Consentimiento informado de pesquisa para la evaluación de antecedentes, condición clínica y diagnóstico *SARS-CoV2* | X | | |
| Toma de hisopado nasofaríngeo para confirmación diagnóstica de SARS-CoV2 por RT-PCR | X[(1)] | | |
| Consentimiento informado de participación en el estudio | | X | |
| Revisión de historia médica | | X | X |
| Signos vitales | | X | |
| Confirmación de elegibilidad | | X | |
| Medicamentos concomitantes | | X | X |
| Asignación aleatoria de tratamiento | | X | |
| Administración de la intervención | | X | |
| Monitoreo de la transfusión | | X | |

| | | | |
|---|---|---|---|
| **Vigilancia de eventos adversos** | | X* | (X) |
| **Toma de muestra de sangre luego de completada la infusión** | | | Única muestra a las 24 hs (+/- 4 hs) |
| **Vigilancia de síntomas respiratorios** | | X | X |

(1) Este procedimiento no será requerido si se encuentra un resultado disponible por un método valido al momento de pesquisar el paciente

Se considerará que el paciente completa su participación al llegar el día 15 (si el paciente ya está dado de alta), al alta hospitalaria entre el día 15 y el día 25 o al cumplirse el día 25 de continuar hospitalizado.

#### 6. Comité Independiente de Monitoreo de Datos

Un Comité Independiente de Monitoreo de Datos supervisará la seguridad de los sujetos durante todo el estudio. El CIMD incluirá, como mínimo, miembros con experiencia en el manejo de medicina interna, enfermedades infecciosas, y un bio-estadístico con experiencia específica en el diseño, análisis y monitoreo de seguridad de estudios clínicos. El CIMD operará bajo un plan aprobado y tendrá la responsabilidad de monitorear las medidas de resultado / puntos finales, eventos adversos (EA) y eventos adversos serios (EAS), y recomendar la finalización del estudio si aparece en algún momento durante el estudio que los participantes (o un subgrupo de participantes) se encuentran en riesgo indebido como resultado de su participación. El CIMD se reunirá (por teleconferencia) semanalmente para revisar el acumulado de datos sobre seguridad y eficacia y realizará un análisis interino de los datos. El CIMD emitirá recomendaciones escritas en base a sus reuniones acerca de la continuidad del estudio. Las actas de cada reunión se registrarán en minutas documentadas en base a una agenda de trabajo preestablecida. Cualquier información de salud protegida específica del participante revisado por el CIMD se mantendrá completamente confidencial. Las sesiones serán cerradas sin acceso a terceros.

#### 7. Monitoreo de Seguridad

El investigador en cada institución y a nivel general supervisará la seguridad del paciente del estudio en su(s) sitio(s) según los requisitos de este protocolo y de acuerdo con las Buenas Prácticas Clínicas (GCP) actuales. El investigador monitoreará los datos de seguridad en todos los sitios de estudio. El monitoreo de seguridad se realizará de manera continua (por ejemplo, revisión individual de EAS, EAs y puntos finales) y de forma acumulada periódica.

##### Evento adverso (EA)

Un EA es cualquier acontecimiento médico desfavorable en un paciente al que se le administró un fármaco del estudio que puede tener o no una relación causal con el fármaco del estudio. Por lo tanto, una EA es cualquier signo desfavorable e involuntario (incluido el hallazgo anormal de laboratorio), síntoma o enfermedad que se asocia temporalmente con el uso de un medicamento del estudio, independientemente de si se considera relacionado con el medicamento del estudio.

Un EA también incluye cualquier empeoramiento (es decir, cualquier cambio clínicamente significativo en la frecuencia y / o intensidad) de una afección preexistente que se asocia temporalmente con el uso del producto en investigación.

La progresión de covid-19 no se considerará un EA si es claramente coherente con el patrón de progresión típico de la enfermedad

Si existe alguna incertidumbre acerca de que un EA se deba solo a la progresión de covid-19, se informará como un EA o EAS como se describe en la sección respectiva.

##### Evento adverso serio (EAS)

Un EAS, por definición, es cualquier evento médico desfavorable que a cualquier dosis:

- Resulte en la muerte: incluye todas las muertes, incluso aquellas que parecen no tener relación alguna con el fármaco del estudio (por ejemplo, un accidente automovilístico en el que un paciente es pasajero).
- Es potencialmente mortal: en opinión del investigador, el paciente corre un riesgo inmediato de muerte en el momento del evento. Esto no incluye un AE que si hubiera ocurrido en una forma más severa, podría haber causado la muerte.
- Requiere hospitalización o prolongación de la hospitalización existente. La hospitalización se define como el ingreso a un hospital o sala de emergencias por más de 24 horas. La prolongación de la hospitalización existente se define como una estadía en el hospital que es más larga de lo

previsto originalmente para el evento, o se prolonga debido al desarrollo de un nuevo EA según lo determinado por el investigador o el médico tratante.

- Resulta en discapacidad/incapacidad persistente o significativa (interrupción sustancial de la capacidad de uno para llevar a cabo funciones normales de la vida).
- Es un evento médico importante: los eventos médicos importantes pueden no poner en peligro la vida de inmediato o provocar la muerte u hospitalización, pero pueden poner en peligro al paciente o pueden requerir intervención para prevenir uno de los otros resultados graves enumerados anteriormente (por ejemplo, tratamiento intensivo en una emergencia o en casa para broncoespasmo alérgico; discrasias sanguíneas que no resultan en hospitalización).

En el caso específico de este estudio se considerarán como EA o EAS, según la descripción detallada en la sección 11.2, aquellos que se inicien dentro de las 12 horas posteriores a la administración del tratamiento y los agravamientos clínicos y muertes asociadas con estos eventos. No se considerarán EA o EAS aquellos eventos clínicos que se inicien luego de las 12 horas de iniciación del tratamiento, que constituirán parte de la evolución clínica de los pacientes enrolados infectados con SARS CoV2.

##### Periodo de Recolección de Eventos Adversos

El período de informe EA / EAS comienza cuando el participante se incluye inicialmente en el estudio (fecha de firma del consentimiento informado de participación) y recibe la infusión y continuará para los eventos que se inicien en las siguientes 12 horas desde su inicio (ver sección 11.2). Durante el seguimiento posterior a las 12 horas iniciales de la administración del producto de investigación, se monitoreará la evolución clínica asociada a los criterios de valoración primario y secundarios.

#### 8. Abandono Prematuro del Estudio

Los sujetos podrán abandonar el estudio en cualquier momento si así lo deseasen, pero serán igualmente monitoreados durante el periodo estipulado para garantizar su seguridad.

#### 9. Reemplazo de Pacientes

Aquellos pacientes que decidan abandonar el estudio antes de la administración de la droga serán reemplazados, de ser necesario, para asegurar un número adecuado de pacientes evaluables.

#### 10. Justificación del tamaño muestral y análisis estadístico

Existe una incertidumbre significativa en el tamaño del efecto esperado de la intervención, y considerando que se espera que el ensayo se complete en un período de tiempo relativamente corto, el estudio está diseñado para tener un análisis intermedio cuando los resultados del 50% de los sujetos hayan sido adquiridos.

Dada la relativa complejidad de implementar esta intervención, la diferencia mínimamente importante clínicamente se establece en una reducción relativa del 40%, para una tasa de resultado esperada del 50% en el grupo control que se reduce al 30% en el grupo de intervención. Un tamaño de muestra total de 210 sujetos (105 por brazo de prueba) tendrá una potencia del 80%, a un nivel de significancia (alfa) de 0.05 utilizando una prueba z de dos lados con corrección de continuidad. Estos resultados suponen que se realizan 2 pruebas secuenciales utilizando la función de gasto O'Brien-Fleming para determinar los límites de la prueba, como se describe en la tabla a continuación.

| **Look** | **Time** | **Lower Bndry** | **Upper Bndry** | **Nominal Alpha** | **Power** |
|---|---|---|---|---|---|
| 1 | 0.50 | -2.96259 | 2.96259 | 0.003 | 0.168 |
| 2 | 1.00 | -1.96857 | 1.96857 | 0.049 | 0.806 |

Bajo la estrategia de análisis primario, usaremos la distribución de límite de producto de Kaplan-Meier para comparar los grupos de tratamiento durante el tiempo necesario para alcanzar el resultado primario. También se informará una estimación del riesgo relativo y el intervalo de confianza del 95%.

Por recomendación del Comité Independiente de Monitoreo de Seguridad, se realizará además un análisis estratificado por edad (65-74 años y ≥75 años) y se ajustará el análisis estadístico por aquellos factores de riesgo cuya distribución entre grupos pueda afectar el resultado.

#### 11. Artículo de prueba y medicina transfusional

##### 11.1. Donación voluntaria de plasma de convaleciente

Todos los procedimientos se harán acorde al PLAN ESTRATÉGICO PARA REGULAR EL USO DE PLASMA DE PACIENTES RECUPERADOS DE COVID-19 CON FINES TERAPÉUTICOS. (IF-2020-26315442-APN-SCS#MS)

https://www.boletinoficial.gob.ar/detalleAviso/primera/227976/20200418

El plasma de convaleciente se obtendrá invitando a participar como donante voluntario a pacientes que hayan padecido la enfermedad covid-19 y se hayan recuperado satisfactoriamente en la Ciudad Autónoma y Provincia de Buenos Aires siguiendo los criterios de donación establecidos por las autoridades. Si el paciente desea donar, de forma voluntaria y altruista firmara el consentimiento de donación de plasma (ANEXO).

Los donantes serán identificados por las autoridades ministeriales e institucionales si se trata de un organismo privado, contactados, e invitados a donar en uno de los cinco Bancos de Sangre participantes de este estudio:

a) Instituto de Hemoterapia de la Provincia de La Plata
b) Hospital Militar Central
c) Fundación Hemocentro Buenos Aires
d) Fundación Hematológica Sarmiento
e) CEMIC

Para poder donar plasma, el paciente debe cumplir con las condiciones determinadas a la fecha (https://www.boletinoficial.gob.ar/detalleAviso/primera/227976/20200418) o seguir la actualización de ellas que realice el Ministerio.

##### Titulación de anticuerpos anti-SARS Cov2

Tras el proceso de la donación de plasma por aféresis o hemodonación, adicionalmente a las muestras que se toman para controles de rutina del banco de sangre, se obtendrá un tubo seco de 5 mL de sangre entera rotulados con la identificación dada al donante, identificación del Banco de Sangre y etiqueta que identifica la finalidad del estudio “PCC19” para la realización del título de anticuerpos. Estos se realizarán mediante el ensayo de ELISA anti-proteína S de SARS CoV2, que a luz de los datos existentes sobre protección contra el virus permiten una extrapolación indirecta de la capacidad de neutralización del suero, optimizado por el Instituto Leloir a cargo de la Dra. Gamarnik, y actualmente utilizado por los equipos de salud nacionales para estudiar prevalencia de inmunidad. Asimismo, de ser posible, se correlacionará este dato con el título de anticuerpos neutralizantes contra un pseudovirus codificando la proteína S optimizado por el mismo laboratorio.

El tubo será almacenado en un lugar designado dentro del Banco y enviado en un transporte autorizado de muestras biológicas al laboratorio de la Fundación INFANT (con más de 18 años de experiencia en realización de ensayos de estas características) donde se realizará la técnica de ELISA y la titulación de anticuerpos neutralizantes.

Los títulos obtenidos por ELISA de S serán luego clasificados y aquellos que estén por encima de 1:1.000 seleccionados para proveer plasma de convaleciente para este estudio.

Los plasmas de alto título adicionales serán almacenados asimismo para enviar al Centro de Hemoderivados de Córdoba (UNC) que producirá gammaglobulina enriquecida con anticuerpos anti-SARS-CoV2.

**Plasmaféresis (Instituto de Hemoterapia de la Provincia de La Plata, Hospital Militar Central, Fundación Hemocentro Buenos Aires, y Fundación Hematológica Sarmiento)**

La donación de plasma se efectuará a través de un procedimiento de aféresis con equipos e insumos descartables aprobados para tal fin, durante el cual se recomienda extraer un volumen no mayor al 15% de la volemia del donante. El anticoagulante a utilizar será ACD-A o similar. En los casos en que no se realizare la reposición del volumen, la extracción de plasma no deberá superar los 600 ml por procedimiento.
Se deberá respetar el intervalo de 48 hs. entre cada procedimiento, y no superar las 2 donaciones en una semana o 24 donaciones en un periodo de 12 meses.
Las unidades de PCC19 se podrán separar en alícuotas de 300ml respetando el circuito cerrado y siguiendo las indicaciones del procedimiento operativo estándar vigente.

**Hemodonación (CEMIC)**

1-La obtención de plasma se realizará con un equipo Cobe Spectra. Se extraerán 1,5 a 2 L de plasma de acuerdo al peso y tolerancia al procedimiento del donante. En caso de inaccesibilidad venosa u oposición del donante, se extraerá entre 400 y 500 ml de sangre entera (hemodonación clásica)
2-El reemplazo de volumen se efectuará con solución fisiológica y de albúmina.
3-Durante el procedimiento el plasma será derivado a bolsas de transferencia de similar volumen cada una (aproximadamente 500ml). Se denominará Bolsa 1 a la inicialmente obtenida, Bolsa 2 la media y Bolsa 3 la final.
4-Las bolsas recolectadas serán rotuladas como detallado abajo
5-Se congelarán en el ultrafreezer vertical Righi de -56°C (rotulado como Freezer # I, identificado con rótulo de plasma de convaleciente COVID-19, en el Banco de Sangre) separadas en estantes por tipo de bolsa (1, 2 o 3).

6-El plasma de convaleciente permanecerá almacenado hasta su uso o caducidad (1 año), con criterios de seguridad apropiados.
7-El descarte se efectuará bajo normas de seguridad que se estipulen.
8- Se guardarán alícuotas de suero/plasma para futuras determinaciones congelado a -80ºC, en tubos de Eppendorf del PC de las diferentes etapas, y congelados y almacenados a -80°C: Previo al procedimiento de aféresis de las bolsas 1, 2 y 3 (previo al congelamiento) luego del descongelamiento de las bolsas 1, 2 y 3.

**Etiquetado**

Todas las unidades de PC COVID-19 deben cumplir con el etiquetado requerido para el plasma fresco congelado de cada Banco de Sangre y además deberán tener una clara identificación, en la que deberá constar:

· "Tipo de Producto: PC COVID-19"
· "Resultado de título de anticuerpos para SARS-Cov-2" (si están disponibles)
· "PRECAUCIÓN: producto de uso exclusivo para investigación"
. "PROTOCOLO: Fundación INFANT – Suero enriquecido con anticuerpos anti SARS-CoV2"

**Cadena de Frío de PCCOVID-19**

1. Las unidades se almacenarán debidamente separadas del resto de las unidades de plasma habilitadas para uso transfusional, en un lugar claramente identificado para tal fin.

2. Las unidades de PCC19 se almacenarán a temperatura inferior a -25 C, la cual permite un almacenamiento de 36 meses y entre -18C y -25 para 3 meses de almacenamiento.

3. El transporte de las unidades se realizará de forma tal que se garantice fehacientemente el mantenimiento de la cadena de frío.

4. La conservación de las unidades de plasma una vez descongeladas deberá realizarse entre 2 y 6ºC hasta las 24 horas de finalizado el descongelamiento. Pasado ese tiempo, se deberán descartar.

**Trazabilidad**

El Banco de Sangre y el Servicio de Transfusión de la Institución solicitante implementarán un sistema de registros que garantice la trazabilidad entre donantes y receptores. En esta situación especial, además, deberán disponer de un registro específico por tratarse de una terapia transfusional administrada en un contexto experimental.

#### 11.2. Administración de la intervención

Una vez evaluados los signos vitales y verificada la elegibilidad, el equipo no ciego de Medicina Transfusional Central será notificado del resultado del proceso de asignación aleatoria y se trasladará al hospital asignado con el producto de tratamiento enmascarado y en frío. Al arribar procederá nuevamente verificar el código de tratamiento asignado con el miembro de laboratorio de Medicina Transfusional del hospital correspondiente (ver 5.2.c).

Como placebo, se utilizarán 250 ml de solución salina al 0,9% estéril enmascarada y administrada de la misma forma y velocidad que el plasma. Para evitar sesgos y mantener el ciego del estudio, el plasma y la solución salina al 0,9% serán enmascarados, utilizando una bolsa opaca y cinta que no permitirán diferenciar de qué producto se trata. El miembro de Medicina Transfusional Central será quien entregue al miembro del equipo local para que administre la intervención en conjunto con el equipo central. De esta manera tanto los miembros de Medicina Transfusional Central como los miembros de Medicina Transfusional de los hospitales serán no ciegos, mientras que todo el resto del equipo delegado en funciones en el estudios en cada Centro de Investigación como los pacientes serán ciegos a la intervención.

Para preparar el plasma, se descongelará la bolsa de 250 ml a 37°C, según procedimientos operativos del Departamento de Medicina Transfusional del Hospital. Se infundirá al receptor en forma lenta (en no menos de 1.5 horas y hasta 2 horas) de acuerdo a las condiciones hemodinámicas del paciente y se monitorearán por 12 horas y registrarán los efectos adversos tempranos y tardíos atribuibles a la transfusión.

Ventana nocturna para pasaje del producto de investigación: entre las 20.00 hs y las 8.00 hs, se tolerará una ventana de +8 horas para el pasaje del Producto de investigación. Siendo posible un máximo de 80 hs (72+8) de comienzo de síntomas al momento del inicio de la infusión. Esta excepción sólo será permitida en este rango horario nocturno.

##### Monitoreo de eventos adversos asociados a la transfusión de plasma

###### *Sobrecarga circulatoria asociada a transfusión (TACO)*

La sobrecarga circulatoria asociada a transfusión (transfusion-associated circulatory overload, TACO) es la complicación pulmonar más frecuente y es un factor de riesgo independiente para morbimortalidad hospitalaria, con mayor incidencia en pacientes críticos [58]. La frecuencia estimada de TACO varía del 1% a 5% según el sistema de hemovigilancia sea [59], hasta los 8% en pacientes post quirúrgicos de avanzada, y 11% en pacientes críticos [60 61 62]. Los factores de riesgo incluyen enfermedad cardíaca, pulmonar o renal, edad ≥70 años y un balance de fluidos positivo, pre transfusional [63 64]. La mayoría de los casos de TACO se previenen disminuyendo la velocidad de transfusión y la instauración de las medidas habituales para sobrecarga hídrica en poblaciones susceptibles. La sobrecarga constituye un EA solicitado, pero de no mejorar en el curso de 120 minutos después de la infusión o de requerir -a juicio de los profesionales responsables- el paso del paciente a cuidados intensivos por mala progresión clínica a causa de dicha sobrecarga, deberá informarse de inmediato como un EAS.

*Criterios diagnósticos de TACO* [19]
Durante o hasta 12 horas posteriores a la transfusión, TACO se caracteriza por la presencia de un total de 3 o más de los criterios listados:
A. Compromiso respiratorio agudo o que empeora
B. Evidencia de edema pulmonar agudo o que empeora basado en la clínica y / o imágenes radiográficas del tórax y / u otra evaluación no invasiva de la función cardíaca (ecocardiograma)
C. Evidencia de cambios cardiovasculares no explicados por la condición médica subyacente del paciente, incluido: taquicardia, hipertensión, presión de pulso ensanchada, distensión venosa yugular, silueta cardíaca agrandada y / o edema periférico
D. Evidencia de sobrecarga de líquidos, incluyendo cualquiera de los siguientes: un balance positivo de líquidos; respuesta a la terapia diurética, o diálisis combinada con mejoría clínica; y variación del peso del paciente
E. Aumento del nivel de péptido natriurético de tipo B (por ejemplo, BNP o NT-pro BNP) por encima del rango de referencia específico del grupo de edad y mayor de 1,5 veces el valor de pre transfusión.

***Injuria Pulmonar Aguda Asociada a Transfusión (TRALI)***
Es un síndrome de dificultad respiratoria aguda, será considerado un EAS y ocurre durante o dentro de las seis horas posteriores a la administración de la transfusión. Estudios prospectivos de TRALI en diversas encontraron tasas de incidencia muy bajas (0,0008% a 0,001% de pacientes transfundidos) [19] después de la implementación de estrategias para prevenir la transfusión de componentes de la sangre obtenido de donantes femeninas multíparas. En nuestro estudio no se transfundirá plasma convaleciente de mujeres multíparas.
El tratamiento del paciente con TRALI incluye la interrupción inmediata de la transfusión.

*Criterios diagnósticos de TRALI: [28]*

TRALI Tipo I: pacientes que no tienen factores de riesgo de SDRA y cumplen con los siguientes criterios:

A. i. Inicio agudo

ii. Hipoxemia ($PaO_2$ / $FiO_2$ ≤ 300 o $SpO_2$ <90% en aire ambiente).

iii. Evidencia clara de edema pulmonar bilateral en imágenes (radiografía de tórax, tac o ecografía)

iv. No hay evidencia de hipertrofia ventricular izquierda o, si está presente, no es la principal contribuyente de la hipoxemia

B. Inicio durante o dentro de las 6 horas de la transfusión

C. Sin relación temporal con un factor de riesgo alternativo para SDRA

TRALI Tipo II: pacientes que tienen factores de riesgo de SDRA (pero que no han sido diagnosticados con SDRA) o que tienen SDRA leve existente (Pa / Fi de 200-300), pero cuyo estado respiratorio se deteriora y se considera que se debe a una transfusión basado en:

a. Los resultados descritos en las categorías a y b de TRALI Tipo I, y

b. Estado respiratorio estable en las 12 horas previas a la transfusión

***Reacciones Alérgicas – Anafilácticas***

Las reacciones alérgicas ocurren en general durante o dentro de las 4 horas posteriores a la transfusión y están más frecuentemente asociadas a la transfusión de plaquetas (302 cada 100 000 unidades de plaquetas) [29]. Los síntomas son causados por mediadores como la histamina, liberados en la activación de mastocitos y basófilos [30]. En general la presentación clínica es leve (erupción cutánea, prurito, urticaria, y angioedema localizado) y constituye un EA solicitado. Las reacciones transfusionales alérgicas leves en general resuelven con la administración del tratamiento habitual para reacciones alérgicas, pudiendo reiniciar la infusión de la unidad. La transfusión debe suspenderse si los síntomas reaparecen. La incidencia de reacciones anafilácticas es de 8/100,000 unidades de plaquetas transfundidas. Las reacciones más severas generalmente se presentan con broncoespasmo, dificultad respiratoria e hipotensión y constituyen un EAS y debe ser reportado como tal inmediatamente [31-35].

***Reacciones transfusionales hemolíticas agudas***

Las reacciones de transfusionales hemolíticas agudas (RTHA) pueden ocurrir cuando se transfunden glóbulos rojos incompatibles o en menor frecuencia, grandes cantidades de plasma incompatible con el ABO del paciente. El mecanismo fisiopatológico central es la hemólisis intravascular y se considerará un EAS. Se presenta con fiebre repentina y escalofríos, dolor retroperitoneal y disnea, hemoglobinuria y hasta coagulación intravascular diseminada, insuficiencia renal aguda y shock. Dado que la fiebre y los escalofríos pueden ser los únicos signos tempranos, es importante controlar al paciente durante la transfusión y detenerla transfusión inmediatamente si hay algún cambio en los signos vitales o ante la aparición de síntomas inesperados. El tratamiento se basa en medidas de sostén para mitigar los síntomas. La identificación adecuada del paciente y el cumplimiento de todos los procesos relacionados con el manejo de muestras pre-transfusionales y con la administración de la transfusión, son esenciales para prevenir la RTHA [36].

***Infecciones Transmisibles por Transfusión [39]***
El riesgo de transmisión de enfermedades infecciosas por transfusiones se ha reducido dramáticamente, secundario a los grandes avances en el testeo de las unidades, inactivación de patógenos para unidades de plasma y sus componentes [51] y a la implementación de rigurosos métodos de selección de donantes. En nuestro medio, las muestras son testeadas para HBV, HCV, HIV, HTLV y *Trypanosoma cruzi* [52]. En Estados Unidos, el riesgo de adquirir HBV y HIV por transfusión es de 1:280.000 y 1:1.467.000, respectivamente [53 54]. Gracias al congelamiento de las muestras de plasma, la contaminación de las mismas por patógenos bacterianos y CMV es extremadamente rara. Hay solo una decena de casos entre los millones de transfusiones realizadas, reportados en Alemania y Canadá de transmisión bacteriana. Se estima que la fuente potencial de esta contaminación habrían sido los baños de agua utilizados para el descongelamiento de las unidades. Esto es fácilmente evitable con la limpieza y esterilización correctas de los descongeladores [39].

#### 12. Consideraciones éticas

##### Declaración de Buena Práctica Clínica

Este estudio se llevará a cabo de conformidad con el protocolo y con las siguientes consideraciones:

- los principios éticos que tienen su origen en la Declaración de Helsinki,
- las pautas vigentes de las buenas prácticas clínicas (Good Clinical Practice, GCP) del Consejo Internacional de Armonización (International Council of Harmonisation, ICH),
- las leyes y regulaciones vigentes.

El protocolo, los formularios de Consentimiento Informado (FCI) y otros documentos relevantes han sido revisados y aprobados por Fundación INFANT, las autoridades participantes y enviados al Comité de Ética (CE) para su evaluación.

##### Comité de ética (CE):

Un CE debidamente constituido, como se describe en las pautas de ICH para GCP, debe revisar y aprobar:

- El protocolo, FCI y cualquier otro material que se proporcione a los participantes antes de que cualquier sujeto pueda ingresar en el estudio
- Cualquier enmienda o modificación al protocolo de estudio o al FCI antes de la implementación, a menos que el cambio sea necesario para eliminar un peligro inmediato para los pacientes, en cuyo caso el CE debe ser informado lo antes posible

Además, debe ser informado de cualquier evento que pueda afectar la seguridad de los pacientes o la realización continua del estudio clínico.

INFANT debe recibir una copia de la carta de aprobación del CE antes de enviar los suministros de medicamentos al investigador de cada hospital. La carta de aprobación debe incluir el título del estudio, los documentos revisados y la fecha de la revisión.

El investigador debe mantener en el archivo los registros de la revisión del CE y la aprobación de todos los documentos del estudio

##### Proceso de Consentimiento Informado

El FCI utilizado por el investigador será el aprobado por el CE correspondiente.

Es responsabilidad del investigador o profesional delegado obtener el consentimiento informado por escrito de cada sujeto antes de su participación en el estudio y después de que los objetivos, procedimientos y riesgos potenciales del estudio se han explicado completamente en un lenguaje que el sujeto pueda entender. El FCI debe estar firmado y fechado por el investigador o personal delegado que realizó el proceso de toma de consentimiento.

El investigador o personal delegado le explicará al participante la naturaleza del estudio y responderá todas las preguntas sobre el estudio. Les informará que su participación es voluntaria. El participante deberá firmar el formulario de consentimiento informado (FCI) antes de que se realice cualquier actividad específica del estudio.

A los participantes que pueden entender pero que no pueden escribir y/o leer se les leerá el CI en presencia de un testigo imparcial, que firmará y fechará el FCI para confirmar que se obtuvo el consentimiento informado.

Se firmarán dos ejemplares. El investigador debe conservar un ejemplar como parte del registro del estudio del paciente, y se debe entregar el otro ejemplar firmado al participante.

- Pacientes con vulnerabilidad cognitiva:
  Podrán ser incluidos previa toma de consentimiento por parte de su representante según corresponda:
  a. Pacientes que posean una declaración jurídica de incapacidad: deberá ser quien haya sido designado como curador siempre que exista esa declaración
  b. Pacientes que puedan tomar una decisión por si mismos previa asistencia de un "apoyo" según las disposiciones del Código Civil: por lo general el paciente puede designar a quien lo ayude a decidir.
  c. Pacientes sin declaración judicial de incapacidad y que tampoco hayan designado un apoyo o representante: resultará de aplicación la ley de derechos de los pacientes.
  Los medios para acreditar la representación pueden consistir en cualquier documento que acredite la designación como curador (si aplica) o bien cualquier directiva anticipada del paciente en donde establezca quién lo representa en temas de salud, o documentos que acrediten el parentesco según la Ley 26529.

#### 13. Confidencialidad y Protección de Datos Personales

El investigador tomará todas las medidas apropiadas para garantizar que se mantenga el anonimato de cada sujeto del estudio en los registros para investigación transmitidos fuera del centro de salud. Los documentos originales y esenciales del estudio deben mantenerse en estricta confidencialidad en cada centro de investigación por el equipo del estudio.

Los datos personales de los participantes se almacenarán en el centro del estudio en un formato impreso y/o electrónico protegidos por contraseña o en un cuarto bajo llave, para garantizar el acceso exclusivo del personal autorizado del estudio.

Para proteger los derechos y las libertades de las personas físicas en relación con el tratamiento de datos personales, se asignará a los participantes un código alfanumérico único y específico. Los registros o conjuntos de datos de los participantes que se transfieran contendrán el código alfanumérico; los nombres de los participantes no se transferirán. Todos los demás datos identificables transferidos entre los investigadores se identificarán mediante este código único y específico del participante. El centro del estudio mantendrá una lista confidencial de los participantes que participaron en el estudio bajo estricta seguridad (protegidos por contraseña o en un cuarto bajo llave), vinculando el código alfanumérico de cada participante con su identidad real.

#### 14. Documentación del Estudio

Los registros y los documentos, incluidos los FCI firmados, respecto a la realización de este estudio, serán conservados por el investigador durante 15 años después de la finalización del estudio, a menos que las regulaciones locales o las políticas institucionales requieran un período de conservación más prolongado.
El investigador de cada institución participante deberá consultar con INFANT antes de descartar o destruir cualquier documento esencial del estudio después de la finalización o interrupción del estudio. En caso de ser aprobado, los registros deben destruirse de manera que se garantice la confidencialidad.

#### 15. Monitoreo de estudio

##### 15.1. Monitoreo de sitios de estudio

El monitor del estudio visitará cada sitio antes de inscribir al primer paciente y periódicamente durante el estudio de acuerdo con el Plan de Monitoreo del Estudio Aprobado.

El monitor recibirá semanalmente de cada uno de los centros un informe con el número de pacientes en periodo de pesquisa y enrolados y verificará que el envió de datos se encuentre al día y completo, realizando de ser necesario seguimiento a cada uno de los centros.

Cada centro contará con una persona designada por el Investigador a verificar procesos realizados bajo protocolo y datos originales y se documentará dicha verificación por escrito.

Las visitas de monitoreo y las actividades de monitoreo centralizado se realizarán de acuerdo con todos los requisitos reglamentarios aplicables y normas vigentes en Argentina. Se entiende que el monitor contactará a cada investigador y a su equipo responsable regularmente y se le permitirá verificar la generación de los distintos registros del estudio.

Será responsabilidad del monitor inspeccionar los sistemas de captura de datos a intervalos regulares durante todo el estudio para verificar el cumplimiento del protocolo y la integridad, precisión y consistencia de los datos; y adhesión al ICH GCP y normativa local sobre la realización de investigaciones clínicas. El monitor debe tener acceso a informes de laboratorio, medicación de estudio, registros asociados con él, y otros registros médicos necesario para verificar las entradas en el EDC.

Se asignará un **monitor no ciego** cuyo rol estará limitado a auditar los datos de logística manejo y registro del tratamiento de estudio de acuerdo con el protocolo aprobado y las instrucciones de tratamiento:

- recepción y condiciones de almacenaje
- randomización y administración
- retorno o destrucción/ reconciliación final

##### 15.2. Requisitos del documento fuente

Se requiere que los investigadores preparen y mantengan registros de pacientes adecuados y precisos (documentos fuente).

El investigador debe mantener todos los documentos fuente en el archivo de los datos volcados al formulario del estudio (CRF). Algunos datos generados del estudio podrán volcarse directamente en el sistema de EDC (captura directa de datos), previa aprobación de dicho proceso por los Comités de Ética intervinientes. Los formularios de informes de casos y los documentos fuente deben estar disponibles en todo momento.

##### 15.3. Requisitos del formulario de informe de caso

Los datos del estudio obtenidos en el curso del estudio serán registrados en formularios de informes de casos (CRF) y enviados a INFANT en formato scan. Una vez recibidos serán archivados y procesados mediante el ingresados al del sistema EDC por personal capacitado de INFANT. Todos los CRF requeridos deben completarse para cada paciente inscrito en el estudio. El investigador debe conservar una copia del libro de casos de CRF de cada paciente como parte del registro del estudio y debe estar disponible en todo momento para su inspección por parte de representantes autorizados del patrocinador y las autoridades reguladoras.

#### 16.. Auditorias e inspecciones

Este estudio puede estar sujeto a una auditoría o inspección de garantía de calidad. Si esto ocurre, el investigador es responsable de:

- Brindar acceso a todas las instalaciones, datos de estudio y documentos necesarios para la inspección o auditoría.
- Comunicar cualquier información que surja de la inspección de las autoridades reguladoras a INFANT de inmediato.
- Tomar todas las medidas apropiadas solicitadas por INFANT para resolver los problemas encontrados durante la auditoría o inspección.

Los documentos sujetos a auditoría o inspección incluyen, entre otros, todos los documentos fuente, CRF, registros médicos, correspondencia, FCI, archivos del CE, documentación de certificación y control de calidad de laboratorios de apoyo, y registros relevantes para el estudio que se mantienen Instalaciones de farmacia. Las condiciones de almacenamiento del material de estudio también están sujetas a inspección. Además, los representantes del patrocinador pueden observar la conducta de cualquier aspecto del estudio clínico o sus actividades de apoyo tanto dentro como fuera de la institución del investigador.

En todos los casos, se debe respetar la confidencialidad de los datos.

### ANEXO

#### INSUFICIENCIA CARDIACA
#### ESCALA NYHA (NEW YORK HEART ASSOCIATION)
#### VALORACIÓN FUNCIONAL DE INSUFICIENCIA CARDÍACA.

| CLASIFICACIÓN FUNCIONAL NYHA | |
|---|---|
| Clase I | Sin limitación de la actividad física. El ejercicio físico normal no causa fatiga, palpitaciones o disnea. |
| Clase II | Ligera limitación de la actividad física. Sin síntomas en reposo. La actividad ordinaria ocasiona fatiga, palpitaciones o disnea. |
| Clase III | Marcada limitación de la actividad física. Sin síntomas en reposo. Actividad física menor que la ordinaria ocasiona fatiga, palpitaciones o disnea. |
| Clase IV | Incapacidad para llevar a cabo cualquier actividad física; los síntomas de insuficiencia cardíaca están presentes incluso en reposo y aumentan con cualquier actividad física. |

#### CIRROSIS (CONSENSO HIPERTENSION PORTAL BAVENO IV)

| ESTADIOS DE CIRROSIS | |
|---|---|
| Estadío 1 | Ausencia de varices esofágicas y de ascitis |
| Estadío 2 | Varices esofágicas sin antecedente de hemorragia y sin ascitis. |
| Estadío 3 | Presencia de ascitis con o sin varices esofágicas. |
| Estadío 4 | Hemorragia gastrointestinal por hipertensión portal, con o sin ascitis. |

#### ENFERMEDAD RENAL CLASIFCIACION KDIGO.

| GRADO | FILTRACIÓN GLOMERULAR ML/MIN/1,73 M2 | DESCRIPCION |
|---|---|---|
| GRADO 1 | > 90 G5 | Normal o elevado |
| GRADO 2 | 60-89 | Ligeramente disminuido |
| GRADO 3a | 45-59 | Ligera a moderadamente disminuido |
| GRADO 3b | 30-44 | Moderada a gravemente disminuido |
| GRADO 4 | 15-29 | Gravemente disminuido |
| GRADO 5 | < 15 | Fallo renal |

#### Referencias

1. Fei Zhou*, Ting Yu*, Ronghui Du*, Guohui Fan*, Ying Liu*, Zhibo Liu*, Jie Xiang*, Yeming Wang, Bin Song, Xiaoying Gu, Lulu Guan, Yuan Wei, Hui Li, Xudong Wu, Jiuyang Xu, Shengjin Tu, Yi Zhang, Hua Chen, Bin Cao. Clinical course and risk factors for mortality of adult inpatients with COVID-19 in Wuhan, China: a retrospective cohort study. Lancet 2020; 395: 1054–62 doi: 10.1016/S0140-6736(20)30566-3

2. Chen N., Zhou M., Dong X., Qu J., Gong F., Han Y. Epidemiological and clinical characteristics of 99 cases of 2019 novel coronavirus pneumonia in Wuhan, China: a descriptive study. Lancet. 2020;395:507–513. 10.1016/S0140-6736(20)30211-7

3. Garg S, Kim L, Whitaker M, et al. Hospitalization Rates and Characteristics of Patients Hospitalized with Laboratory-Confirmed Coronavirus Disease 2019 — COVID-NET, 14 States, March 1–30, 2020. MMWR Morb Mortal Wkly Rep 2020;69:458–464. DOI: http://dx.doi.org/10.15585/mmwr.mm6915e3

4. Ministerio de Salud de Argentina. www.argentina.gob.ar/salud/coronavirus-COVID-19

5. Willan, J., King, A. J., Jeffery, K., & Bienz, N. (2020). Challenges for NHS hospitals during covid-19 epidemic. BMJ, m1117.doi:10.1136/bmj.m1117

6. Goodarz Kolifarhood,[1,3] Mohammad Aghaali,[1] Hossein Mozafar Saadati,[1] Niloufar Taherpour,[1]Sajjad Rahimi,[1,2] Neda Izadi,[3] and Seyed Saeed Hashemi Nazari[4,*] . Epidemiological and Clinical Aspects of COVID-19; a Narrative Review. Arch Acad Emerg Med. 2020; 8(1): e41. PMCID: PMC7117787. PMID: 32259130

7. Center for Disease Control and Prevention Interim Guidance for Healthcare Facilities: Preparing for Community Transmission of COVID-19 in the United States

8. Orenstein, W. A., Heymann, D. L., Ellis, R. J., Rosenberg, R. L., Nakano, J., Halsey, N. A.,Witte, J. J. (1981). Prophylaxis of varicella in high-risk children:

Dose-response effect of zoster immune globulin. The Journal of Pediatrics, 98(3), 368–373.doi:10.1016/s0022-3476(81)80697-x

9. Centers for Disease Control and Prevention. Measles (Rubeola) For Healthcare Professionals.

20. Yun Feng ; Yun Ling , Tao Bai , Yusang Xie ; Jie Huang , Jian Li , Weining Xiong , Dexiang Yang , Rong Chen ; Fangying Lu ; Yunfei Lu , et al. COVID-19 with Different Severity: A Multi-center Study of Clinical Feature. doi.org/10.1164/rccm.202002-0445OC

21. Zhou F, Yu T, Du R, Fan G, Liu Y, Liu Z, Xiang J, Wang Y, Song B, Gu X, Guan L, Wei Y, Li H, Wu , Xu J, Tu S, Zhang Y, Chen H, Cao B. Clinical course and risk factors for mortality of adult inpatients with COVID-19 in Wuhan, China: a retrospective cohort study. Lancet. 2020 Mar 28;395(10229):1054-1062. doi: 10.1016/S0140-6736(20)30566-3

22. Afaf Alblooshi, Alia Alkalbani, Ghaya Albadi, Hassib Narchi, and Graham Hall. Is forced oscillation technique the next respiratory function test of choice in childhood asthma.World J Methodol. 2017 Dec 26; 7(4): 129–138. doi: 10.5662/wjm.v7.i4.129

23. Garg S, Kim L, Whitaker M, et al. Hospitalization Rates and Characteristics of Patients Hospitalized with Laboratory-Confirmed Coronavirus Disease 2019 — COVID-NET, 14 States, March 1–30, 2020. MMWR Morb Mortal Wkly Rep 2020;69:458–464. DOI: http://dx.doi.org/10.15585/mmwr.mm6915e3

24. Feng, Y. et al. COVID-19 with Different Severity: A Multi-center Study of Clinical Features. Am. J. Respir. Crit. Care Med. **0**, null. https://doi.org/10.1164/rccm.202002-0445OC

25. Grasselli G, Zangrillo A, Zanella A, et al. Baseline Characteristics and Outcomes of 1591 Patients Infected With SARS-CoV-2 Admitted to ICUs of the Lombardy Region, Italy. *JAMA.* Published online April 06, 2020. doi:10.1001/jama.2020.5394

26. Boletín Integrado de Vigilancia. Dirección Nacional de Epidemiología y Análisis de la Situación de Salud. N° 424 – SE 39 – Septiembre de 2018

32. Chen T, Dai Z, Mo P, Li X, Ma Z, Song S, Chen X, Luo M, Liang K, Gao S, Zhang Y, Deng L, Xiong Y. Clinical characteristics and outcomes of older patients with coronavirus disease 2019 (COVID-19) in Wuhan, China (2019): a

single-centered, retrospective study. The Journals of Gerontology: Series A, glaa089, https://doi.org/10.1093/gerona/glaa089

33. Q. Li, X. Guan, P. Wu, et al. Early Transmission Dynamics in Wuhan, China, of Novel Coronavirus–Infected Pneumonia. N Engl J Med (January 2020), doi: 10.1056/NEJMoa2001316NEJMoa2001316

34. Y. Liu, A.A. Gayle, A. Wilder-Smith, J. Rocklöv, The reproductive number of COVID-19 is higher compared to SARS coronavirus. J Travel Med (February 2020), 10.1093/jtm/taaa021

35. K. Duan, et al. The feasibility of convalescent plasma therapy in severe COVID-19 patients: a pilot study. medRxiv 2020.03.16.20036145; doi: https://doi.org/10.1101/2020.03.16.20036145

36. Cheryl D. Fryar, M.S.P.H., Yechiam Ostchega, Ph.D., R.N., Craig M. Hales, M.D., M.P.H., Guangyu Zhang, Ph.D., and Deanna Kruszon-Moran, M.S. Hypertension Prevalence and Control Among Adults: United States, 2015–2016. U.S. DEPARTMENT OF HEALTH AND HUMAN SERVICES. Center for Disease Control and Prevention.

37. U.S. Department of Health and Human Services, Center for Disease Control and Prevention. National Diabetes Statistics Report, Estimates of Diabetes and Its Burden in the United States

38. Craig M. Hales, M.D., Margaret D. Carroll, M.S.P.H., Cheryl D. Fryar, M.S.P.H., and Cynthia L. Ogden, Ph.D. Prevalence of Obesity and Severe Obesity Among Adults: United States, 2017–2018. U.S. DEPARTMENT OF HEALTH AND HUMAN SERVICES. Center for Disease Control and Prevention.

39. Lara J. Akinbami, M.D.; and Xiang Liu, M.Sc. Chronic Obstructive Pulmonary Disease Among Adults Aged 18 and Over in the United States, 1998–2009. Center for Disease Control and Prevention.

40. Coronavirus Disease 2019 (COVID-19) Daily Data Summary, NYC Health, April 16, 2020. https://www1.nyc.gov/assets/doh/downloads/pdf/imm/covid-19-daily-data-summary-deaths-04172020-1.pdf

41. Neutralizing antibody responses to SARS-CoV-2 in a COVID-19 recovered patient cohort and their implications Fan Wu, Aojie Wang, Mei Liu, Qimin Wang,

Jun Chen, Shuai Xia, Yun Ling, Yuling Zhang, Jingna Xun, Lu, Shibo Jiang, Hongzhou Lu, Yumei Wen, Jinghe Huang medRxiv 2020.03.30.20047365; doi: https://doi.org/10.1101/2020.03.30.20047365

42. Reinfection could not occur in SARS-CoV-2 infected rhesus macaques, Linlin Bao, Wei Deng, Hong Gao, Chong Xiao, Jiayi Liu, Jing Xue, Qi Lv, Jiangning Liu, Pin Yu, Yanfeng Xu, Feifei Qi, Yajin Qu, Fengdi Li, Zhiguang Xiang, Haisheng Yu, Shuran Gong, Mingya Liu, Guanpeng Wang, Shunyi Wang, Zhiqi Song, Wenjie Zhao, Yunlin Han, Linna Zhao, Xing Liu, Qiang Wei, Chuan Qin bioRxiv 2020.03.13.990226; doi: https://doi.org/10.1101/2020.03.13.990226

43. Rhinesmith, E., & Fu, L. (2018). Tetanus Disease, Treatment, Management. Pediatrics in Review, 39(8), 430–432. doi:10.1542/pir.2017-0238

44. Christenson, J. C., & Manaloor, J. J. (2016). Hepatitis A, B, and C. Pediatrics in Review, 37(10), 426–438. doi:10.1542/pir.2015-0075

45. John Hodgson.The pandemic pipeline. Nature Biotechnology. doi: 10.1038/d41587-020-00005-z

46. W. Guan, Z. Ni, Yu Hu, W. Liang, C. Ou, J. He, L. Liu, H. Shan, C. Lei, D.S.C. Hui, B. Du, L. Li, G. Zeng, K.-Y. Yuen, R. Chen, C. Tang, T. Wang, P. Chen, J. Xiang, S. Li, Jin-lin Wang, Z. Liang, Y. Peng, L. Wei, Y. Liu, Ya-hua Hu, P. Peng, Jian-ming Wang, J. Liu, Z. Chen, G. Li, Z. Zheng, S. Qiu, J. Luo, C. Ye, S. Zhu, and N. Zhong, for the China Medical Treatment Expert Group for Covid-19. Clinical Characteristics of Coronavirus Disease 2019 in China. The New England Journal of Medicine. DOI: 10.1056/NEJMoa2002032

47. Temet M. McMichael, Ph.D., Dustin W. Currie, Ph.D., Shauna Clark, R.N., Sargis Pogosjans, M.P.H., Meagan Kay, D.V.M., Noah G. Schwartz, M.D., James Lewis, M.D., Atar Baer, Ph.D., Vance Kawakami, D.V.M., Margaret D. Lukoff, M.D., Jessica Ferro, M.P.H., Claire Brostrom-Smith, M.S.N., Thomas D. Rea, M.D., Michael R. Sayre, M.D., Francis X. Riedo, M.D., Denny Russell, B.S., Brian Hiatt, B.S., Patricia Montgomery, M.P.H., Agam K. Rao, M.D., Eric J. Chow, M.D., Farrell Tobolowsky, D.O., Michael J. Hughes, M.P.H., Ana C. Bardossy, M.D., Lisa P. Oakley, Ph.D., Jesica R. Jacobs, Ph.D., Nimalie D. Stone, M.D., Sujan C. Reddy, M.D., John A. Jernigan, M.D., Margaret A. Honein, Ph.D., Thomas A. Clark, M.D., and Jeffrey S. Duchin, M.D. Epidemiology of Covid-19 in a Long-Term Care Facility in King County, Washington. The New England Journal of Medicine. DOI: 10.1056/NEJMoa2005412

48. D.F. Gudbjartsson, A. Helgason, H. Jonsson, O.T. Magnusson, P. Melsted, G.L. Norddahl, J. Saemundsdottir, A. Sigurdsson, P. Sulem, A.B. Agustsdottir, B. Eiriksdottir, R. Fridriksdottir, E.E. Gardarsdottir, G. Georgsson, O.S. Gretarsdottir, K.R. Gudmundsson, T.R. Gunnarsdottir, A. Gylfason, H. Holm, B.O. Jensson, A. Jonasdottir, F. Jonsson, K.S. Josefsdottir, T. Kristjansson, D.N. Magnusdottir, L. le Roux, G. Sigmundsdottir, G. Sveinbjornsson, K.E. Sveinsdottir, M. Sveinsdottir, E.A. Thorarensen, B. Thorbjornsson, A. Löve, G. Masson, I. Jonsdottir, A.D. Möller, T. Gudnason, K.G. Kristinsson, U. Thorsteinsdottir, and K. Stefansson . Spread of SARS-CoV-2 in the Icelandic Population. New England Journal of Medicine. DOI: 10.1056/NEJMoa2006100

49. Michelle L. Holshue, M.P.H., Chas DeBolt, M.P.H., Scott Lindquist, M.D., Kathy H. Lofy, M.D., John Wiesman, Dr.P.H., Hollianne Bruce, M.P.H., Christopher Spitters, M.D., Keith Ericson, P.A.-C., Sara Wilkerson, M.N., Ahmet Tural, M.D., George Diaz, M.D., Amanda Cohn, M.D., LeAnne Fox, M.D., Anita Patel, Pharm.D., Susan I. Gerber, M.D., Lindsay Kim, M.D., Suxiang Tong, Ph.D., Xiaoyan Lu, M.S., Steve Lindstrom, Ph.D.,Mark A. Pallansch, Ph.D., William C. Weldon, Ph.D., Holly M. Biggs, M.D., Timothy M. Uyeki, M.D., and Satish K. Pillai, M.D. First Case of 2019 Novel Coronavirus in the United States. New England Journal of Medicine. DOI:10.1056/NEJMoa2001191

50. Pavan K. Bhatraju, M.D., Bijan J. Ghassemieh, M.D., Michelle Nichols, M.D., Richard Kim, M.D., Keith R. Jerome, M.D., Arun K. Nalla, Ph.D., Alexander L. Greninger, M.D., Sudhakar Pipavath, M.D., Mark M. Wurfel, M.D., Ph.D., Laura Evans, M.D., Patricia A. Kritek, M.D., T. Eoin West, M.D., M.P.H., Andrew Luks, M.D., Anthony Gerbino, M.D., Chris R. Dale, M.D.,Jason D. Goldman, M.D., Shane O'Mahony, M.D.,and Carmen Mikacenic, M.D. Covid-19 in Critically Ill Patients in the Seattle Region — Case Series New England Journal of Medicine. DOI: 10.1056/NEJMoa2004500

51. W. Guan, Z. Ni, Yu Hu, W. Liang, C. Ou, J. He, L. Liu, H. Shan, C. Lei, D.S.C. Hui, B. Du, L. Li, G. Zeng, K.-Y. Yuen, R. Chen, C. Tang, T. Wang, P. Chen, J. Xiang, S. Li, Jin-lin Wang, Z. Liang, Y. Peng, L. Wei, Y. Liu, Ya-hua Hu, P. Peng, Jian-ming Wang, J. Liu, Z. Chen, G. Li, Z. Zheng, S. Qiu, J. Luo, C. Ye, S. Zhu, and N. Zhong. Clinical Characteristics of Coronavirus Disease 2019 in China. New England Journal of Medicine. DOI: 10.1056/NEJMoa2002032

52. Arciuolo, R. J., Jablonski, R. R., Zucker, J. R., & Rosen, J. B. (2017). *Effectiveness of Measles Vaccination and Immune Globulin Post-Exposure*

*Prophylaxis in an Outbreak Setting—New York City, 2013. Clinical Infectious Diseases, 65(11), 1843–1847.* doi:10.1093/cid/cix639

53. Swamy, G. K., & Dotters-Katz, S. K. (2019). *Safety and varicella outcomes after varicella zoster immune globulin administration in pregnancy. American Journal of Obstetrics and Gynecology.* doi:10.1016/j.ajog.2019.07.003

54. Levin MJ, Duchon JM, Swamy GK, Gershon AA. Varicella zoster immune globulin (VARIZIG) administration up to 10 days after varicella exposure in pregnant women, immunocompromised participants, and infants: varicella outcomes and safety results from a large, open-label, expanded-access program. PLoS One 2019;14: e0217749

55. MMWR. Recommendation of the Immunization Practices Advisory Committee (ACIP) Postexposure Prophylaxis of Hepatitis B. https://www.cdc.gov/mmwr/preview/mmwrhtml/00022736.htm

67. Transfusion-associated circulatory overload (TACO) Definition (2018) IHN/ISBT haemovigilance working party/AABB

77. CDC. NHSN Biovigilance Component, Hemovigilance Module Surveillance Protocol v2.1.3. Atlanta: Centers for Disease Control and Prevention, 2014.
78. Simons FE, Ardusso LR, Bilo MB, et al, and the World Allergy Organization. World Allergy Organization anaphylaxis guidelines: summary. J Allergy Clin Immunol 2011; 127: 587, e1–22

79. Tinegate H, Birchall J, Gray A, et al, and the BCSH Blood Transfusion Task Force. Guideline on the investigation and management of acute transfusion reactions. Prepared by the BCSH Blood Transfusion Task Force. Br J Haematol 2012; 159: 143–53.

80. 15 Lin RY, Curry A, Pesola GR, et al. Improved outcomes in patients with acute allergic syndromes who are treated with combined H1 and H2 antagonists. Ann Emerg Med 2000; 36: 462–68

81. 16 Runge JW, Martinez JC, Caravati EM, Williamson SG, Hartsell SC. Histamine antagonists in the treatment of acute allergic reactions. Ann Emerg Med 1992; 21: 237–427

82. Fung M, Grossman BJ, Hillyer CD, Westhoff CM. Technical Manual, 18th edn. Glen Burnie, MD: AABB Press, 2014.

96. Normas Administrativas y Técnicas, RM 797/13 – 139/14 – 1507/15 Dirección de Sangre y Hemoderivados Ministerio de Salud de la Nación. http://www.msal.gob.ar/disahe/images/stories/pdf/normas-hemoterapia.pdf

97. Zou S, Dorsey KA, Notari EP, Foster GA, Krysztof DE, Musavi F, Dodd RY, Stramer SL. Prevalence, incidence, and residual risk of human immunodeficiency virus and hepatitis C virus infections among United States blood donors since the introduction of nucleic acid testing. Transfusion. 2010; 50:1495–504. [PubMed: 20345570]

98. Zou S, Stramer SL, Notari EP, Kuhns MC, Krysztof D, Musavi F, Fang CT, Dodd RY. Current incidence and residual risk of hepatitis B infection among blood donors in the United States. Transfusion. 2009; 49:1609–20. [PubMed: 19413732]
