## Supplementary material for "Prevention of severe COVID-19 in the elderly by early high-titer plasma": Translated Protocol

### EVALUATION OF THE EFFICACY OF COVID-19 CONVALESCENT PLASMA ADMINISTRATION IN REDUCING THE PROGRESSION TO SEVERE DISEASE IN OLDER ADULTS WITH MILD SYMPTOMS DUE TO SARS-COV2

**Hospital Central de San Isidro Principal Investigator:** Dr. Ramiro Larrea
**Dr. Carlos Bocalandro Hospital Principal Investigator:** Dr. Aníbal Rondan
**Principal Investigator Simply Evita Hospital:** Dra. Valeria Fernández Viña
**Evita Pueblo Hospital Principal Investigator:** Dra. Sandra Azcárate
**San Juan de Dios Hospital Principal Investigator:** Dra. Ivonne Ritou
**COVID- PAMI Centers Principal Investigator**: Dr. Gonzalo Perez Marc
**Principal Investigator of the Program**: Dr. Fernando P. Polack (INFANT)

**Co- Investigators:** Dra. Romina Libster (INFANT / CONICET), Dr. Gonzalo Pérez Marc (HMC), Dr. Diego Wappner (SMG), Dr. Jorge Lantos ( Los Arcos), Dr. Ricardo Valentini (CEMIC), Dr. Federico Etchenique (Ecco), Dr. Ma ximiliano De Zan (OSECAC), Dr. Gabriel Leberzstein (OSECAC) , Dr. Miguel González (Finochietto), Dr. Andrea Gamarnik (Leloir / CONICET), Dr. Jorge Geffner (UBA / CONICET ), Bioq. Silvina Coviello (INFANT), Dr. Mauricio Caballero (INFANT / CONICET) , Dr. Damián Alvarez Paggi (INFANT / CONICET), Dr. Sebastián Esperante (INFANT / CONICET) , Dr. Federico Dimase (HMC), Dr. Susana Pastor Argüello (HMC) , Dr. Juan Molinos (Olivos Clinic), Dr. Pablo Cruz (Galician Center), Dra. María Dolores Silveyra (Anchorena Sanatorium), Dr. Alfonso Raggio (Antarctica Sanatorium), Dr. Juan Sebastián Riera (Ministry of Health of the Province of Buenos Aires), Dr. Enio García (Ministry of Health of the Province of Buenos Aires), Dr. Juan Canela (Ministry of Health of the Province of Buenos Aires), Dr. Mario Rovere (Ministry of Health of the Province of Buenos Aires) , Dr. Fernando Althabe (WHO), Dr. Eduardo Bergel (IECS),

**Advisors:** Dr. Daniel Stamboulian (FUNCEI)

#### 1. INTRODUCTION

SARS-CoV2 is a new and particularly aggressive virus for elderly individuals, who represent between 73% and 90% of fatal cases in different regions of the world [28, 30, 40]. Mortality, the need for intensive care, mechanical ventilation, oxygen requirements, and hospitalizations for respiratory disease increase markedly with advancing age in patients [2, 3, 29, 32, 46]. In fact, the frequency of death from disease due to SARS-CoV2 (COVID-19) is 8% in patients between 70 and 80 years old, and increases to 14.8% in those over 80 [23]. Specifically in hospitalized patients older than 75 years, mortality is 29.4% [24], and in intensive care it amounts to an alarming 43.5% [25]. Several comorbidities associated with vascular and / or pulmonary disease worsen the prognosis of those infected even in patients younger than 70, including arterial hypertension, diabetes, obesity, and chronic obstructive pulmonary disease (COPD) [3,23,30,31]. To date, the virus has caused more than 290,000 deaths worldwide, more than 300 of them in Argentina which is contained through a transitional strategy quarantine quasi-universal [4]. There is no specific treatment available yet. The results of the efficacy trials for SARS-CoV2 vaccines will be known within a minimum of 10-14 months.

SARS-CoV2 is an enveloped virus that contains single stranded RNA in the positive sense bound to a nucleoprotein (protein N), within a capsid composed of matrix proteins (protein M). The envelope contains spine-shaped glycoproteins (protein S) that bind to the cellular receptor ACE2 in humans and generate neutralizing antibodies [13]. Primary SARS-CoV2 infection, like other respiratory diseases, generates an antibody response with early production of IgM followed by specific IgG against viral proteins [14]. Detection of virus neutralizing antibodies is common, reaching high levels of up to 1: 21,500 $PRNT_{50}$, mostly exceeding 1: 500 titers after 28 days of symptom onset in mild patients [41]. Approximately 5% of patients have neutralizing antibody levels <1:40, especially the younger ones [41]. A study of primary respiratory tract infection in rhesus macaques, using $10^6$ pfu of SARS-CoV2 followed by a second intratracheal challenge using the same virus and doses 28 days later, demonstrated the absence of reinfection after an extensive evaluation by RT-PCR in tissues post-necropsy [42]. Neutralizing titers before the second inoculum ranged from 1:8-1:16 $PRNT_{50}$ [42]. **These findings - limited indeed, given the novelty of the problem - suggest that even low neutralizing antibody titers could prevent severe disease due to the virus.**

Protecting the populations most vulnerable to SARS-CoV2 is imperative, in the context of a highly infectious pathogen ($R_0$ estimated between 2 and 3.28) [27, 33, 34]. Its contagiousness rapidly overwhelms the health capacity of developed countries in different regions of the world [5] and threatens the health resources of developing countries [6]. Regions of Italy, Spain and the United States have been overwhelmed by the pandemic, which represents a serious threat to Argentina, coinciding with the beginning of the annual circulation of other lung pathogens, such as the common flu and respiratory syncytial virus [26]. Obviously, it would be impossible to respond to an avalanche of critically ill patients [7], and any strategy aimed at reducing infections and the development of severe disease **is directly aligned with the medical objective of saving lives and with the health needs of the country in order to provide the best possible care for those patients who need it**.

Post-exposure prophylaxis to viruses and treatment in early stages of disease, through defenses obtained in convalescent plasma, is a widespread practice with successful antecedents in the history of medicine at a national and global level. As early as 1960, Brunell et al. managed to prevent clinical varicella in siblings with sick parents, using anti-varicella zoster immunoglobulins obtained from the plasma of previously infected patients [8]. Today, for example, enriched immunoglobulin for measles is used in unvaccinated people who were in contact with a patient for the previous 6 days with an efficacy close to 100% [9, 52]; a similar product enriched for varicella zoster virus is administered to unvaccinated pregnant women, newborns, and immunosuppressed patients in contact with infected persons within 96 hours, reducing the incidence of severe disease by 90% [10, 53, 54]; tetanus immunoglobulin supplements vaccination in soiled wound victims with incomplete primary immunization [43]; gamma globulin against hepatitis B is used as prevention for newborns of infected mothers with an efficacy of 75% [44, 55]; and rabies gamma globulin is ~ 100% effective in the prophylaxis of post-injury due to a rabid animal [11,56]. **Interventions using human immunity derived from convalescent plasma have been shown to be safe, have prevented countless cases of severe disease, and have preserved lives around the world.**

The main objective of our study is to evaluate the efficacy of convalescent plasma in reducing the progression to severe disease in people between 65-74 years with at least one comorbidity and in all those ≥75 years of age who present with mild symptoms of less than 48 hs of evolution at the time of screening and with an early diagnosis of COVID-19. *Our central hypothesis is that a single dose convalescent plasma compared against placebo (saline at 0.9%) administered up to 72 hours after the onset of symptoms by SARS-COV2, prevent progression to severe respiratory disease in subjects 65-74 years with at least one comorbidity and in ≥ 75 years irrespective of the presence of comorbidities .*

#### 2. OBJECTIVES OF THE STUDY

##### 2.1. Primary objective

To evaluate the efficacy of convalescent plasma , starting 12 hours after administration, in reducing the progression to severe respiratory disease in people between 65-74 years with at least one comorbidity and in all those ≥75 years with mild symptoms and a diagnosis of early COVID-19 until 15 days of administration of treatment (16-18 days of disease considering the pre-symptomatic period before enrollment).

##### 2.2. Secondary Objectives

The Secondary Objectives include determining if the administration of convalescent plasma up to 15 days or thereafter up to a maximum of *25 days of the administration of treatment in those still hospitalized*:

- Reduces the need for oxygen support with maximal oxygen therapy ($O_2$ reservoir mask) and/or use of non-invasive respiratory support (NIV support including CPAP) and/or admission to ICU and/or requirement of invasive mechanical ventilation due to SARS-CoV2 in patients between 65-74 years with at least one comorbidity and in all those ≥75 years.
- Reduces critical illness defined as (a) presence of acute respiratory failure (PaO2 / FiO2 ≤ 200 mm Hg.) and / or (b) shock (defined by need of vasoactive drugs to maintain MAP equal to or greater than 65 mm Hg) , and / or (c) multi-organ dysfunction syndrome (MODS ): acute kidney injury (defined by a creatinine increase two or more times from baseline or a creatinine increase of 0.3 mg / dl) , elevated liver enzymes (transaminases) greater than three times the upper normal value , acute cardiomyopathy (defined by troponin elevation above the normal level and/or new electrocardiographic or echocardiographic changes of myocardial damage) in patients 65-74 years with at least one comorbidity and in all those ≥ 75 years due to COVID-19.
- Reduces mortality due to SARS-CoV2 in patients between 65-74 years with at least one comorbidity and in all those ≥ 75 years due to COVID-19.
- Decreases the length of oxygen support due to hypoxemia associated with COVID-19.
- Describe the safety of convalescent plasma administration in patients between 65-74 years with at least one comorbidity and in all those ≥75 years with COVID-19.
- Explore the concentration of IgG anti-S SARS COV2 in plasma of participants associated with the absence of severe respiratory disease obtained at 24 hours of infusion.

##### 2.3. Exploratory Objectives

- Explore whether, rather than protect against the severe respiratory disease, administration of convalescent plasma delays the onset of the disease.
- Explore the effects of convalescent plasma on primary humoral immune response against infection by SARS COV2 at 90 days post-illness.

- Explore whether the effects on respiratory and cardiovascular disease and long-term mortality were affected by the intervention at 12 months of enrollment.
- Explore the correlation between viral load determined by RT-PCR at diagnosis and the response to plasma treatment.

#### 3. EFFICACY ASSESSMENT CRITERIA

##### 3.1.1. Primary Efficacy Assessment Criterion

Severe respiratory disease due to SARS-CoV2, confirmed by detection of viral RNA by RT-PCR, and defined by the presence of any of the following two variables in a non-exclusive way: (a) respiratory rate ≥30 per minute, (b) oxygen saturation in room air <93% [20,21]. The primary endpoint will be determined from 12 hours of the start of the infusion until 15 days since administering treatment.

##### 3.1.2. Secondary Efficacy Assessment Criteria *(up to 15 days or, thereafter, up to a maximum of 2 5 days after the administration of treatment if continued hospitalization)*

- Need for oxygen support with maximum oxygen therapy ($O_2$ reservoir mask) and/or non-invasive respiratory support (NIV support including CPAP) and/or admission to ICU and/or requirement of invasive mechanical ventilation, due to SARS-CoV2 .
- Critical illness defined as (a) presence of acute respiratory failure (PaO2 / $FiO_2$ ≤ 200 mmHg.) and/or (b) shock, and/or (c) multi-organ dysfunction syndrome (MODS) according to criteria detailed in section 2.2: acute kidney injury, elevated liver enzymes, acute cardiomyopathy, due to SARS- Cov2.
- Mortality due to COVID-19.
- Combination of the secondary efficacy endpoints # 1 (need for oxygen support with…) and / or # 2 (defined critical illness…) and / or # 3 (mortality from covid-19).
- Duration of oxygen requirement in patients with COVID-19 due to $O_2$ saturation in room air <93%.

#### 4. HYPOTHESIS AND JUSTIFICATION

##### 4.1. Hypothesis

Our central hypothesis is that a convalescent plasma dose compared to placebo, and administered up to 72 hours from the onset of mild symptoms, will prevent progression to severe respiratory disease due to covid-19 in patients between 65-74 years with at least one comorbidity and in all those ≥75 years who present with less than 48 hours of evolution and with mild symptoms.

##### 4.2. Justification

The efficacy of an economically accessible therapeutic preparation against SARS-CoV2 analyzed by means of a randomized, double-blind study in vulnerable patients would allow its applicability to be immediately scaled up nationally and internationally through massive plasma donation campaigns in convalescent patients awaiting definitive solutions such as successful drugs, monoclonal antibodies, or vaccines [17]. Even if there is concentrated partial protection in the lower respiratory tract [18], *this same strategy could hypothetically allow immunization of subjects through controlled replication of the germ in the upper respiratory tract* (as in the case of passive administration of antibodies against respiratory syncytial virus in infants [19]).

#### 5. STUDY DESIGN

A randomized, double-blind, placebo-controlled study to test the efficacy of convalescent plasma, administered up to 72 hours after the onset of symptoms, to prevent progression to severe respiratory disease. The study will be conducted in subjects aged 65-74 years at the least one comorbidity and all ≥ 75 years who presented with mild symptoms of less than 48 hours at the time of screening (as defined in section 5.1) and early diagnosis of COVID-19. Subjects will be followed for a minimum observation period of 15 days in which participants will remain under medical supervision daily to determine the primary endpoint. Monitoring will extend thereafter, up to 25 days if subjects remain symptomatic and hospitalized, to analyze secondary endpoints.

The study is projected to incorporate an estimated maximum of 210 patients (see 9.0 Sample size). Participants will be randomized in a 1: 1 ratio.

**Table 1. Treatment Assignments**

| Treatment Group | Range of subjects assigned by branch | Test Article | Intravenous Dosing Volume | Intervention Administration Day |
|---|---|---|---|---|
| TO | 1 05 | Placebo ( 0.9% saline solution ) | 25 0 cc | Day 0 |
| B | 1 05 | Convalescent plasma | | |

All participants will receive a single intravenous dose (EV) - according to the ADMINISTRATIVE AND TECHNICAL REGULATIONS RM 797/13 - 139/14 - 1507/15

Directorate of Blood and Blood Products Ministry of Health of the Nation (at http: // www. msal.gob.ar/disahe/images/stories/pdf/normas-hemoterapia.pdf ) on Day 0 with the assigned test article, convalescent plasma or placebo (see Table 1). For each subject, participation in the study will last a minimum of 15 days or until the resolution of symptoms from the administration of the test article with a maximum of 25 days. Participating subjects will be monitored during and after administration of the test article to assess expected and unexpected safety events after plasma administration. A Data and Safety Monitoring Board will oversee the enrollment, efficacy, and safety of the participants throughout the study.

The study will be conducted at the San Juan de Dios Hospital, the Simply Evita Hospital, the Dr. Carlos Bocalandro Hospital , the Evita Pueblo Hospital, the San Isidro Central Hospital and the Antarctic Sanatorium of the Province of Buenos Aires in close coordination with health effectors of the provincial Ministry of Health and the Hospital Central Militar, the Sanatorio of Los Arcos, CEMIC, the Research Center OSECAC , the Sanatorio Finochietto , the Galician Center of Buenos Aires, the Sanatorio Anchorena and Olivos Clinic and will not interfere with the care and standards of patient care established for notification and management of this disease.

**MOBILE-COVID TEAM.** Given the context of the COVID-19 situation in the AMBA with the reduction in the availability of beds and the overload of hospital staff added to the exponential increase in severe cases, mildly symptomatic patients without oxygen requirements and from nursing homes find it extremely difficult (almost impossible) to be admitted to the hospitals. Therefore, to facilitate the incorporation of the main potential beneficiaries of this intervention to the study (that is, the most vulnerable elderly) and given the enormous existing public health need to provide therapeutic alternatives against this scourge, a COVID-Mobile unit will be added to monitor volunteers in COVID-PAMI centers where the administration of the investigational product (now clearly demonstrated as safe in publications in thousands of patients) are performed to the same standards and the same equipment as in all other research sites. The COVID-Mobile team will record the evolution of these participants who are under the care of doctors and health personnel from PAMI *on site*. Any transfer of these patients to centers of greater complexity due to the natural evolution of their disease will be the decision of the treating physicians of the PAMI, and the mobile research team will limit itself to continuing to collect data on their clinical progress in the same way as it does today when a patient is transferred or receives an early discharge from another institution.

##### 5.1. Eligibility criteria

###### 5.1.1 Inclusion Criteria:

A subject must meet the following criteria to be included in the study:

1. Age ≥ 75 years or age between 65-74 years with at least one of the following comorbidities:
   a. Diagnosis of arterial hypertension under pharmacological treatment.
   b. Known diagnosis of diabetes in treatment with one or more of the available drugs (Appendix I).
   c. Obesity (BMI -Body mass index- ≥ 30 kg / m2),
   d. Chronic obstructive pulmonary disease (COPD) treated with any of the available drugs (Appendix I).
   e. Cardiovascular disease defined as a) known diagnosis of coronary heart disease, b) history of ischemic stroke or hemorrhagic, or c) ICC (defined as FE < 40%) .
   f. Chronic kidney disease (defined as a reduction in GFR below 60 ml/min/1.73 $m^2$).
2. Present for less than 48 hs : ( A ) axillary temperature ≥ 37,5°C or fever equivalence (defined as feeling cold and/or chills and/or unexplained sweating) , combined with (a ) dry cough and / or ( b) difficulty breathing and / or ( c) odynophagia and / or (d) anosmia and/or (e) dysgeusia and / or (e) any of the following symptoms : fatigue, anorexia, myalgia or rhinorrhea .
3. SARS-CoV2 diagnosis confirmed by RT-PCR
4. Being able, according to medical judgment, to understand and comply with the study procedures and to consent or not to their participation.
5. Provide informed consent.
6. Patients with cognitive vulnerability with support in decision-making by their representative, can be included as detailed below. (see section 12)

##### 5.1.2 Exclusion Criteria

A subject who meets any of the following criteria will be excluded from the study:

1. Present at the time of inclusion SEVERE RESPIRATORY DISEASE defined as (a) respiratory rate ≥30 per minute and / or (b) oxygen saturation in ambient air <93%.
2. HEART FAILURE in functional class III and IV (according to the functional classification of the New York Heart Association).
3. Known diagnosis of CHRONIC KIDNEY INSUFFICIENCY in filtration stages G4 and G5 (according to KDIGO classification).
4. Previous diagnosis of PRIMARY HYPOGAMMAGLOBULINEMIA (congenital and hereditary).
5. Previous diagnosis of MONOCLONAL GAMMOPATHIES (multiple myeloma, Waldenström macroglobulinemia, primary amyloidosis, heavy chain disease, among others).
6. Previous diagnosis of SELECTIVE DEFICIENCY OF IgA.
7. Previous diagnosis of MYELODYSPLASIC SYNDROMES. [WHO classification: refractory thrombocytopenia (TR), refractory anemia (RA), etc.]

8. Previous diagnosis of CHRONIC LYMPHOPROLIFERATIVE SYNDROMES. (WHO classification: peripheral B lymphocyte lymphomas, T lymphocyte and NK cell lymphomas, Hodgkin lymphoma).
9. KNOWN HYPERSENSITIVITY to the administration of immunoglobulins, plasma, monoclonal antibodies and / or vaccines.
10. Active cancer, defined as receiving or having received or receiving chemotherapy, radiotherapy, or new designer or monoclonal drug treatments in the last six months.
11. Known HIV, HBV, or HCV infection.
12. Chronic administration (defined as more than 14 calendar days) of immunosuppressants or other drugs that modify the immune system **at the time of enrollment** or within the 6 months prior to administration of study medication. An immunosuppressive dose of glucocorticoids will be defined as a systemic dose of ≥10mg of prednisone per day or its equivalent.
13. Solid organ transplant.
14. Known chronic liver disease diagnosed with stage II-III-IV cirrhosis.
15. Chronic lung disease with oxygen requirement.
16. Any other physical, psychiatric or social condition that may, at the discretion of the investigator, increase the risks of participation in the study for the participant or that may lead to the collection of incomplete or inaccurate safety data.
17. Patients who are receiving any type of anticoagulant therapy orally: family of dicoumarins (Warfarin such as Acenocoumarol) or direct acting anticoagulants (DOAC) (Rivaroxaban, Dabigatran, Apixaban) as well as those who are receiving heparin in any of its types for anticoagulant therapy purposes.

##### 5.2. Study procedures (Table 2)

##### 5.2.a. Participating population and identification process of potential subjects.

Subjects 65-74 years with at least one comorbidity and all ≥ 75 years who present for less than 48 hours the following symptoms: ( **a** ) temperature ≥ 37,5°C or fever equivalence (defined by feeling cold or chills or unexplained sweating ) , combined with ( **b** ) dry cough and / or dyspnea and / or anosmia / dysgeusia and / or odynophagia and / or fatigue and / or anorexia and / or myalgias and / or rhinorrhea .

Those patients who meet the inclusion criteria and do not meet any of the aforementioned exclusion criteria will be invited to participate in the research phase of the study in which, if they accept (expressing their agreement by signing the informed consent - research ANNEX), a sample of respiratory secretions is obtained by swabbing the nasopharynx and oropharynx (according to the procedure and regulations of the Argentinean Ministry of Health) to assay for RNA of SARS-COV2 by reverse transcriptase-polymerase chain reaction (RT-PCR).

The sample will be processed and analyzed by the laboratory designated for participating institutions daily, using the same RT-PCR test to assess the presence of RNA of SARS-COV2. This determination will be made as soon as possible (within a maximum of 24 hours) and under all the biosecurity standards indicated by the health authorities. The samples will be kept for later testing by RT-PCR for co-infections with influenza A and B, respiratory syncytial virus and other winter viral pathogens at the INFANT Foundation.

Patients who already have a positive result ( + ) considered valid by MOH standards will be invited to participate in the research phase of the study and will not be swabbed again .

##### 5. 2 .b. Enrollment for participation in the study

If SARS-Co V 2 RNA is detected in the respiratory sample, the candidate will be informed about the details of the study and eligibility will be verified again. The candidates may decide not to participate in the investigation by their free will and without any consequence whatsoever. In the event that the subject is interested in participating in the study, and after their consent is attested with the signing of the informed consent for participation (ANNEX), a unique identification number will be assigned. Once its alphanumeric code has been assigned, vital signs will be obtained and a complete physical examination will be performed to obtain the following demographic and clinical information [1-3, 25, 47-51]:

- Sex
- Age
- Usual medications and those of the last 15 days.
- History of smoking
- Comorbidities of primary interest: arterial hypertension, diabetes, coronary heart disease, heart failure, history of stroke and AMI, COPD, chronic kidney disease, and obesity.
- Secondary comorbidities: asthma, history of cancer, liver diseases, and neurological diseases.

##### 5.2.c. Random assignment

All participants with confirmed eligibility will be randomly assigned to a treatment group. The randomization of participating subjects will be conducted by the person in charge of randomizing the study using an electronic system. Once the patient is randomized, the person in charge of the allocation will communicate with a member of the Transfusion Medicine Central Team in the study who will remove the opaque bag treatment indicated in the freezer of the Central Laboratory. The Transfusion Medicine team will proceed to take the cold bag to the hospital. There, he/she will complete the preparation of the masked product and administer the indicated treatment to the participating patient, using a venous access previously obtained by a member of the hospital team for the infusion. Only the person responsible for randomization and the physician in the Transfusion Medicine Central Team administering treatment will know the product administered to each patient (unblind). Neither the patient nor the researchers will have knowledge of the administered treatment, conducting a double-blind study. The unblind team will keep a record of the product assignments and their administrations that will be kept confidential. In the eventual need to open the blind, such records may be used.

**5. 2 .d. Preparation and administration of the test article**

The preparation and administration of the convalescent plasma or placebo (physiological solution) will be performed according to the ADMINISTRATIVE AND TECHNICAL REGULATIONS RM 797/13 - 139/14 - 1507/15 Directorate of Blood and Hemoderivatives Ministry of Health of the Nation. ( http://www.msal.gob.ar/disahe/images/stories/pdf/normas-hemoterapia.pdf ).

Once masked, the team of Transfusion Medicine will administered the product intravenously for 1.5 hours and up to 2 hours. After administration, local reactions at the injection site and systemic reactions will be monitored for 12 hours. *This is a minimal risk study, since the administration of plasma and saline (placebo in this study) are a daily habit in medical practice.* In the event of a medical emergency, when it is known that the treatment assignment may influence the medical care of the patient, the investigator or designee may request that the subject be unblinded for the emergency. However, prior to breaking the blind the investigator must make all reasonable efforts to discuss the decision to break the blind. The investigator is expected to provide a reason for the need to break the blind, based on a significant change in the participant's immediate or short-term medical care that will result from knowledge of the treatment assignment.

**5. 2 .e. Taking a blood sample for titration of IgG anti-S SARS CoV2 in serum.**

At 24 hours of completion of infusion of the treatment, a sample of 5 ml of venous blood will be obtained from all participants for titration of serum IgG anti-S SARS-COV2 and held at the Foundation INFANT. The assay will be performed by the ELISA method (COVIDAR IgG, Leloir / CONICET and Genetech IgG and sNT). Serum will be preserved to -20 °C and transferred by a shuttle authorized for sample handling system transport biological the Foundation.

Window for sample collection: +/- 4 hours. The sample may be taken within a minimum period of 20 hours and a maximum of 28 hours after the administration of the Research Product is complete.

**5. 2. f. Daily clinical monitoring**

We will perform one control daily from Day 0 and at least 15 days from the infusion and up to 25 days for those subjects who remain hospitalized with symptoms after day 15. Patients who are discharged prior to the day 15 will be monitored at home by a team of physicians trained for this purpose working in collaboration with the research centers. The following clinical information will be collected daily (using a questionnaire designed for this purpose) by study personnel and **without interfering with the patient's medical care or altering the standards recommended by the health care**

**authorities for the management of patients with COVID-19** : (a) respiratory rate (looking at the chest excursion for 30 seconds) , (b) oxygen saturation, (c) temperature, (d) central heart rate , (e) cough and duration, (f) sore throat and duration , (g) feeling of fever / chills, (h) need for supplemental oxygen provision and duration, (i) need for intensive care and duration, (j) need for non-invasive or invasive respiratory support and duration, (k) evidence of SDMO : according to criteria for acute kidney injury , criteria for elevated liver enzymes and criteria for acute cardiomyopathy (detailed in section 2.2) , (l) survival.

**Table 2. Schedule of study procedures.**

| **Study Day** | **-1 Research** | **0 Day 0** | **Daily follow up** |
|---|---|---|---|
| **Informed consent for the evaluation of history, clinical condition and *SARS-COV2 diagnosis*** | X | | |
| **Nasopharyngeal swab collection for confirmation of SARS-CoV2 by RT-PCR** | X [1] | | |
| **Informed consent to participate in the study** | | X | |
| **Medical history review** | | X | X |
| **Vital signs** | | X | |
| **Eligibility confirmation** | | X | |
| **Concomitant medications** | | X | X |
| **Random assignment of treatment** | | X | |
| **Administration of the intervention** | | X | |
| **Monitoring of transfusion** | | X | |
| **Surveillance of adverse events** | | X * | (X) |
| **Taking a blood sample after the infusion is complete** | | | 24 hours (+/- 4 hours) |
| **Surveillance of respiratory symptoms** | | X | X |

(1) This procedure will not be required if a result is available by a valid method at the time of patient screening.

We consider that the patient completes its participation when reaching day 15 (if the patient is already discharged) , at the time of discharge between days 15 and 25, or at the expiration of day 25 if he/she continues hospitalized.

#### 6. Data Safety Monitoring Board

A Data Safety Monitoring Board (DSMB) will monitor the safety of the subjects throughout the study. The DSMB includes at least experienced members in managing internal medicine, infectious diseases, and a biostatistician with specific experience in the design, analysis and safety of clinical trials. The DSMB will operate under an approved plan and will have the responsibility to monitor the outcomes/end points, adverse events (AE) and serious adverse events (SAE) , and recommend termination of the study if it appears at any point during the study that participants (or a subset of participants) are at undue risk as a result of their participation. The DSMB will meet (by teleconference) weekly to review the accumulated data on safety and efficacy and will conduct an interim analysis of the data. The DSMB will issue written recommendations based on its meetings regarding the continuity of the study. The minutes of each meeting will be recorded in documented minutes based on a pre-established work schedule. Any participant-specific protected health information reviewed by the DSMB will be kept completely confidential. Sessions will be closed without access to third parties.

#### 7. Security Monitoring

The researcher at each institution and at a general level will supervise the safety of the study patient at his/her site(s) according to the requirements of this protocol and in accordance with current Good Clinical Practices (GCP). The investigator will monitor safety data at all study sites. Security monitoring will be carried out on an ongoing basis (for example, individual review of EA S, EAs and endpoints) and on a periodic cumulative basis.

##### Adverse event (AE)

An AE is any unfavorable medical event in a patient who was administered a study drug that may or may not have a causal relationship to the study drug. Therefore, an AE is any unfavorable and unintended sign (including abnormal laboratory finding), symptom, or disease that is temporarily associated with the use of a study drug, regardless of whether it is considered related to the study drug.

An AE also includes any worsening (ie, any clinically significant change in frequency and / or intensity) of a pre-existing condition that is temporarily associated with the use of the investigational product.

Covid-19 progression will not be considered an AE if it is clearly consistent with the typical progression pattern of the disease

If there is any uncertainty that an AE is due to covid-19 progression only, it will be reported as an AE or SAE, as described in the appropriate protocol section.

**Serious adverse event (SAE)**

A SAE, by definition, is any unfavorable medical event that at any dose:

- Results in death: Includes all deaths, even those that appear to be unrelated to the study drug (for example, a car accident in which a patient is a passenger).
- It is life threatening: In the opinion of the investigator, the patient is at immediate risk of death at the time of the event. This does not include an AE which, if it had occurred in a more severe form, could have caused death.
- Requires hospitalization or extension of existing hospitalization. Hospitalization is defined as admission to a hospital or emergency room for more than 24 hours. Prolongation of existing hospitalization is defined as a hospital stay that is longer than originally planned for the event, or prolonged due to the development of a new AE as determined by the investigator or treating physician.
- Results in persistent or significant disability (substantial disruption of one's ability to carry out normal life functions).
- It is a major medical event - Major medical events may not immediately endanger life or result in death or hospitalization, but may endanger the patient or may require intervention to prevent one of the other serious outcomes listed above (for example, intensive treatment in an emergency room or at home for allergic bronchospasm; blood dyscrasias that do not result in hospitalization).

In the case specific of this study we will consider AE or SAE , according to the detailed description in section 11.2, those that are initiated within 12 hours after administration of the IP and clinical exacerbations and deaths associated with these events. We will not consider as AE and SAEs those events occurring after 12 hours of initiation of treatment, that are consistent with the clinical course in patients infected with SARS COV2.

**Adverse Event Collection Period**

The reporting period for AE/SAE begins when the participant was initially included in the study (date of signature of informed consent for participation) and receives the infusion and continues for events that occur in the next 12 hours (see section 11.2 ) . During the follow up period, after the initial 12 hours of IP administration subjects will be monitored for the clinical outcome associated with primary and secondary endpoints.

#### 8. Leaving the Study Prematurely

Subjects may leave the study at any time if they wish, but we will make every effort to monitor them during the stipulated period to guarantee their safety .

#### 9. Patient Replacement

Those patients who decide to drop out of the study prior to drug administration will be replaced, if necessary, to ensure an adequate number of evaluable patients.

#### 10. Justification of the sample size and statistical analysis

There is significant uncertainty in the expected effect size of the intervention, and considering that the trial is expected to be completed in a relatively short period of time, the study is designed to have an interim analysis when the results of 50% of the subjects have been acquired.

Given the relative complexity of implementing this intervention, the minimally clinically important difference is established in a relative reduction of 40%, for an expected result rate of 50% in the control group that is reduced to 30% in the intervention group. A total sample size of 210 subjects (105 per test arm) will have 80% power, at a significance level (alpha) of 0.05 using a two-sided z-test with continuity correction. These results assume that 2 sequential tests are performed using the O'Brien-Fleming expenditure function to determine the limits of the test, as described in the table below.

| | | Lower | Upper | Nominal | |
|---|---|---|---|---|---|
| **Look** | **Time** | **Bndry** | **Bndry** | **Alpha** | **Power** |
| 1 | 0.50 | -2.96259 | 2.96259 | 0.003 | 0.168 |
| 2 | 1.00 | -1.96857 | 1.96857 | 0.049 | 0.806 |

Under the primary analysis strategy, we will use the Kaplan-Meier product cutoff distribution to compare treatment groups over time required to reach the primary outcome. An estimate of the relative risk and the 95% confidence interval will also be reported.

Based on a recommendation of the DSMB, we will also conduct an analysis stratified by age (65-74 years and ≥75 years) and adjust the statistical analysis by those significant risk factors whose distribution between groups can affect the outcome.

#### 11. Test article and transfusion medicine

##### 11.1. Voluntary donation of convalescent plasma

All procedures will be done in accordance with the STRATEGIC PLAN TO REGULATE THE USE OF PLASMA BY COVID-19 RECOVERED PATIENTS FOR THERAPEUTIC PURPOSES. (IF-2020-26315442-APN-SCS # MS)

https://www.boletinoficial.gob.ar/detalleAviso/primera/227976/20200418

Convalescent plasma will be obtained by inviting patients who experienced COVID-19 and have recovered satisfactorily in the Autonomous City and Province of Buenos Aires to participate as a voluntary donor, following the donation criteria established by the authorities. If the patient wishes to donate, they will voluntarily and altruistically sign the plasma donation consent (ANNEX).

Donors will be identified by the ministerial and institutional authorities (if it is a private body), contacted, and invited to donate in one of the five Blood Banks participating in this study:

a) Institute of Hemotherapy of the Province of La Plata
b) Central Military Hospital
c) Hemocentro Buenos Aires Foundation
d) Sarmiento Hematology Foundation
e) CEMIC

In order to donate plasma, the patient must comply with the conditions determined to date ( https://www.boletinoficial.gob.ar/detalleAviso/primera/227976/20200418 ) or follow updates from the Ministry of Health.

**Anti-SARS Cov2 antibody titration**

After donating plasma by apheresis or hemodonation, in addition to the samples obtained for routine blood bank controls, a dry tube of 5 mL of whole blood will be obtained, labeled with the identification given to the donor, identification of the Blood Bank and label that identifies the purpose of the "PCC19" study to perform the antibody titer. These will be asaayed using the SARS CoV2 anti-protein S ELISA test, which in light of the existing data on protection against the virus allows an indirect extrapolation of the serum's neutralization capacity, optimized by the Leloir Institute in charge of Dr. Gamarnik, and currently used by health teams in Argentina to study immunity prevalence. Likewise, if possible, data will be correlated with the neutralizing antibody titer against a pseudovirus encoding protein S optimized by the same laboratory.

Tubes will be stored in a designated location within the Bank and shipped in an approved biological sample shipment to the INFANT Foundation (over 18 years of experience in testing) for assaying ELISAs and neutralizing antibodies.

The titers obtained by S ELISA will then be classified and those that are above 1: 1,000 selected to provide convalescent plasma for this study.

The additional high titer plasmas will also be stored to be sent to the Córdoba Blood Products Center (UNC), which will produce gammaglobulin enriched with anti-SARS-CoV 2 antibodies.

**Plasmapheresis (Institute of Hemotherapy of the Province in La Plata, Central Military Hospital, Hemocentro Buenos Aires Foundation, and Sarmiento Hematology Foundation)**

Plasma donation will be conducted through an apheresis procedure with equipment and disposable supplies approved for this purpose, during which it is recommended to extract a volume no greater than 15% of the donor's blood volume. The anticoagulant to be used will be ACD-A or similar. In cases where volume replacement is not performed, plasma extraction should not exceed 600 ml per procedure.
The 48 hours interval must be respected between each procedure, not to exceed 2 donations in a week or 24 donations in a 12-month period.
The PCC19 units can be separated into 250ml aliquots, respecting the closed circuit and following the indications of the current standard operating procedure.

**Hemodonation (CEMIC)**

1-Plasma will be obatined with a Cobe Spectra equipment. 1.5 to 2 L of plasma will be extracted according to the donor's weight and tolerance to the procedure. In case of venous inaccessibility or opposition from the donor, between 400 and 500 ml of whole blood will be drawn (classic hemodonation)
2-The volume replacement will use NS and albumin solution.
3-During the procedure the plasma will be transferred to transfer bags of similar volume each (approximately 500ml).
4-The bags will be labeled as detailed below.
5-They will be frozen in the -56°C Righi vertical ultrafreezer (labeled Freezer # I, identified with COVID-19 convalescent plasma label in the Blood Bank) separated on shelves by bag type (1, 2 or 3).
6-The convalescent plasma will remain stored until its use or expiration (1 year), with appropriate safety criteria.
7-Disposal will be conducted under stipulated security regulations.
8- Stored aliquots of serum / plasma frozen for future determinations at -80°C in Eppendorf tubes.

**Labelled**
All units of PC COVID-19 must comply with the labeling required for fresh frozen plasma from each Blood Bank and must also have a clear identification, which must include:

- "Product Type: PC COVID-19"
- "Result of antibody titer for SARS-CoV-2" (if available)
- "CAUTION: product for research use only"
- "PROTOCOL: INFANT Foundation - Serum enriched with anti SARS-CoV2 antibodies"

**Cold Chain of PCCOVID-19**

1. The units are stored properly separated from the remaining plasma units and labelled for transfusional use in a place clearly identified for this purpose.

2. The PCC19 units will be stored at temperatures below -25°C, which allows a storage of 36 months and between -18°C and -25°C for 3 months of storage.

3. The units will be transported in a way to maintain the cold chain reliably.

4. Plasma units should be stored between 2 ºC and 6ºC until 24 hours after thawing. After then, they should be discarded.

**Traceability**

The Blood Bank and the Transfusion Service of the requesting Institution will implement a registry system that guarantees traceability between donors and recipients. In this special situation, they must also have a specific registry as this is a transfusion therapy administered in an experimental context.

#### 11.2. Administration of the intervention

After assessing vital signs and verifying elegibility, the unblinded team from the Medical Transfusion Center will be notified of the result of the randomization process and drive to the assigned hospital with the masked treatment product cold. On arrival, they will proceed again to check the code assigned for treatment with a member of the laboratory of Transfusion Medicine of the hospital (see 5.2.c).

Placebo will be a 250 ml of normal saline (0.9%) sterile, masked and administered in the same way and speed as plasma. To avoid bias and maintain the study blinded, plasma and saline 0.9% will be masked using an opaque bag and tape that will not allow to differentiate both products. Members of the Medical Transfusion Central Team will conduct the intervention with a member of the local team. Thus, both members of the Transfusion Medicine Central Team and the respective member of Transfusion Medicine at the hospitals will not be blind, while the rest of the team with delegated functions in the studies in each Center and the patients will be blind to the intervention.

To prepare the plasma, 250 ml bags will be thawed at 37 ° C, according to the operating procedures of the Department of Transfusion Medicine at the Hospital. Plasma will be infused slowly (at least 1.5 hours and up to 2 hours) according to the hemodynamics of the patient and monitored for 12 hours and adverse effects early and late attributable to transfusion recorded.

Night window for the passage of the research product: between 8:00 p.m. and 8:00 a.m., a window of +8 hours will be tolerated for the passage of the research product. A maximum of 80 hours (72 + 8) from onset of symptoms will be allowed in these cases before starting the infusion. This exception will only be allowed in this night time.

**Monitoring of adverse events associated with plasma transfusion**

***Transfusion-associated circulatory overload (TACO)***

Transfusion-associated circulatory overload (TACO) is the most common pulmonary complication and is an independent risk factor for hospital morbidity and mortality, with a higher incidence in critically ill patients [58]. The estimated frequency of TACO varies from 1% to 5% depending on the hemovigilance system [59], up to 8% in advanced post-surgical patients, and 11% in critically ill patients [60 61 62]. Risk factors include heart, lung, or kidney disease, age ≥70 years, and a positive, pre-transfusion fluid balance [63-64]. Most cases of TACO are prevented by slowing the transfusion rate and establishing the usual measures for fluid overload in susceptible populations. Overload constitutes a requested AE, but if it does not improve within 120 minutes after the infusion or requires - in the opinion of the responsible professionals - transfer of the patient to intensive care due to poor clinical progression, should be reported immediately as a SAE.

*Diagnostic criteria for TACO* [19]
During or up to 12 hours after the transfusion, TACO is characterized by the presence of a total of 3 or more of these criteria:
A. Acute or worsening respiratory compromise
B. Evidence of acute or worsening pulmonary edema based on clinical and / or chest radiographic images and / or other noninvasive assessment of cardiac function (echocardiogram)
C. Evidence of cardiovascular changes not explained by the patient's underlying medical condition, including: tachycardia, hypertension, widened pulse pressure, jugular venous distention, enlarged cardiac silhouette, and/or peripheral edema.
D. Evidence of fluid overload, including any of the following: a positive fluid balance; response to diuretic therapy, or dialysis combined with clinical improvement; and variation of the patient's weight.
E. Increased level of B-type natriuretic peptide (eg, BNP or NT-pro BNP) above the age-specific reference range and greater than 1.5 times the pre-transfusion value.

***Transfusion Associated Acute Pulmonary Injury (TRALI)***
This is an acute respiratory distress syndrome, it will be considered a SAE and it occurs during or within six hours after the administration of the transfusion. Prospective studies of TRALI reported very low incidence rates (0.0008% to 0.001% of transfused patients) [19] after the implementation of strategies to prevent the transfusion of blood components obtained from multiparous female donors. In our study, multiparous women will not be invited to donate plasma.
Treatment of a patient with TRALI involves immediate interruption of the transfusion.

*TRALI diagnostic criteria: [28]*
TRALI Type I: patients who do not have risk factors for ARDS and meet the following criteria:
A. i. Acute onset
ii. Hypoxemia ($PaO_2$/ $FiO_2$ ≤ 300 or $SpO_2$<90% in room air).
iii. Pulmonary edema with clear evidence of bilateral images ( CXR, CT or ultrasound)
iv. There is no evidence of left ventricular hypertrophy or, if present, it is not the main contributor to hypoxemia.
B. Initiation during or within 6 hours of transfusion.
C. Temporarily unrelated to an alternative risk factor for ARDS.

TRALI Type II: patients who have risk factors for ARDS (but have not been diagnosed with ARDS) or who have existing mild ARDS (Pa / Fi 200-300), but whose respiratory status is deteriorating and is considered to be due to a transfusion based on:
a. The results described in categories a and b of TRALI Type I, and
b. Stable respiratory status within 12 hours prior to transfusion

***Allergic - Anaphylactic Reactions***
Allergic reactions generally occur during or within 4 hours of transfusion and are most frequently associated with platelet transfusions (302 per 100,000 platelet units) [29]. Symptoms are caused by mediators such as histamine, released in mast cell and basophil activation [30]. Frequently, the clinical presentation is mild (rash, pruritus, urticaria, and angioedema are localized) and they are a solicited AE. Mild allergic transfusion reactions generally resolve with the administration of the usual treatment for allergic reactions, being able to restart the infusion of the unit. The transfusion should be stopped if symptoms reappear. The incidence of anaphylactic reactions is 8 / 100,000 units of transfused platelets. The most severe reactions generally present with bronchospasm, respiratory distress, and hypotension and constitute a SAE and should be reported as such immediately [31-35] .

***Acute hemolytic transfusion reactions***
Acute hemolytic transfusion reactions (ATRs) can occur when incompatible red blood cells are transfused or, less frequently, large amounts of plasma incompatible with the patient's ABO are transfused. The central pathophysiological mechanism is intravascular hemolysis and is considered a SAE. It presents with sudden fever and chills, retroperitoneal pain and dyspnea, hemoglobinuria and even disseminated intravascular coagulation, acute renal failure, and shock. Since fever and chills may be the only early signs, it is important to monitor the patient during the transfusion and stop the transfusion immediately if there is any change in vital signs or unexpected symptoms appear. Treatment is based on supportive measures to alleviate symptoms. Proper patient identification and adherence to all procedures related to handling pretransfusion specimens and administration of transfusions are essential to prevent ATRs [36].

***Transfusion Communicable Infections [39]***
The risk of transmission of infectious diseases through transfusions has been dramatically reduced, secondary to the great advances in unit testing, and pathogen inactivation for plasma units and their components [51] and the implementation of rigorous donor selection methods. In our environment, the samples are tested for HBV, HCV, HIV, HTLV and *Trypanosoma cruzi* [52]. In the United States, the risk of acquiring HBV and HIV by transfusion is 1:280,000 and 1:1,467,000, respectively [53 54]. Due to freezing of plasma samples, contamination by bacterial pathogens and CMV is extremely rare. There are only a dozen cases among the millions of transfusions reported in Germany and Canada of bacterial transmission. It is estimated that the potential source of this contamination would have been the water baths used to defrost the units. This is easily avoidable with proper cleaning and sterilization of defrosters [39].

#### 12. Ethical considerations

##### Statement of Good Clinical Practice

This study will be conducted in accordance with the protocol and with the following considerations:

- the ethical principles that have their origin in the Declaration of Helsinki,
- the current Good Clinical Practice (GCP) guidelines from the International Council of Harmonization (ICH),
- current laws and regulations.

The protocol, the Informed Consent forms (ICF) and other relevant documents have been reviewed and approved by INFANT Foundation, the participating authorities and sent to the Ethics Committee (EC) for evaluation.

##### Ethics Committee (EC):

A properly constituted EC, as outlined in the ICH guidelines for GCP, must review and approve:

- The protocol, ICF and any other materials that are provided to the participants before any subject can enter the study
- Any amendment or modification to the study protocol or to the CRF prior to implementation, unless the change is necessary to eliminate an immediate danger to patients, in which case the EC should be informed as soon as possible

In addition, they must be informed of any event that may affect the safety of patients or the continued conduct of the clinical study.

INFANT should receive a copy of the approval letter from the EC before sending drug supplies to the investigator at each hospital. The approval letter should include the title of the study, the documents reviewed, and the date of the review.

The investigator must keep on file the records of the EC review and approval of all study documents

##### Informed Consent Process

The ICF used by the researcher will be approved by the corresponding EC.

It is the responsibility of the investigator or delegated professional to obtain the written informed consent from each subject prior to their participation in the study and after the objectives, procedures, and potential risks of the study have been fully explained in language that the subject can understand. The ICF must be signed and dated by the researcher or delegated personnel who performed the consenting process.

The investigator or delegated staff will explain the nature of the study to the participant and answer all questions about the study. He/she will inform them that their participation is voluntary. The participant must sign the informed consent form (ICF) before any specific study activity takes place.

Participants who can understand but cannot write and / or read will have the IC read to them in the presence of an impartial witness, who will sign and date the ICF to confirm that informed consent was obtained.

Two copies will be signed. The investigator should keep one copy as part of the patient's study record, and the other signed copy should be given to the participant.

- Patients with cognitive vulnerability:
  They may be included after obtaining informed consent from their legal representative as appropriate:
  to. Patients who have a statement of disability: duty of who has been designated as curator whenever there is such a statement.
  b. Patients who can decide for themselves prior assistance of a "support" person according to provisions of the Law: usually the patient can designate who can help him/her decide.
  c. Patients without a judicial declaration of incapacity and who have not appointed a support or representative either: the patients' rights law will apply.
  The means to prove the representation can consist of any document that proves the appointment as curator (if applicable) or any advance directive of the patient where it establishes who represents him in health matters, or documents that prove the relationship according to Law 26529.

**13. Confidentiality and Protection of Personal Data**

The investigator will take all appropriate measures to ensure that the anonymity of each study subject is maintained in research records transmitted outside the health center. The original and essential documents of the study must be kept in strict confidence at each research center by the study team.

The personal data of the participants will be stored in the study center in a password-protected print and/or electronic format or in a locked room, to guarantee the exclusive access of authorized study personnel.

To protect the rights and freedoms of natural persons in relation to the processing of personal data, a unique and specific alphanumeric code will be assigned to the participants. The records or data sets of the participants that are transferred will contain the alphanumeric code; the names of the participants will not be transferred. All other identifiable data transferred between researchers will be identified by this unique, participant-specific code. The study center will maintain a confidential list of participants who participated in the study under strict security (password protected or in a locked room), linking the alphanumeric code of each participant with their real identity.

**14. Study Documentation**

Records and documents, including signed FCIs, regarding the conduct of this study will be retained by the investigator for 15 years after study completion, unless local regulations or institutional policies require a longer retention period.
The investigator from each participating institution should consult with INFANT before discarding or destroying any essential study documents after study completion or interruption. If approved, records must be destroyed in a way that ensures confidentiality.

#### 15. Study monitoring

##### 15.1. Monitoring of study sites

The study monitor will visit each site prior to enrolling the first patient and periodically during the study in accordance with the Approved Study Monitoring Plan.

The monitor receives weekly reports from each of the centers with the number of patients in a period of research and enrolled.

Monitoring visits and centralized monitoring activities will be carried out in accordance with all applicable regulatory requirements and standards in force in Argentina. It is understood that the monitor will contact each researcher and their responsible team regularly and will be allowed to verify the generation of the different study records.

It will be the monitor's responsibility to inspect the data capture systems at regular intervals throughout the study to verify compliance with the protocol and the integrity, precision and consistency of the data; and adherence to the ICH GCP and local regulations on conducting clinical research. The monitor should have access to laboratory reports, study medication, records associated with it, and other medical records necessary to verify the entries in the EDC.

An unblinded **monitor** will be assigned whose role will be limited to auditing the logistic data, management and study treatment registration according to the approved protocol and treatment instructions:

- reception and storage conditions
- randomization and administration
- return or destruction / final reconciliation

##### 15.2. Source document requirements

Investigators are required to prepare and maintain adequate and accurate patient records (source documents).

The researcher must keep all the source documents in the file of the data submitted to the study form (CRF). Some data generated from the study may be entered directly into the EDC system (direct data capture), after approval of the process by the Ethics Committees involved. Case report forms and source documents must be available at all times.

##### 15.3. Case Report Form Requirements

Study data obtained in the course of the study will be recorded in case report forms (CRF) and sent to INFANT in scan format. Once received, they will be archived and processed by entering the EDC system by trained INFANT personnel. All required CRFs must be completed for each patient enrolled in the study. The investigator should retain a copy of each patient's CRF case book as part of the study record and should be available at all times for inspection by authorized representatives of the sponsor and regulatory authorities.

#### 16 .. Audits and inspections

This study may be subject to a quality assurance audit or inspection. If this occurs, the investigator is responsible for:

- Provide access to all facilities, study data and documents necessary for inspection or audit.
- Communicate any information arising from the inspection of the regulatory authorities to INFANT immediately.
- Take all appropriate measures requested by INFANT to resolve the problems found during the audit or inspection.

Documents subject to audit or inspection include, but are not limited to, all source documents, CRF, medical records, correspondence, FCI, CE files, supporting laboratory quality control and certification documentation, and relevant records for the study being conducted. they maintain pharmacy facilities. The storage conditions of study material are also subject to inspection. Additionally, sponsor representatives may observe the conduct of any aspect of the clinical trial or its support activities both within and outside of the investigator's institution.

In all cases, the confidentiality of the data must be respected.

#### ANNEX

##### HEART FAILURE
##### NYHA SCALE (NEW YORK HEART ASSOCIATION)
##### FUNCTIONAL ASSESSMENT OF HEART FAILURE.

| NYHA FUNCTIONAL CLASSIFICATION | |
|---|---|
| Class I | Without limitation of physical activity. Normal physical exercise does not cause fatigue, palpitations, or dyspnea. |
| Class II | Slight limitation of physical activity. No symptoms at rest. Ordinary activity causes fatigue, palpitations, or dyspnea. |
| Class III | Marked limitation of physical activity. No symptoms at rest. Less than ordinary physical activity causes fatigue, palpitations or dyspnea. |
| Class IV | Inability to carry out any physical activity; symptoms of heart failure are present even at rest and increase with any physical activity. |

##### CIRRHOSIS (PORTAL BAVENO IV HYPERTENSION CONSENSUS)

| CIRRHOSIS STAGES | |
|---|---|
| Stage 1 | Absence of esophageal varices and ascites |
| Stage 2 | Esophageal varices without a history of bleeding and without ascites. |
| Stage 3 | Presence of ascites with or without esophageal varices. |
| Stage 4 | Gastrointestinal bleeding due to portal hypertension, with or without ascites. |

**KIDNEY DISEASE CLASSIFCIATION KDIGO.**

| DEGREE | GLOMERULAR FILTRATION ML / MIN / 1.73 M2 | DESCRIPTION |
|---|---|---|
| GRADE 1 | > 90 G5 | Normal or elevated |
| GRADE 2 | 60-89 | Slightly diminished |
| GRADE 3 | 45-59 | Slight to moderately decreased |
| GRADE 3b | 30-44 | Moderate to severely diminished |
| GRADE 4 | 15-29 | Severely diminished |
| GRADE 5 | <15 | Kidney failure |

#### References


1. Fei Zhou *, Ting Yu *, Ronghui Du *, Guohui Fan *, Ying Liu *, Zhibo Liu *, Jie Xiang *, Yeming Wang, Bin Song, Xiaoying Gu, Lulu Guan, Yuan Wei, Hui Li,

Xudong Wu, Jiuyang Xu, Shengjin Tu, Yi Zhang, Hua Chen, Bin Cao. Clinical course and risk factors for mortality of adult inpatients with COVID-19 in Wuhan, China: a retrospective cohort study. Lancet 2020; 395: 1054–62 doi: 10.1016 / S0140-6736 (20) 30566-3

2. Chen N., Zhou M., Dong X., Qu J., Gong F., Han Y. Epidemiological and clinical characteristics of 99 cases of 2019 novel coronavirus pneumonia in Wuhan, China: a descriptive study. Lancet. 2020; 395: 507–513. 10.1016 / S0140-6736 (20) 30211-7

3. Garg S, Kim L, Whitaker M, et al. Hospitalization Rates and Characteristics of Patients Hospitalized with Laboratory-Confirmed Coronavirus Disease 2019 - COVID-NET, 14 States, March 1–30, 2020. MMWR Morb Mortal Wkly Rep 2020; 69: 458–464. DOI: http://dx.doi.org/10.15585/mmwr.mm6915e3

4. Ministry of Health of Argentina. www.argentina.gob.ar/salud/coronavirus-COVID-19

5. Willan, J., King, AJ, Jeffery, K., & Bienz, N. (2020). Challenges for NHS hospitals during covid-19 epidemic. BMJ, m1117. doi: 10.1136 / bmj.m1117

6. Goodarz Kolifarhood , [1,3] Mohammad Aghaali , [1] Hossein Mozafar Saadati , [1] Niloufar Taherpour , [1] Sajjad Rahimi , [1,2] Neda Izadi , [3] and Seyed Saeed Hashemi Nazari[4,*]. Epidemiological and Clinical Aspects of COVID-19; a Narrative Review. Arch Acad Emerg Med . 2020; 8 (1): e41. PMCID: PMC7117787. PMID: 32259130

7. Center for Disease Control and Prevention Interim Guidance for Healthcare Facilities: Preparing for Community Transmission of COVID-19 in the United States

8. Orenstein, WA, Heymann, DL, Ellis, RJ, Rosenberg, RL, Nakano, J., Halsey, NA, Witte, JJ (1981). Prophylaxis of varicella in high-risk children: Dose-response effect of zoster immune globulin. The Journal of Pediatrics, 98 (3), 368–373.doi: 10.1016 / s0022-3476 (81) 80697-x

9. Centers for Disease Control and Prevention. Measles (Rubella) For Healthcare Professionals.

20. Yun Feng ; Yun Ling , Tao Bai , Yusang Xie ; Jie Huang , Jian Li , Weining Xiong , Dexiang Yang , Rong Chen ; Fangying Lu ; Yunfei Lu , et al. COVID-19 with Different Severity: A Multi-center Study of Clinical Feature. doi.org/10.1164/rccm.202002-0445OC

21. Zhou F , Yu T , Du R , Fan G , Liu Y , Liu Z , Xiang J , Wang Y , Song B , Gu X , Guan L , Wei Y , Li H , Wu , Xu J , Tu S , Zhang Y , Chen H , Cao B . Clinical course and risk factors for mortality of adult inpatients with COVID-19 in Wuhan, China: a retrospective cohort study. Lancet. 2020 Mar 28; 395 (10229): 1054-1062. doi: 10.1016 / S0140-6736 (20) 30566-3

22. Afaf Alblooshi , Alia Alkalbani , Ghaya Albadi , Hassib Narchi , and Graham Hall . Is forced oscillation technique the next respiratory function test of choice in childhood asthma. World J Methodol . 2017 Dec 26; 7 (4): 129–138. doi: 10.5662 / wjm.v7.i4.129

23. Garg S, Kim L, Whitaker M, et al. Hospitalization Rates and Characteristics of Patients Hospitalized with Laboratory-Confirmed Coronavirus Disease 2019 - COVID-NET, 14 States, March 1–30, 2020. MMWR Morb Mortal Wkly Rep 2020; 69: 458–464. DOI: http://dx.doi.org/10.15585/mmwr.mm6915e3

24. Feng, Y. et al. COVID-19 with Different Severity: A Multi-center Study of Clinical Features. Am. J. Respir. Crit. Care Med. **0** , null. https://doi.org/10.1164/rccm.202002-0445OC

25. Grasselli G, Zangrillo A, Zanella A, et al. Baseline Characteristics and Outcomes of 1591 Patients Infected With SARS-CoV-2 Admitted to ICUs of the Lombardy Region, Italy. *JAMA.* Published online April 06, 2020. doi: 10.1001 / jama.2020.5394

26. Integrated Surveillance Bulletin. National Directorate of Epidemiology and Analysis of the Health Situation. N ° 424 - EW 39 - September 2018

27. Report of the WHO-China Joint Mission on Coronavirus Disease 2019 (COVID-19), 16-24 February 2020. Available from: https://www.who.int/docs/default-source/coronaviruse/who-china-joint -mission-on-covid-19-final-report.pdf . Accessed March 09, 2020.

28. Updates on COVID-19 in Republic of Korea. April 15, 2020, Division of Risk assessment and International cooperation, Korean Center for Disease Control and Prevention https://www.cdc.go.kr/board/board.es?mid=a30402000000&bid=0030&act=view&list_no=366892&tag=&nPage= 1

29. Wu, JT, Leung, K., Bushman, M., Kishore, N., Niehus, R., de Salazar, PM,… Leung, GM (2020). *Estimating clinical severity of COVID-19 from the transmission dynamics in Wuhan, China. Nature Medicine.* doi: 10.1038 / s41591-020-0822-7

30. Coronavirus disease 2019 (COVID-19) in the EU / EEA and the UK - eighth update, European Center for Disease Prevention and

Control. https://www.ecdc.europa.eu/en/publications-data/rapid-risk-assessment-coronavirus-disease-2019-covid-19-pandemic-eighth-update

31. Immediate and Near Future Prediction of COVID-19 Patients in the US Population Aged 65+ With the Prior Medical Conditions of Hypertension, Cardiovascular and Lung Diseases: Methods, Models and Acute Care Estimates. Arni SR Srinivasa Rao, Douglas D Miller, Adam E Berman, David C Hess, Steven G Krantz medRxiv 2020.04.12.20062166; doi: https://doi.org/10.1101/2020.04.12.20062166

32. Chen T , Dai Z , Mo P , Li X , Ma Z , Song S , Chen X , Luo M , Liang K , Gao S , Zhang Y , Deng L , Xiong Y . Clinical characteristics and outcomes of older patients with coronavirus disease 2019 (COVID-19) in Wuhan, China (2019): a single-centered, retrospective study. The Journals of Gerontology: Series A, glaa089, https://doi.org/10.1093/gerona/glaa089

33. Q. Li, X. Guan, P. Wu, et al. Early Transmission Dynamics in Wuhan, China, of Novel Coronavirus – Infected Pneumonia. N Engl J Med (January 2020), doi: 10.1056 / NEJMoa2001316NEJMoa2001316

34. Y. Liu, AA Gayle, A. Wilder-Smith, J. Rocklöv, The reproductive number of COVID-19 is higher compared to SARS coronavirus. J Travel Med (February 2020), 10.1093 / jtm / taaa021

35. K. Duan, et al. The feasibility of convalescent plasma therapy in severe COVID-19 patients: a pilot study. medRxiv 2020.03.16.20036145; doi: https://doi.org/10.1101/2020.03.16.20036145

36. Cheryl D. Fryar, MSPH, Yechiam Ostchega, Ph.D., RN, Craig M. Hales, MD, MPH, Guangyu Zhang, Ph.D., and Deanna Kruszon-Moran, MS Hypertension Prevalence and Control Among Adults: United States , 2015–2016. US DEPARTMENT OF HEALTH AND HUMAN SERVICES. Center for Disease Control and Prevention.

37. US Department of Health and Human Services, Center for Disease Control and Prevention. National Diabetes Statistics Report, Estimates of Diabetes and Its Burden in the United States

45. John Hodgson.The pandemic pipeline. Nature Biotechnology. doi: 10.1038 / d41587-020-00005-z

46. W. Guan, Z. Ni, Yu Hu, W. Liang, C. Ou, J. He, L. Liu, H. Shan, C. Lei, DSC Hui, B. Du, L. Li, G. Zeng, K.-Y. Yuen, R. Chen, C. Tang, T. Wang, P. Chen, J. Xiang, S. Li, Jin-lin Wang, Z. Liang, Y. Peng, L. Wei, Y. Liu, Ya-hua Hu, P. Peng, Jian-ming Wang, J. Liu, Z. Chen, G. Li, Z. Zheng, S. Qiu, J. Luo, C. Ye, S. Zhu, and N. Zhong, for the China Medical Treatment Expert Group for Covid-19. Clinical Characteristics of Coronavirus Disease 2019 in China. The New England Journal of Medicine. DOI: 10.1056 / NEJMoa2002032

47. Temet M. McMichael, Ph.D., Dustin W. Currie, Ph.D., Shauna Clark, RN, Sargis Pogosjans, MPH, Meagan Kay, DVM, Noah G. Schwartz, MD, James Lewis, MD, Atar Baer, Ph.D., Vance Kawakami, DVM, Margaret D. Lukoff, MD, Jessica Ferro, MPH, Claire Brostrom-Smith, MSN, Thomas D. Rea, MD, Michael R. Sayre, MD, Francis X. Riedo, MD, Denny Russell, BS, Brian Hiatt, BS, Patricia Montgomery, MPH, Agam K. Rao, MD, Eric J. Chow, MD, Farrell Tobolowsky, DO, Michael J. Hughes, MPH, Ana C. Bardossy, MD, Lisa P Oakley, Ph.D., Jesica R. Jacobs, Ph.D., Nimalie D. Stone, MD, Sujan C. Reddy, MD, John A. Jernigan, MD, Margaret A. Honein, Ph.D., Thomas A. Clark, MD, and Jeffrey S. Duchin, MD Epidemiology of Covid-19 in a Long-Term Care Facility in King

County, Washington. The New England Journal of Medicine. DOI: 10.1056 / NEJMoa2005412

48. DF Gudbjartsson, A. Helgason, H. Jonsson, OT Magnusson, P. Melsted, GL Norddahl, J. Saemundsdottir, A. Sigurdsson, P. Sulem, AB Agustsdottir, B. Eiriksdottir, R. Fridriksdottir, EE Gardarsdottir, G. Georgsson , OS Gretarsdottir, KR Gudmundsson, TR Gunnarsdottir, A. Gylfason, H. Holm, BO Jensson, A. Jonasdottir, F. Jonsson, KS Josefsdottir, T. Kristjansson, DN Magnusdottir, L. le Roux, G. Sigmundsdottir, G. Sveinbjornsson, KE Sveinsdottir, M. Sveinsdottir, EA Thorarensen, B. Thorbjornsson, A. Löve, G. Masson, I. Jonsdottir, AD Möller, T. Gudnason, KG Kristinsson, U. Thorsteinsdottir, and K. Stefansson. Spread of SARS-CoV-2 in the Icelandic Population. New England Journal of Medicine. DOI: 10.1056 / NEJMoa2006100

49. Michelle L. Holshue, MPH, Chas DeBolt, MPH, Scott Lindquist, MD, Kathy H. Lofy, MD, John Wiesman, Dr.PH, Hollianne Bruce, MPH, Christopher Spitters, MD, Keith Ericson, PA-C., Sara Wilkerson, MN, Ahmet Tural, MD, George Diaz, MD, Amanda Cohn, MD, LeAnne Fox, MD, Anita Patel, Pharm.D., Susan I. Gerber, MD, Lindsay Kim, MD, Suxiang Tong, Ph.D ., Xiaoyan Lu, MS, Steve Lindstrom, Ph.D., Mark A. Pallansch, Ph.D., William C. Weldon, Ph.D., Holly M. Biggs, MD, Timothy M. Uyeki, MD, and Satish K. Pillai, MD First Case of 2019 Novel Coronavirus in the United States. New England Journal of Medicine. DOI: 10.1056 / NEJMoa2001191

50. Pavan K. Bhatraju, MD, Bijan J. Ghassemieh, MD, Michelle Nichols, MD, Richard Kim, MD, Keith R. Jerome, MD, Arun K. Nalla, Ph.D., Alexander L. Greninger, MD, Sudhakar Pipavath , MD, Mark M. Wurfel, MD, Ph.D., Laura Evans, MD, Patricia A. Kritek, MD, T. Eoin West, MD, MPH, Andrew Luks, MD, Anthony Gerbino, MD, Chris R. Dale , MD, Jason D. Goldman, MD, Shane O'Mahony, MD, and Carmen Mikacenic, MD Covid-19 in Critically Ill Patients in the Seattle Region - Case Series New England Journal of Medicine. DOI: 10.1056 / NEJMoa2004500

51. W. Guan, Z. Ni, Yu Hu, W. Liang, C. Ou, J. He, L. Liu, H. Shan, C. Lei, DSC Hui, B. Du, L. Li, G. Zeng, K.-Y. Yuen, R. Chen, C. Tang, T. Wang, P. Chen, J. Xiang, S. Li, Jin-lin Wang, Z. Liang, Y. Peng, L. Wei, Y. Liu, Ya-hua Hu, P. Peng, Jian-ming Wang, J. Liu, Z. Chen, G. Li, Z. Zheng, S. Qiu, J. Luo, C. Ye, S. Zhu, and N. Zhong. Clinical Characteristics of Coronavirus Disease 2019 in China. New England Journal of Medicine. DOI: 10.1056 / NEJMoa2002032

52. Arciuolo, RJ, Jablonski, RR, Zucker, JR, & Rosen, JB (2017). *Effectiveness of Measles Vaccination and Immune Globulin Post-Exposure Prophylaxis in an Outbreak Setting — New York City, 2013. Clinical Infectious Diseases, 65 (11), 1843–1847.* doi: 10.1093 / cid / cix639

53. Swamy, GK, & Dotters-Katz, SK (2019). *Safety and varicella outcomes after varicella zoster immune globulin administration in pregnancy. American Journal of Obstetrics and Gynecology.* doi: 10.1016 / j.ajog.2019.07.003

54. Levin MJ, Duchon JM, Swamy GK, Gershon AA. Varicella zoster immune globulin (VARIZIG) administration up to 10 days after varicella exposure in pregnant women, immunocompromised participants, and infants: varicella outcomes and safety results from a large, open-label, expanded-access program. PLoS One 2019; 14: e0217749

55. MMWR. Recommendation of the Immunization Practices Advisory Committee (ACIP) Postexposure Prophylaxis of Hepatitis B. https://www.cdc.gov/mmwr/preview/mmwrhtml/00022736.htm

67. Transfusion-associated circulatory overload (TACO) Definition (2018) IHN / ISBT haemovigilance working party / AABB

77. CDC. NHSN Biovigilance Component, Hemovigilance Module Surveillance Protocol v2.1.3. Atlanta: Centers for Disease Control and Prevention, 2014.

78. Simons FE, Ardusso LR, Bilo MB, et al, and the World Allergy Organization. World Allergy Organization anaphylaxis guidelines: summary. J Allergy Clin Immunol 2011; 127: 587, e1–22

79. Tinegate H, Birchall J, Gray A, et al, and the BCSH Blood Transfusion Task Force. Guideline on the investigation and management of acute transfusion reactions. Prepared by the BCSH Blood Transfusion Task Force. Br J Haematol 2012; 159: 143–53.

80. 15 Lin RY, Curry A, Pesola GR, et al. Improved outcomes in patients with acute allergic syndromes who are treated with combined H1 and H2 antagonists. Ann Emerg Med 2000; 36: 462-68

81. 16 Runge JW, Martinez JC, Caravati EM, Williamson SG, Hartsell SC. Histamine antagonists in the treatment of acute allergic reactions. Ann Emerg Med 1992; 21: 237–427

82. Fung M, Grossman BJ, Hillyer CD, Westhoff CM. Technical Manual, 18th edn. Glen Burnie, MD: AABB Press, 2014.

96. Administrative and Technical Regulations, RM 797/13 - 139/14 - 1507/15 Directorate of Blood and Blood Products Ministry of Health of the Nation. http://www.msal.gob.ar/disahe/images/stories/pdf/normas-hemoterapia.pdf

97. Zou S, Dorsey KA, Notari EP, Foster GA, Krysztof DE, Musavi F, Dodd RY, Stramer SL. Prevalence, incidence, and residual risk of human immunodeficiency virus and hepatitis C virus infections among United States blood donors since the introduction of nucleic acid testing. Transfusion. 2010; 50: 1495–504. [PubMed: 20345570]

98. Zou S, Stramer SL, Notari EP, Kuhns MC, Krysztof D, Musavi F, Fang CT, Dodd RY. Current incidence and residual risk of hepatitis B infection among blood donors in the United States. Transfusion. 2009; 49: 1609-20. [PubMed: 19413732]
