## Supplementary material for "Prevention of severe COVID-19 in the elderly by early high-titer plasma": Analysis Plan

IECS
INSTITUTO DE EFECTIVIDAD
CLINICA Y SANITARIA

### EFFICACY EVALUATION OF COVID-19 CONVALESCENT PLASMA ADMINISTRATION IN THE DECREASE OF PROGRESSION TO SEVERE DISEASE IN ELDERLY ADULTS WITH MILD SYMPTOMS OF SARS-COV2

Final Analysis Plan

November 6th 2020

Report to:
Main Researcher: Fernando Polack

Written by:
Biostatistics: L. Gibbons

**Table of contents**


EFFECTIVENESS ASSESSMENT CRITERIA 4

Primary criteria of effectiveness evaluation 4

Secondary criteria of effectiveness evaluation 5

STATISTICAL ANALYSIS 5

SAMPLE SIZE AND DETENTION RULES ACCORDING THE PROTOCOL. 6

VARIABLES DEFINITION 7

Table 1: Primary outcomes definition. 7

Table 2: Secondary outcomes definition. 8

Table 2: Secondary outcomes definition (cont.) 10

Table 3: Adverse events definition 11

RESULT TABLES 12

Figure 1: Consort 12

Table 4: Characteristics of ITT population 13

Table 5: Characteristics of ITTm population 14

Table 6: Primary Endpoint. Proportion of cases - ITT 15

Table 7: Primary Endpoint: Time to event - ITT 15

Figure 2: Kaplan-Meier survival curve of severe respiratory disease due to SARS-CoV2 by treatment group – ITT Analysis 15

Table 8: Primary outcome: Proportion of primary endpoint cases by age group - ITT 15

Table 9: Primary outcome. Time to event by group age - ITT 15

Figure 3: Kaplan-Meier survival curve of severe respiratory disease due SARS-CoV2 in 65-74 years old by treatment group – ITT Analysis 16

Figure 4: Kaplan-Meier survival curve of severe respiratory disease due SARS-CoV2 in ≥75 years old by treatment group – ITT Analysis 16

Table 10: Secondary outcomes: Case proportions –ITT 16

Table 11: Secondary outcomes: Time to event – ITT analysis 17

Figure 5: Kaplan- Meier survival curve. Time to support with maximum oxygen therapy and/ or non-invasive respiratory support and/or admission to UCI and/or invasive mechanical ventilation assistance requirement by treatment group- ITT analysis 17

Figure 6: Kaplan- Meier survival curve. Time to critical systemic Illness by treatment group- ITT analysis 17

Figure 7: Kaplan- Meier survival curve. Time to mortality due to COVID-19 by treatment group- ITT analysis 17

Figure 8: Kaplan- Meier survival curve. Time to Support with maximum oxygen therapy and/ or non-invasive respiratory support and/or admission to UCI and/or invasive mechanical ventilation assistance requirement and/or critical systemic Illness and/or mortality – ITT analysis 17

Table 12: Requested adverse events – ITT analysis 18

Table 13: Primary Endpoint. Proportion of cases - ITTm 19

Table 14: Primary Endpoint: Time to event - ITTm 19
Figure 9: Kaplan-Meier survival curve of severe respiratory disease due to SARS-CoV2 by treatment group – ITTm Analysis 19
Table 15: Primary outcome: Proportion of primary endpoint cases by age group - ITTm 19
Table 16: Primary outcome. Time to event by age group - ITTm 19
Figure 10: Kaplan-Meier survival curve of severe respiratory disease due SARS-CoV2 in 65-74 years old by treatment group – ITTm analysis 19
Figure 11: Kaplan-Meier survival curve of severe respiratory disease due SARS-CoV2 in ≥75 years old by treatment group – ITTm analysis 19
Table 17: Secondary Outcomes: Case proportions – ITTm analysis 20
Table 18: Secondary outcomes: Time to event – ITTm analysis 21
Figure 12: Kaplan- Meier survival curve. Time to support with maximum oxygen therapy and/ or non-invasive respiratory support and/or admission to UCI and/or invasive mechanical ventilation assistance requirement by treatment group – ITTm analysis 21
Figure 13: Kaplan- Meier survival curve. Time to critical systemic Illness by treatment group – ITTm Analysis 21
Figura 14: Kaplan- Meier survival curve. Time to mortality due to COVID-19 by treatment group- ITT analysis – ITTm Analysis 22
Figura 15: Kaplan- Meier survival curve. Time to Support with maximum oxygen therapy and/ or non-invasive respiratory support and/or admission to UCI and/or invasive mechanical ventilation assistance requirement and/or critical systemic Illness and/or mortality – ITTm analysis 22
Table 19: Requested adverse events – ITTm analysis 22

#### OBJETIVES

To evaluate the efficacy of convalescent plasma, from 12 hours of administration, in reducing progression to severe respiratory disease in subjects between 65-74 years of age with at least one comorbidity or ≥75 years old, with mild symptomatology and with early COVID-19 diagnosis up to 15 days of treatment administration (16-18 days of disease considering the symptomatic period previous enrollment).

Secondary outcomes include determining if convalescent plasma administration up to 15 days, or from there up to a maximum of 25 days since treatment administration if the patient remains in hospital, achieve to:

- To reduce the need of respiratory support with maximum oxygen therapy (oxygen reservoir mask) and/or the use of non-invasive respiratory support (NIV support, including CPAP) and/or admission to the intensive care unit (ICU) and/or invasive mechanical ventilation assistance because of SARS-CoV2 in patients between 65-74 years of age with at least one comorbidity or in patients ≥75 years old with COVID-19.
- To reduce critical systemic illness defined as (a) presence of acute respiratory failure (PaO2/FiO2 ≤ 200 mm Hg.) and/ or (b) shock (defined by the need of support with vasoactive drugs to maintain the mean arterial pressure ≥ 65 mm Hg.) and/or (c) multiorganic shock syndrome: acute renal injury (defined by creatinine increase two or more times from baseline or creatinine increase of 0.3 mg/dl), increase of hepatic enzymes (transaminase) in three or more times the normal range, acute cardiomyopathy (define by the increase of troponin above normal range and/ or new electrocardiographic or echocardiographic abnormalities due to myocardial damage) due to SARS-Cov2 in patients between 65-74 years of age with at least one comorbidity or in patients ≥75 years old with COVID-19.
- To reduce the mortality due to SARS-Cov2 in patients between 65-74 years of age with at least one comorbidity or in all patients ≥75 years old with COVID-19.
- To describe the safety of convalescent plasma administration in patients between 65-74 years of age with at least one comorbidity or in all patients ≥75 years old with COVID-19.
- To explore the concentration of anti-SARS-CoV2 in the plasma of the participants associated with the absence of severe respiratory disease obtained 24 hours after infusion.

#### EFFECTIVENESS ASSESSMENT CRITERIA

##### Primary criteria of effectiveness evaluation

Severe respiratory disease due to SARS-CoV2, confirmed by viral ARN detection with RT-PCR, and defined by the presence of any of the following two variables in a non-exclusive way: (a) respiratory rate (RR) ≥30 per minute (b) O2 sat<93% at room air. The primary endpoint will be determined from 12 hours from the start of the infusion until 15 days since the administration of the treatment (12h – 15 d).

##### Secondary criteria of effectiveness evaluation

*(Up to 15 days or from there up to a maximum of 25 days since treatment administration if the patient remains in hospital).*

- Need of respiratory support with maximum oxygen therapy (oxygen reservoir mask) and/or the use of non-invasive respiratory support (NIV support including CPAP) and/or admission to the intensive care unit (ICU) and/or invasive mechanical ventilation assistance due to SARS-CoV2.
- Critical systemic illness defined as: (a) presence of acute respiratory failure (PaO2/FiO2 ≤ 200 mm Hg.) and/ or (b) shock and/or (c) multiorganic shock syndrome, acute renal injury, increase of hepatic enzymes and acute cardiomyopathy due of SARS-Cov2.
- Mortality due to SARS-Cov2.
- Combination of secondary criteria of effectiveness evaluation, #1 (Need of respiratory support…) and/ or #2 (Severe disease define as...) and/or #3 (Mortality due to SARS-Cov2).
- Duration of oxygen support requirement in patients with COVID-19 due to O2 sat<93% at room air.
- Comparison of IgG antibodies titer.

#### STATISTICAL ANALYSIS

The socio-demographic characteristics, vital signs and comorbidities of primary and secondary interest of the participants will be described. For both primary and secondary outcomes the proportion by treatment group and the relative risk with its 95% confidence interval will be reported. A logistic regression model will be used in case it is necessary to adjust for covariates. Time to event will be evaluated using Kaplan- Meier survival analysis comparing the groups with Log-Rank test. When necessary Cox regression model will be used to make adjustments. The analysis of the primary outcome will be done stratifying by age groups.

The analysis will be made over two populations:

1) Intention-to-treat (ITT). All the randomized patients with follow-up until 15 days from transfusion beginning date (or since randomization if they had not received treatment) will be included.
2) Modified intention-to-treat (mITT). All the randomized patients with follow-up until 15 days from transfusion beginning date will be included, with the exception of patients with the following conditions:
    a. Has not received the treatment
    b. Has lost eligibility criteria between randomization and treatment administration, regardless if they had received the treatment. (I.e., those patients who, in advance or at the time of starting treatment, had already developed severe disease, primary outcome of the study).

In addition, serious and non-serious adverse events according to treatment group will be reported

Main conclusions will be based on the mITT population analysis.

#### SAMPLE SIZE AND DETENTION RULES ACCORDING THE PROTOCOL.

- Given the relative complexity of implementing this intervention, the minimally clinically important difference was set at a 40% relative reduction for an expected outcome rate of 50% in the control group reduced to 30% in the intervention group.
- A total sample size of 210 subjects (105 per trial arm) was estimated to have 80% power at a significance level (alpha) of 0.05 using a two-sided z-test with continuity correction.

#### VARIABLES DEFINITION

Table 1: Primary outcomes definition.

| N | Variable names | Definition | Variables in the CRF |
|---|---|---|---|
| (1) | Severe respiratory disease due to SARS-CoV2 | Presence of respiratory rate ≥30 per minute or O2 sat<93% at room air from 12 hours from the start of the infusion until 15 days since the administration of the treatment (12h – 15 d).<br>- Yes (At least one day had some of the two events)<br>- No (None of the two events in the 15 days) | Variables definition in (2) and (3) |
| (2) | Respiratory rate (RR) ≥30 per minute | Presence of respiratory rate ≥30 per minute from 12 hours from the start of the infusion until 15 days since the administration of the treatment (12h – 15 d).<br>- Yes (At least one day had some of the two events)<br>- No (None of the two events in the 15 days) | Respiratory rate ≥30 per minute (**Form 7, variable 4**).<br><br>12 hours from the start of the infusion until 15 days since the administration of the treatment (12h – 15 d). (**Form 5, Variable 2 y 5**) |
| (3) | Oxygen saturation <93% at room air | Presence O2 sat<93% at room air from 12 hours from the start of the infusion until 15 days since the administration of the treatment (12h – 15 d).<br>- Yes (At least one day had some of the two events)<br>- No (None of the two events in the 15 days) | Oxygen saturation <93% at room air between (**Form 7, variable 5**).<br><br>12 hours from the start of the infusion until 15 days since the administration of the treatment (12h – 15 d). (**Form 5, Variable 2 y 5**) |

Table 2: Secondary outcomes definition.

| N | Variable names | Definition | Variables in the CRF |
|---|---|---|---|
| (4) | Oxygen support and/or non-invasive respiratory support and/or admission to ICU and/or invasive mechanical ventilation assistance. | Oxygen support and/or non-invasive respiratory support and/or admission to ICU and/or invasive mechanical ventilation assistance from 12 hours from the start of the infusion until 15/25 days if the patient remains in hospital.<br>- Yes (At least one of the four events was registered in the 15/25 days)<br>- No (None of the four events in the 15/25 days) | Variables definition in (5), (6), (7) y (8) |
| (5) | Respiratory support with maximum oxygen therapy (oxygen reservoir mask) | Need of oxygen 100% support from 12 hours from the start of the infusion until 15/25 days if the patient remains in hospital.<br>- Yes (the event was registered in the 15/25 days)<br>- No (The event was not registered in the 15/25 days) | Oxygen support (**Form 7, Variable 14**) + (**Form 7, variable 14b**)<br><br>12 hours from the start of the infusion until 15/25 days if the patient remains in hospital. (**Form 5, Variable 2 y 5**) |
| (6) | Non-invasive respiratory support | Need of non-invasive respiratory support from 12 hours from the start of the infusion until 15/25 days if the patient remains in hospital.<br>- Yes (the event was registered in the 15/25 days)<br>- No (The event was not registered in the 15/25 days) | Non-invasive respiratory support (**Form 7, variable 16**)<br><br>12 hours from the start of the infusion until 15/25 days if the patient remains in hospital. (**Form 5, Variable 2 y 5**) |
| (7) | UCI admission | Admission to the intensive care unit (ICU) from 12 hours from the start of the infusion until 15/25 days if the patient remains in hospital.<br>- Yes (the event was registered in the 15/25 days)<br>- No (The event was not registered in the 15/25 days) | UCI admission (**Form 7, variable 15**)<br><br>12 hours from the start of the infusion until 15/25 days if the patient remains in hospital. (**Form 5, Variable 2 y 5**) |
| (8) | Invasive mechanical ventilation assistance requirement | Invasive mechanical ventilation assistance requirement from 12 hours from the start of the infusion until 15/25 days if the patient remains in hospital.<br>- Yes (the event was registered in the 15/25 days)<br>- No (The event was not registered in the 15/25 days) | Invasive mechanical ventilation assistance requirement (**Form 7, variable 17**)<br><br>12 hours from the start of the infusion until 15/25 days if the patient remains in hospital. (**Form 5, Variable 2 y 5**). |
| (9) | Critical systemic illness | Presence of acute respiratory failure, shock or multiorganic shock syndrome | Variables definition (10), (11) y (12) |

| | | | |
|---|---|---|---|
| | | from 12 hours from the start of the infusion until 15/25 days if the patient remains in hospital.<br>- Yes (At least one of the three events was registered in the 15/25 days)<br>- No (None of the three events in the 15/25 days) | |
| (10) | Acute respiratory failure | Presence of acute respiratory failure: PaO2/FiO2 ≤ 200 mmHg from 12 hours from the start of the infusion until 15/25 days if the patient remains in hospital.<br>- Yes (the event was registered in the 15/25 days)<br>- No (The event was not registered in the 15/25 days) | Acute respiratory failure (**Form 7, variable 18**)<br><br>12 hours from the start of the infusion until 15/25 days if the patient remains in hospital. (**Form 5, Variable 2 y 5**). |
| (11) | Shock | Need of support with vasoactive drugs to maintain the mean arterial pressure ≥ 65 mm Hg from 12 hours from the start of the infusion until 15/25 days if the patient remains in hospital.<br>- Yes (the event was registered in the 15/25 days)<br>- No (The event was not registered in the 15/25 days) | Shock (**Form 7, variable 20**).<br><br>12 hours from the start of the infusion until 15/25 days if the patient remains in hospital. (**Form 5, Variable 2 y 5**). |

Table 2: Secondary outcomes definition (cont.)

| N | Variable names | Definition | Variables in the CRF |
|---|---|---|---|
| (12) | Multiorganic shock syndrome | Presence of some of the following events from 12 hours from the start of the infusion until 15/25 days if the patient remains in hospital.<br>- Acute renal injury: increase in two or more times from baseline or creatinine increase of 0.3 mg/dl.<br>- Increase of hepatic enzymes (transaminase) in three or more times the normal range.<br>- Acute cardiomyopathy: increase of troponin above normal range and/ or new electrocardiographic or echocardiographic abnormalities due to myocardial damage.<br><br>- Yes (At least one of the three events was registered in the 15/25 days)<br>- No (None of the three events in the 15/25 days) | Acute renal injury (**Form 7, variable 19.a**)<br><br>Increase of hepatic enzymes (**Form 7, variable 19.b**)<br><br>Acute cardiomyopathy (**Form 7, variable 19.c**)<br><br>12 hours from the start of the infusion until 15/25 days if the patient remains in hospital (**Form 5, Variable 2 y 5**). |
| (13) | Mortality due to covid-19 | Mortality due to covid-19 rom 12 hours from the start of the infusion until 15/25 days if the patient remains in hospital.<br>- Yes (the event was registered in the 15/25 days)<br>- No (The event was not registered in the 15/25 days) | Patient death (**Form 7, variable 21**).<br><br>12 hours from the start of the infusion until 15/25 days if the patient remains in hospital (**Form 5, Variable 2 y 5**). |
| (14) | Combination of secondary criteria of effectiveness evaluation- (4) or (9) or (13) | Need of respiratory support with oxygen and/or the use of non-invasive respiratory support and/or admission to the ICU and/or critical systemic illness or mortality due to COVID-19 from 12 hours from the start of the infusion until 15/25 days if the patient remains in hospital.<br>- Yes (At least one of the six events was registered in the 15/25 days)<br>- No (None of the six events in the 15/25 days) | Variables definition in (4), (9) y (13) |
| (15) | Duration of oxygen support requirement in patients with COVID-19 due to O2 sat<93% at room air. | Number of days that oxygen support was required | Oxygen support (**Form 7, variable 14**) |

Table 3: Adverse events definition

| N | Variable names | Definition | Variables in the CRF |
|---|---|---|---|
| (16) | Cardiac volume overload associated with the transfusion | Presence of cardiac volume overload associated with the transfusion during the infusion and between 12 hours after.<br>- Yes (the event was registered)<br>- No (The event was not registered) | **Form 6, variable 3** |
| (17) | Allergic or anaphylactic reaction | Presence of allergic or anaphylactic reaction during the infusion and between 12 hours after.<br>- Yes (the event was registered)<br>- No (The event was not registered) | **Form 6, variable 4** |
| (18) | Thrombophlebitis | Presence of thrombophlebitis during the infusion and between 12 hours after.<br>- Yes (the event was registered)<br>- No (The event was not registered) | **Form 6, variable 5** |
| (19) | Vasovagal syndrome | Presence of vasovagal syndrome during the infusion and between 12 hours after.<br>- Yes (the event was registered)<br>- No (The event was not registered) | **Form 6, variable 6** |
| (20) | Hematoma at site | Presence of hematoma at site during the infusion and between 12 hours after.<br>- Yes (the event was registered)<br>- No (The event was not registered) | **Form 6, variable 7** |
| (21) | Nervous injury | Presence of nervous injury at site during the infusion and between 12 hours after.<br>- Yes (the event was registered)<br>- No (The event was not registered) | **Form 6, variable 8** |
| (22) | Tetany (hyperventilation) | Presence of tetany (hyperventilation) during the infusion and between 12 hours after.<br>- Yes (the event was registered)<br>- No (The event was not registered) | **Form 6, variable 9** |
| (23) | Total events | Total number of events | **Sum of (16), (17), (18), (19), (20), (21), (22)** |

#### RESULT TABLES

Figure 1: Consort

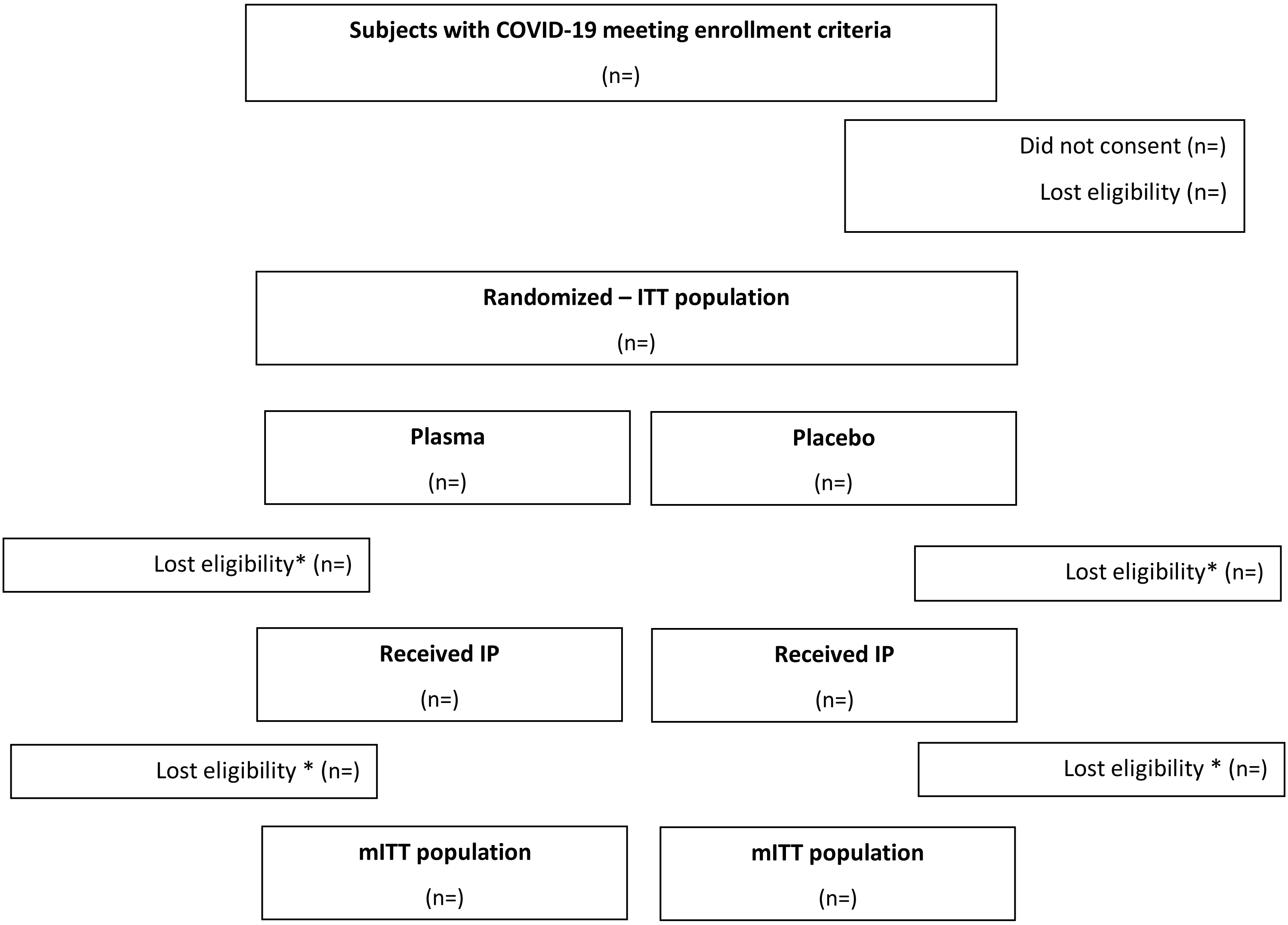


*Subjects that received the intervention but lost eligibility before being transfused (presence of primary outcome before transfusion)

Table 4: Characteristics of ITT population

| Variables | Plasma | | Placebo | |
|---|---|---|---|---|
| **Socio-demographic characteristics** | **n/N** | **%** | **n/N** | **%** |
| Age in years* | | | | |
| Age by categories | | | | |
| • 65-74 years | | | | |
| • ≥75 years | | | | |
| Sex | | | | |
| • Female | | | | |
| • Male | | | | |
| **Vital signs** | | | | |
| Axillary temperature* | | | | |
| Heart Rate* | | | | |
| Systolic blood pressure* | | | | |
| Diastolic blood pressure* | | | | |
| Respiratory rate* | | | | |
| Oxygen saturation at room air* | | | | |
| Hours from initiation of symptoms* | | | | |
| Respiratory rate (≥30 RPM) | | | | |
| Oxygen saturation < 93% | | | | |
| **Comorbidities** | | | | |
| Hypertension under treatment | | | | |
| Diabetes under treatment | | | | |
| Obesity | | | | |
| COPD under treatment | | | | |
| Cardiovascular disease** | | | | |
| Chronic renal failure | | | | |
| **Secondary comorbidities** | | | | |
| Asthma or other respiratory disease | | | | |
| Non-cirrhotic liver disease | | | | |
| Cancer (not active) | | | | |
| Neurologic disease | | | | |
| History of smoking | | | | |
| Receives medications regularly | | | | |
| Received medications in last 15 days | | | | |

* Media and Standard deviation

** Define as Known diagnosis of coronary disease, stroke history, congenital heart failure (EF < 40%).

Table 5: Characteristics of ITTm population

| Variables | Plasma | | Placebo | |
|---|---|---|---|---|
| **Características socio-demográficas** | **n/N** | **%** | **n/N** | **%** |
| **Variables** | | | | |
| **Socio-demographic characteristics** | | | | |
| • Age in years* | | | | |
| • Age by categories | | | | |
| 65-74 years | | | | |
| • ≥75 years | | | | |
| • Sex | | | | |
| Female | | | | |
| Male | | | | |
| **Vital signs** | | | | |
| Axillary temperature* | | | | |
| Heart Rate* | | | | |
| Systolic blood pressure* | | | | |
| Diastolic blood pressure* | | | | |
| Respiratory rate* | | | | |
| Oxygen saturation at room air* | | | | |
| Hours from initiation of symptoms* | | | | |
| Respiratory rate (≥30 RPM) | | | | |
| Oxygen saturation < 93% | | | | |
| **Comorbidities** | | | | |
| Hypertension under treatment | | | | |
| Diabetes under treatment | | | | |
| Obesity | | | | |
| COPD under treatment | | | | |
| Cardiovascular disease** | | | | |
| Chronic renal failure | | | | |
| **Secondary comorbidities** | | | | |
| Asthma or other respiratory disease | | | | |
| Non-cirrhotic liver disease | | | | |
| Cancer (not active) | | | | |
| Neurologic disease | | | | |
| History of smoking | | | | |

* Media and Standard deviation

** Define as Known diagnosis of coronary disease, stroke history, congenital heart failure (EF < 40%).

Table 6: Primary Endpoint. Proportion of cases - ITT

| | Plasma | | Placebo | | Relative Risk (95% CI) | P value |
|---|---|---|---|---|---|---|
| | n/N | % | n/N | % | | |
| **Severe Respiratory Disease** | | | | | | |

Table 7: Primary Endpoint: Time to event - ITT

| | Plasma | | Placebo | | P value* |
|---|---|---|---|---|---|
| | Median | (Q1-Q3) | Median | (Q1-Q3) | |
| **Severe Respiratory Disease** | | | | | |

*Log Rank Test

Figure 2: Kaplan-Meier survival curve of severe respiratory disease due to SARS-CoV2 by treatment group – ITT Analysis

Table 8: Primary outcome: Proportion of primary endpoint cases by age group - ITT

| | Plasma | | Placebo | | Relative Risk (95% CI) | P value |
|---|---|---|---|---|---|---|
| | n/N | % | n/N | % | | |
| **Severe Respiratory Disease** | | | | | | |
| • 65-74 years | | | | | | |
| • ≥75 years | | | | | | |

Table 9: Primary outcome. Time to event by group age - ITT

| | Plasma | | Placebo | | P value |
|---|---|---|---|---|---|
| | Median | (Q1-Q3) | Median | (Q1-Q3) | |
| **Severe Respiratory Disease** | | | | | |
| • 65-74 years | | | | | |
| • ≥75 years | | | | | |

*Log Rank Test

Figure 3: Kaplan-Meier survival curve of severe respiratory disease due SARS-CoV2 in 65-74 years old by treatment group – ITT Analysis

Figure 4: Kaplan-Meier survival curve of severe respiratory disease due SARS-CoV2 in ≥75 years old by treatment group – ITT Analysis

Table 10: Secondary outcomes: Case proportions –ITT

| | **Plasma** | | **Placebo** | | **Relative Risk (95% CI)** | **P value** |
|---|---|---|---|---|---|---|
| | **n/N** | **%** | **n/N** | **%** | | |
| Respiratory rate ≥30 x m | | | | | | |
| Oxygen saturation <93% | | | | | | |
| Support with maximum oxygen therapy and/ or non-invasive respiratory support and/or admission to UCI and/or invasive mechanical ventilation assistance requirement | | | | | | |
| • 100% oxygen support | | | | | - | |
| • Non-invasive respiratory support | | | | | - | |
| • Admission to UCI | | | | | - | |
| • Mechanical ventilation | | | | | - | |
| Critical systemic Illness | | | | | | |
| • Acute respiratory failure | | | | | - | |
| • Shock | | | | | - | |
| • Multiorganic shock syndrome | | | | | - | |
| Mortality due to SARS-CoV2 | | | | | | |
| Support with maximum oxygen therapy and/ or non-invasive respiratory support and/or admission to UCI and/or invasive mechanical ventilation assistance requirement and/or severe disease and/or mortality | | | | | | |
| Duration in days of the oxygen support requirement *Ω | | | | | | |
| Duration in days of the oxygen support requirement *B | | | | | | |

* Median and quartiles (quartile 1 – quartile 3)

Ω Only those who oxygen required were considered (n=in Group A y n=in Group B)

B Only those who oxygen required and survived were considered (n=in Group A y n=in Group B)

Table 11: Secondary outcomes: Time to event – ITT analysis

| | Plasma | | Placebo | | P value |
|---|---|---|---|---|---|
| | **Median** | **(Q1-Q3)** | **Median** | **(Q1-Q3)** | |
| Respiratory rate ≥30 x m | | | | | |
| Oxygen saturation <93% | | | | | |
| Support with maximum oxygen therapy and/ or non-invasive respiratory support and/or admission to UCI and/or invasive mechanical ventilation assistance requirement | | | | | |
| • 100% oxygen support | | | | | - |
| • Non-invasive respiratory support | | | | | - |
| • Admission to UCI | | | | | - |
| • Mechanical ventilation | | | | | - |
| Critical systemic Illness | | | | | |
| • Acute respiratory failure | | | | | - |
| • Shock | | | | | - |
| • Multiorganic shock syndrome | | | | | - |
| Mortality due to SARS-CoV2 | | | | | |
| Support with maximum oxygen therapy and/ or non-invasive respiratory support and/or admission to UCI and/or invasive mechanical ventilation assistance requirement and/or severe disease and/or mortality | | | | | |
| Duration in days of the oxygen support requirement *Ω | | | | | |
| Duration in days of the oxygen support requirement *B | | | | | |

* Median and quartiles (quartile 1 – quartile 3)

Ω Only those who oxygen required were considered (n=in Group A y n=in Group B)

B Only those who oxygen required and survived were considered (n=in Group A y n=in Group B)

Figure 5: Kaplan- Meier survival curve. Time to support with maximum oxygen therapy and/ or non-invasive respiratory support and/or admission to UCI and/or invasive mechanical ventilation assistance requirement by treatment group- ITT analysis

Figure 6: Kaplan- Meier survival curve. Time to critical systemic Illness by treatment group- ITT analysis

Figure 7: Kaplan- Meier survival curve. Time to mortality due to COVID-19 by treatment group- ITT analysis

Figure 8: Kaplan- Meier survival curve. Time to Support with maximum oxygen therapy and/ or non-invasive respiratory support and/or admission to UCI and/or invasive mechanical ventilation assistance requirement and/or critical systemic Illness and/or mortality – ITT analysis

Table 12: Requested adverse events – ITT analysis

| | Serious | | | | Non- serious | | | |
|---|---|---|---|---|---|---|---|---|
| | Plasma | | Placebo | | Plasma | | Placebo | |
| | n/N | % | n/N | % | n/N | % | n/N | % |
| Volume overload | | | | | | | | |
| Allergic reaction | | | | | | | | |
| Trombophlebitis | | | | | | | | |
| Vasovagal syndrome | | | | | | | | |
| Hematoma at site | | | | | | | | |
| Nerve injury | | | | | | | | |
| Tetany (hyperventilation) | | | | | | | | |
| Total events* | | | | | | | | |

* Median and quartiles (quartile 1 – quartile 3)

Table 13: Primary Endpoint. Proportion of cases - ITTm

| | Plasma | | Placebo | | Relative Risk (95% CI) | P value |
|---|---|---|---|---|---|---|
| | n/N | % | n/N | % | | |
| **Severe Respiratory Disease** | | | | | | |

Table 14: Primary Endpoint: Time to event - ITTm

| | Plasma | | Placebo | | P value* |
|---|---|---|---|---|---|
| | Median | (Q1-Q3) | Median | (Q1-Q3) | |
| **Severe Respiratory Disease** | | | | | |

*Log Rank Test

Figure 9: Kaplan-Meier survival curve of severe respiratory disease due to SARS-CoV2 by treatment group – ITTm Analysis

Table 15: Primary outcome: Proportion of primary endpoint cases by age group - ITTm

| | Plasma | | Placebo | | P value |
|---|---|---|---|---|---|
| | Median | (Q1-Q3) | Median | (Q1-Q3) | |
| **Severe Respiratory Disease** | | | | | |
| • 65-74 years | | | | | |
| • ≥75 years | | | | | |

Table 16: Primary outcome. Time to event by age group - ITTm

| | Plasma | | Placebo | | P value |
|---|---|---|---|---|---|
| | Median | (Q1-Q3) | Median | (Q1-Q3) | |
| **Severe Respiratory Disease** | | | | | |
| • 65-74 years | | | | | |
| • ≥75 years | | | | | |

*Log Rank Test

Figure 10: Kaplan-Meier survival curve of severe respiratory disease due SARS-CoV2 in 65-74 years old by treatment group – ITTm analysis

Figure 11: Kaplan-Meier survival curve of severe respiratory disease due SARS-CoV2 in ≥75 years old by treatment group – ITTm analysis

Table 17: Secondary Outcomes: Case proportions – ITTm analysis

| | **Plasma** | | **Placebo** | | **Relative Risk** | **P value** |
|---|---|---|---|---|---|---|
| | **n/N** | **%** | **n/N** | **%** | **(95% CI)** | |
| Respiratory rate ≥30 x m | | | | | | |
| Oxygen saturation <93% | | | | | | |
| Support with maximum oxygen therapy and/ or non-invasive respiratory support and/or admission to UCI and/or invasive mechanical ventilation assistance requirement | | | | | | |
| • 100% oxygen support | | | | | - | |
| • Non-invasive respiratory support | | | | | - | |
| • Admission to UCI | | | | | - | |
| • Mechanical ventilation | | | | | - | |
| Critical systemic Illness | | | | | | |
| • Acute respiratory failure | | | | | - | |
| • Shock | | | | | - | |
| • Multiorganic shock syndrome | | | | | - | |
| Mortality due to SARS-CoV2 | | | | | | |
| Support with maximum oxygen therapy and/ or non-invasive respiratory support and/or admission to UCI and/or invasive mechanical ventilation assistance requirement and/or severe disease and/or mortality | | | | | | |
| Duration in days of the oxygen support requirement *Ω | | | | | | |
| Duration in days of the oxygen support requirement *B | | | | | | |

* Median and quartiles (quartile 1 – quartile 3)

Ω Only those who oxygen required were considered (n=in Group A y n=in Group B)

B Only those who oxygen required and survived were considered (n=in Group A y n=in Group B)

Table 18: Secondary outcomes: Time to event – ITTm analysis

| | Plasma | | Placebo | | P value |
|---|---|---|---|---|---|
| | Median | (Q1-Q3) | Median | (Q1-Q3) | |
| Respiratory rate ≥30 x m | | | | | |
| Oxygen saturation <93% | | | | | |
| Support with maximum oxygen therapy and/ or non-invasive respiratory support and/or admission to UCI and/or invasive mechanical ventilation assistance requirement | | | | | |
| • 100% oxygen support | | | | | - |
| • Non-invasive respiratory support | | | | | - |
| • Admission to UCI | | | | | - |
| • Mechanical ventilation | | | | | - |
| Critical systemic Illness | | | | | |
| • Acute respiratory failure | | | | | - |
| • Shock | | | | | - |
| • Multiorganic shock syndrome | | | | | - |
| Mortality due to SARS-CoV2 | | | | | |
| Support with maximum oxygen therapy and/ or non-invasive respiratory support and/or admission to UCI and/or invasive mechanical ventilation assistance requirement and/or severe disease and/or mortality | | | | | |
| Duration in days of the oxygen support requirement *Ω | | | | | |
| Duration in days of the oxygen support requirement *B | | | | | |

* Median and quartiles (quartile 1 – quartile 3)

Ω Only those who oxygen required were considered (n=in Group A y n=in Group B)

B Only those who oxygen required and survived were considered (n=in Group A y n=in Group B)

Figure 12: Kaplan- Meier survival curve. Time to support with maximum oxygen therapy and/ or non-invasive respiratory support and/or admission to UCI and/or invasive mechanical ventilation assistance requirement by treatment group – ITTm analysis

Figure 13: Kaplan- Meier survival curve. Time to critical systemic Illness by treatment group – ITTm Analysis

Figura 14: Kaplan- Meier survival curve. Time to mortality due to COVID-19 by treatment group- ITT analysis – ITTm Analysis

Figura 15: Kaplan- Meier survival curve. Time to Support with maximum oxygen therapy and/ or non-invasive respiratory support and/or admission to UCI and/or invasive mechanical ventilation assistance requirement and/or critical systemic Illness and/or mortality – ITTm analysis

Table 19: Requested adverse events – ITTm analysis

| | **Serious** | | | | **Non- serious** | | | |
|---|---|---|---|---|---|---|---|---|
| | **Plasma** | | **Placebo** | | **Plasma** | | **Placebo** | |
| | **n/N** | **%** | **n/N** | **%** | **n/N** | **%** | **n/N** | **%** |
| Cardiac volume overload associated with the transfusion | | | | | | | | |
| Allergic or anaphylactic reaction | | | | | | | | |
| Thrombophlebitis | | | | | | | | |
| Vasovagal syndrome | | | | | | | | |
| Hematoma at site | | | | | | | | |
| Nerve injury | | | | | | | | |
| Tetany (hyperventilation) | | | | | | | | |
| Total events* | | | | | | | | |

* Median and quartiles (quartile 1 – quartile 3)
